# Frontline Healthcare workers suffering from psychosomatic disorders during COVID-19 (a pandemic) – A Systematic review

**DOI:** 10.1101/2021.11.09.21266105

**Authors:** Rhythm Joshi, Nidhi.B. Agarwal, Dinesh Bhurani, Mohd Ashif Khan

## Abstract

**Purpose:** The emergence of SARS CoV-2, has imposed high pressure on the healthcare system worldwide. As a consequence, frontline healthcare workers were impacted widely. The aim of this systematic review is to examine the impact of COVID-19 on mental status of FHW during pandemic.

**Methods:** Databases such as PubMed, Scopus, google scholar were searched extensively from the date of inception till April 2021. All cross-sectional studies published in English assessing the mental condition and well-being of frontline caregivers during COVID-19 were included in the study. The quality assessment was done by Newcastle Ottawa scale.

**Results:** Ten thousand eight hundred sixty-nine articles were found. After conscientious literature search, total 78 articles were included satisfying the objective of the review. The highest and lowest values for the rates of depression, anxiety and insomnia was found to be 99.51% & 6.07%, 85.7% & 73.6%, and 5.3% & 11.4%, respectively.

**Conclusion:** It has been found that FHW were psychologically impacted by the pandemic. This could be due to lack of resources such as PPE, organizational support, inefficient relevant knowledge regarding the novel virus, its extremely indelible transmission rates, fear of contamination, stigmatization, and/or due to prevalence of ignorance by government and health policy makers.

**Prospero registration no-** CRD42021244612

## 1. INTRODUCTION

Coronavirus belong to the family Coronaviridae and order Nidovirales. These are non-segmented, enveloped and single stranded RNA viruses classified under genus coronavirus (P. Chen et al., 2020). Coronavirus disease (COVID-19), whose first case reported in Hubei Province of Wuhan, China in November 2019, has created havoc in lives of people all around the world.

COVID-19 pandemic has led to the worst crisis globally affecting more than 5,378 million people and mortality rate of 1.3 million people as of 14 Nov 2020 (C et al., 2020).

Healthcare workers (HCWs) are at risk of developing mental distress and other psychological symptoms due to high work demand (Fauzi et al., 2020). Moreover, recently various studies have reported the prevalence of depression, anxiety, and insomnia among HCW due to current pandemic (de Burgos-Berdud et al., 2021; Santabárbara et al., 2021).

High transmission and mortality rate of COVID-19 among FHW may be one of the reasons for the prevalence of psychiatric morbidity among them. According to reports, by the end of Feb, 2020 more than 3000 HCW in China got transmitted with the virus (Gan et al., 2020) with the prevalence rate of depression, anxiety, and stress-related symptoms to be 50%, 44.7%, and 73.4%, respectively (Elbay et al., 2020a). Also, Italy faced the death of almost 100 physicians and 26 nurses and over 12,680 HCW were infected from COVID-19 till 7th April 2020 (Barranco and Ventura, 2020). France reported high infection rates among HCWs and more than 50 deaths (McKay et al., 2020; Shaukat et al., 2020) Challenges were knocked together for HCWs due to rapid spread of the disease and delayed onset of symptoms in infected patients or asymptomatic carriers (Hall, 2020; McKay et al., 2020; Shaukat et al., 2020).

Conditions similar to that of battle field are being seen by HCWs in these unprecedented challenges. High levels of anxiety and uncertainties are being experienced and thus persistent risks of development of stress exposure syndrome and professional burnout are prevailing among those working in frontline]. (Albott et al., 2020; Antonijevic et al., 2020a; Kannampallil et al., 2020; Keubo et al., 2020).

In this phase of COVID-19 attack, HCW being the frontline warriors are the ones who are least talked about.

In this century world has not faced a pandemic as big as COVID-19, which is also new for HCWs, thus, it is need of today to explore the psychological crisis they are being facing (Thamer H. Alenazi et al., 2020). Hence, this systematic review aims to report the mental health condition of HCW during COVID-19.

## 2. METHOD

### 2.1 Data research strategy

This study was conducted following PRISMA (Preferred Reporting Items for Systematic Reviews and Meta-analysis) guidelines using suitable keywords such as [Mental disorders OR Depression OR Anxiety OR Bipolar disorder OR Cognitive impairment OR Neurodegenerative disorders] AND [Healthcare workers OR Medical professional OR Doctors OR Nurses] AND [COVID-19] in PubMed, Embase/Elsevier, Cochrane, Science direct and Google scholar from inception till April 2021. Additionally, citations of the included studies were screened manually for the identification of relevant studies. Language of the articles was restricted to English.

### 2.2 Selection criteria for included studies

Articles fulfilling the following criteria were included: 1. Studies which examined the impact of SARS-CoV-2 on mental health of healthcare professionals; 2. Studies which investigated mental health of FHW; 3. Use of at least one validated quantitative scoring scale to measure mental health outcomes; 4. Available in English Language; 5. Hospital based studies; 6. Conducted from 31st December 2019 (when China reported the first case of SARS-CoV-2 in Wuhan) till April 2021; 7. Original research (i.e., commentaries, editorials and reviews were excluded), be published in peer reviewed journals. Inclusion of studies in the present systematic review is depicted using PRISMA flowchart (Figure 1).

**Figure 1.**
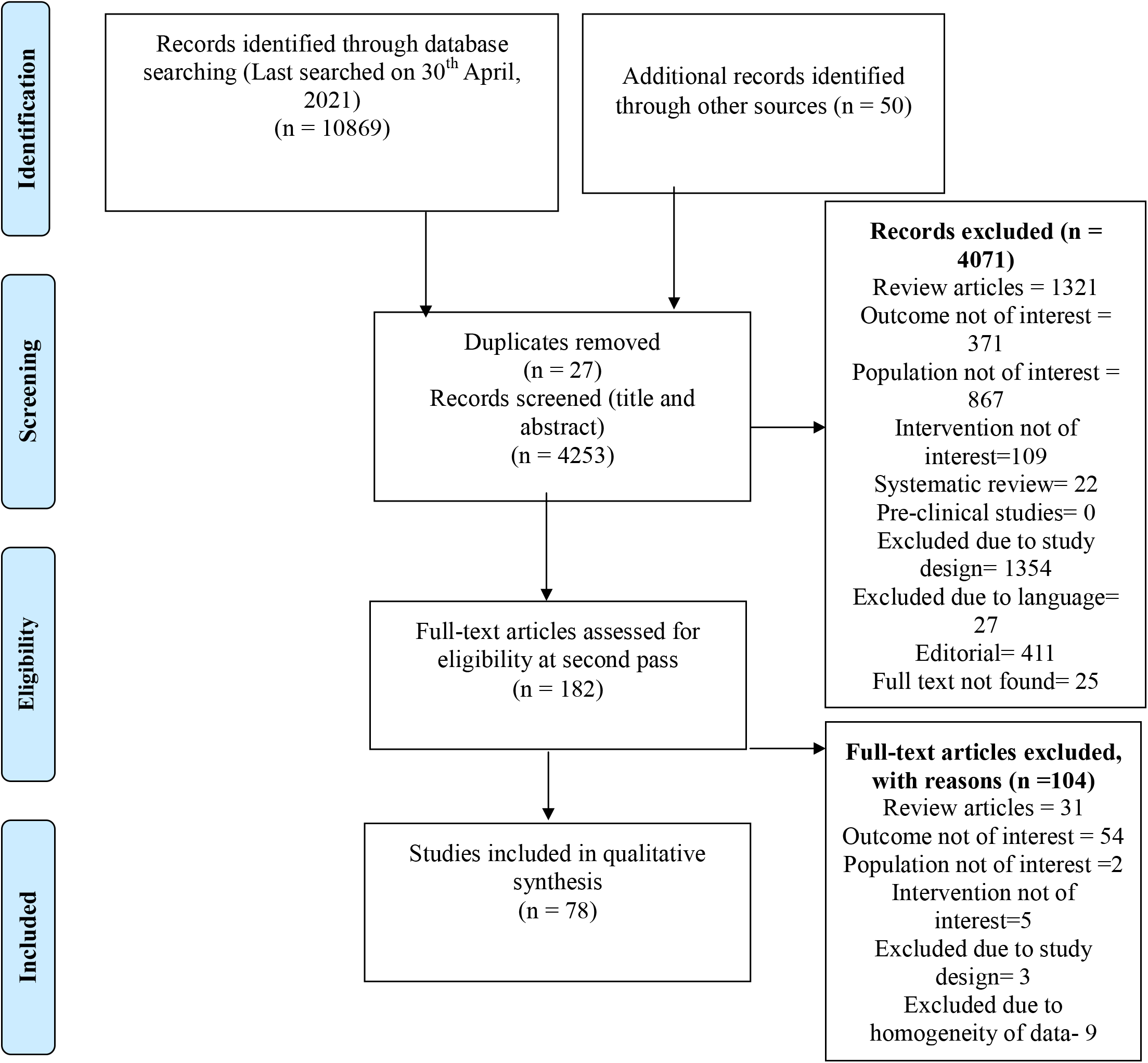
PRISMA Flow diagram showing study selection process

Studies investigating non-hospital–based HCWs, published in languages other than English or having fewer than 20 participants, and editorials or protocols were excluded.

### 2.3 Data derivation and evaluation of study trait

To extract the desirable data, structured format was used and information such as study country, population size, type of mental health, measurement tool used for mental health, severity of outcome, and prevalence rate were tabulated.

### 2.4 Outcomes

The objective of this systematic review was to assess the prevalence of depression, anxiety, insomnia, stress and distress among FHW during COVID-19..

## 3. RESULTS

### 3.1 Literature search

Initially, 10,919 records were found. Out of these, 27 duplicates were removed and 4253 potential studies (title and abstract) were screened. Further, after the exclusion of 4071 articles, full texts of 182 articles were assessed.. Finally, 78 articles were included in the study (Fig. 1) (AlAteeq et al., 2020; Thamer H Alenazi et al., 2020; Alshekaili et al., 2020a; Altintas, 2017; An et al., 2020; Antonijevic et al., 2020a; Ayhan Başer et al., 2020; Azoulay et al., n.d.; Bhargava et al., 2020; Buselli et al., 2020; Caillet et al., 2020a; X. Chen et al., 2020; Kannampallil et al., 2020; Keubo et al., 2020; Liu et al., 2020) (Altintas, 2017)(AlAteeq et al., 2020; Thamer H Alenazi et al., 2020; Alshekaili et al., 2020a)(An et al., 2020; Antonijevic et al., 2020b; Ayhan Başer et al., 2020; Azoulay et al., n.d.; Bhargava et al., 2020; Buselli et al., 2020; Caillet et al., 2020a; X. Chen et al., 2020; Liu et al., 2020)(W. Cai et al., 2020; J. Chen et al., 2020; Q. Chen et al., 2020; X. Chen et al., 2020; Cheng et al., 2020; Chew et al., 2020; Di Tella et al., 2020; Elbay et al., 2020b; Elhadi et al., 2020b; Fadhli et al., n.d.; Gallopeni et al., 2020; Herrero San Martin et al., 2020; Hong et al., 2020; Huang and Zhao, 2020; Janati Idrissi et al., 2020; Johnson et al., 2020; Kang et al., 2020a; Kannampallil et al., 2020; Keubo et al., 2020; Koksal et al., 2020a; Korkmaz et al., 2020; Labrague and De los Santos, 2020; J. Lai et al., 2020; G. Li et al., 2020; R. Li et al., 2020; Liang et al., 2020; M. Liu et al., 2020; Y. Liu et al., 2020a; Lu et al., 2020; Luceño-Moreno et al., 2020; Nie et al., 2020; Ning et al., 2020; Prasad et al., 2020; Que et al., 2020; Rossi et al., 2020; Şahin et al., 2020a; Salopek-Žiha et al., 2020; Sandesh et al., 2020; Shah et al., 2020; Shahrour and Dardas, 2020; Shechter et al., 2020; Si et al., 2020; Skoda et al., 2020; Stojanov et al., 2020; Suryavanshi et al., 2020; Teo et al., 2020; Tian et al., 2020; Wang et al., 2020, 2020; L. Q. Wang et al., 2020; Wańkowicz et al., 2020; Wilson et al., 2020; Xiao et al., 2020; Xing et al., 2020; Yáñez et al., 2020; Yang et al., 2020a; Yin et al., 2020; W. R. Zhang et al., 2020)

### 3.2 Study characteristics

Seventy-eight studies met the inclusion criteria for this systematic review. All studies quantitively synthesize data of the psychological illness such as anxiety, depression, stress disorders, insomnia and PTSD affecting FHW due to COVID-19.

Out of 78 studies, 35 were from China; 4 studies were conducted in USA and Turkey; 3 studies were from Saudi Arabia and Italy; 2 studies were from Serbia, Singapore, Pakistan and India; 1 study each from Oman, Malaysia, Kosovo, Morocco, Norway, Cameroon, Philippines, Croatia, Jordan, Germany, Poland, Peru, Iran and United kingdoms. One study included the dermatologists from all the continents-Asia, North America, Europe and others. (Altintas, 2017)(An et al., 2020)(Q. Chen et al., 2020; X. Chen et al., 2020; Cheng et al., 2020)(G. Li et al., 2020; Wang et al., 2020)(Huang and Zhao, 2020; Juan et al., 2020; Wang et al., 2020)(Kang et al., 2020b; J. Lai et al., 2020; X. Lai et al., 2020; Liang et al., 2020; Liu et al., 2020; Y. Liu et al., 2020b)(Liu et al., 2020; Nie et al., 2020; Ning et al., 2020; L. Q. Wang et al., 2020), (Tian et al., 2020)(W. Cai et al., 2020; Xiao et al., 2020)(Si et al., 2020; Xiaoming et al., 2020)(Y. Liu et al., 2020b; Xiaoming et al., 2020; Xing et al., 2020; Yang et al., 2020b)(Yin et al., 2020; M. Zhan et al., 2020; S. X. Zhang et al., 2020; Yifang Zhou et al., 2020)(B. Zhou et al., 2020; Zhou et al., 2020, 2020; Yifang Zhou et al., 2020; Z. Zhu et al., 2020b).(Civantos et al., 2020; Kannampallil et al., 2020; Prasad et al., 2020; Shechter et al., 2020)(Ayhan Başer et al., 2020; Koksal et al., 2020b; Korkmaz et al., 2020)(Şahin et al., 2020b)(Buselli et al., n.d.; Di Tella et al., 2020; Rossi et al., 2020)(AlAteeq et al., 2020; Thamer H Alenazi et al., 2020)

Sample size of the study ranged from 107 (Altintas, 2017) – 8817 (Xiaoming et al., 2020) with an average age of 19-60< years.

### 3.3 Risk of Bias assessment/ Quality assessment

The quality of included articles was assessed using Newcastle-Ottawa Scale (NOS). To remove the risk of bias, quality assessment was done by two authors (RJ and MAK) independently. The quality was based on three categories i.e., selection, comparability and outcome consisting of 7 questions. In this, a study can be awarded a maximum of one star for each numbered item within the Selection and Exposure categories. A maximum of two stars can be given for Comparability. Later, stars of specific question can be added later or total score would be the number of stars given to the specific set of questions in a category. Scores ranging 7-10 were deemed to be of high quality, 6-7 represent medium quality and 1-6 were considered to be of low quality (Table-1).

**Table 1.** Quality of assessment table

| Sno. | Author and year of study | Selection | Comparability | Outcome | Total | Quality of the study |
| --- | --- | --- | --- | --- | --- | --- |
| 1 | (Altintas, 2017), 2017, Saudi Arabia | 4 | 2 | 3 | 9 | High |
| 2 | (Alshekaili et al., 2020b)Oman | 4 | 2 | 3 | 9 | High |
| 3 | (AlAteeq et al., 2020),2020, Saudi Arabia | 4 | 2 | 3 | 9 | High |
| 4 | (Thamer H Alenazi et al., 2020)2020, Saudi Arabia | 5 | 2 | 2 | 9 | High |
| 5 | (An et al., 2020)2020 China | 5 | 2 | 3 | 10 | High |
| 6 | (Antonijevic et al., 2020a)2020, Serbia | 4 | 2 | 2 | 9 | High |
| 7 | (Ayhan Başer et al., 2020)2020, TURKEY | 4 | 1 | 2 | 7 | Low |
| 8 | (Azoulay et al., n.d.)2020, France | 5 | 1 | 3 | 9 | High |
| 9 | (Bhargava et al., 2020)2020, Asia, North America, Central/ south America, Europe, Other. | 4 | 2 | 2 | 8 | High |
| 10 | (Buselli et al., n.d.)2020, ITALY | 4 | 1 | 2 | 7 | Low |
| 11 | (W. Cai et al., 2020), 2020, China | 5 | 1 | 2 | 8 | High |
| 12 | (Caillet et al., 2020a)2020, France | 5 | 2 | 2 | 9 | High |
| 13 | (Liu et al., 2020),<br>2020, China | 5 | 2 | 2 | 9 | High |
| 14 | (Civantos et al.,<br>2020)2020, US | 4 | 2 | 2 | 8 | High |
| 15 | (Cheng et al.,<br>2020).2020, China | 4 | 1 | 3 | 8 | High |
| 16 | (Chew et al.,<br>2020)2020,<br>Singapore | 5 | 2 | 2 | 9 | High |
| 17 | (Di Tella et al.,<br>2020)2020, Italy | 5 | 1 | 3 | 9 | High |
| 18 | (Elhadi et al.,<br>2020a)2020, Libya | 4 | 2 | 2 | 8 | High |
| 19 | (Fauzi et al.,<br>2020)2020,<br>Malaysia. | 5 | 1 | 2 | 8 | High |
| 20 | (Gallopini et al.,<br>2020)2020, Kosovo | 4 | 2 | 2 | 8 | High |
| 21 | (G. Li et al.,<br>2020)2020, China | 4 | 1 | 3 | 8 | High |
| 22 | (Herrero San Martin et al., 2020)2020, Spain | 5 | 1 | 2 | 8 | High |
| 23 | (Wang et al., 2020), 2020, China | 5 | 2 | 3 | 10 | High |
| 24 | (Hong et al., 2020)2020, China | 5 | 2 | 2 | 9 | High |
| 25 | (J. Zhu et al., 2020)2020, China | 5 | 2 | 2 | 9 | High |
| 26 | (Huang and Zhao, 2020)2020, China | 5 | 1 | 2 | 8 | High |
| 27 | (Janati Idrissi et al., 2020)2020, Morocco | 5 | 1 | 1 | 7 | Low |
| 28 | (Juan et al., 2020), 2020, China | 4 | 2 | 2 | 8 | High |
| 30 | (Kang et al., 2020a)China | 5 | 1 | 2 | 8 | High |
| 31 | (Kannampallil et al., 2020)USA | 4 | 2 | 2 | 8 | High |
| 32 | (Keubo et al., 2020), 2020, Cameroon | 5 | 1 | 3 | 9 | High |
| 33 | (Koksal et al., 2020a), 2020, Turkey | 4 | 2 | 3 | 9 | High |
| 34 | (Korkmaz et al., 2020)Turkey | 4 | 2 | 2 | 8 | High |
| 35 | (Labrague and De los Santos, 2020) Philippines | 4 | 2 | 3 | 9 | High |
| 36 | (J. Lai et al., 2020) , 2020, China | 4 | 2 | 2 | 8 | High |
| 37 | (Lee et al., 2020) Singapore | 4 | 1 | 2 | 7 | Low |
| 38 | (Liang et al., 2020), 2020 China | 4 | 2 | 3 | 9 | High |
| 39 | (Lu et al., 2020), 2020, China | 4 | 1 | 2 | 7 | Low |
| 40 | (Luceño-Moreno et al., 2020) 2020, Spain | 4 | 2 | 2 | 8 | High |
| 41 | (L. Q. Wang et al., 2020)2020, China | 4 | 1 | 2 | 7 | Low |
| 42 | (Nie et al., 2020) 2020, China | 4 | 1 | 2 | 7 | Low |
| 43 | (Ning et al., 2020)2020, China | 5 | 1 | 2 | 8 | High |
| 44 | (Pouralizadeh et al., 2020)2020, Iran | 5 | 2 | 3 | 10 |  |
| 45 | (Prasad et al., 2020)2020, USA | 4 | 1 | 3 | 8 | High |
| 46 | (Q. Chen et al., 2020),2020, China | 5 | 2 | 3 | 10 | High |
| 47 | (Que et al., 2020) 2020, China | 5 | 1 | 2 | 8 | High |
| 48 | (R. Li et al., 2020),<br>2020, China | 5 | 1 | 3 | 9 | High |
| 49 | (Rossi et al.,<br>2020)2020, Italy | 4 | 1 | 2 | 7 | Low |
| 50 | (Şahin et al.,<br>2020a)2020, Turkey | 5 | 1 | 2 | 8 | High |
| 51 | (Salopek-Žiha et al.,<br>2020)2020, Croatia | 5 | 2 | 2 | 9 | High |
| 52 | (Sandesh et al.,<br>2020)2020, Pakistan | 4 | 2 | 2 | 8 | High |
| 53 | (Shah et al.,<br>2020)2020, UK | 5 | 1 | 3 | 9 | High |
| 54 | (Shahrour,<br>2020)2020, Jordan | 4 | 2 | 2 | 8 | High |
| 55 | (Shechter et al.,<br>2020) 2020, NYC | 5 | 1 | 2 | 8 | High |
| 56 | (Si et al., 2020),<br>2020, China | 5 | 1 | 3 | 9 | High |
| 57 | (Skoda et al.,<br>2020)2020.,<br>Germany | 5 | 2 | 3 | 10 | High |
| 58 | (Stojanov et al.,<br>2020)2020, Serbia | 5 | 2 | 2 | 9 | High |
| 59 | (Suryavanshi et al.,<br>2020)2020, India | 4 | 2 | 2 | 8 | High |
| 60 | (Wang et al., 2020)<br>2020, China | 4 | 1 | 2 | 7 | Low |
| 61 | (Tian et al.,<br>2020)2020, China | 5 | 1 | 3 | 9 | High |
| 62 | (Wasim et al.,<br>2020), 2020,<br>Pakistan. | 4 | 2 | 2 | 8 | High |
| 63 | (Wańkowicz et al., 2020)2020, Poland | 4 | 1 | 3 | 8 | High |
| 64 | (Wilson et al., 2020), 2020, India | 5 | 2 | 2 | 9 | High |
| 65 | (X. Chen et al., 2020) 2020, China | 5 | 2 | 2 | 9 | High |
| 66 | (Xiao et al., 2020), 2020, China | 4 | 1 | 2 | 7 | Low |
| 67 | (Xiaoming et al., 2020)2020 China | 5 | 2 | 3 | 10 | High |
| 68 | (Xing et al., 2020), 2020, China | 4 | 2 | 2 | 8 | High |
| 69 | (Yang et al., 2020a)2020, China | 5 | 2 | 2 | 9 | High |
| 70 | (Yáñez et al., 2020), 2020, Peru | 4 | 1 | 2 | 7 | Low |
| 71 | (Y. Liu et al., 2020b), 2020, China | 5 | 2 | 2 | 9 | High |
| 72 | (Yin et al., 2020),<br>2020, China | 4 | 1 | 2 | 7 | Low |
| 73 | (Zhan et al., 2020),<br>2020, China | 5 | 2 | 3 | 10 | High |
| 74 | (Y. Wang et al.,<br>2020a), 2020, China | 5 | 1 | 3 | 9 | High |
| 75 | (Yifang Zhou et al.,<br>2020)China | 4 | 2 | 2 | 8 | High |
| 76 | (W. R. Zhang et al.,<br>2020), 2020, China | 5 | 1 | 3 | 9 | High |
| 77 | (Z. Zhu et al.,<br>2020a),2020, China | 4 | 2 | 3 | 8 | High |
| 78 | (Zhou et al.,<br>2020)2020, China | 4 | 2 | 2 | 8 | High |

### 3.4 Depression and Anxiety

Out of 78 studies, 56 articles discussed about the anxiety and depression, suffered by the FHW staff (Table 2).

**Table 2.**
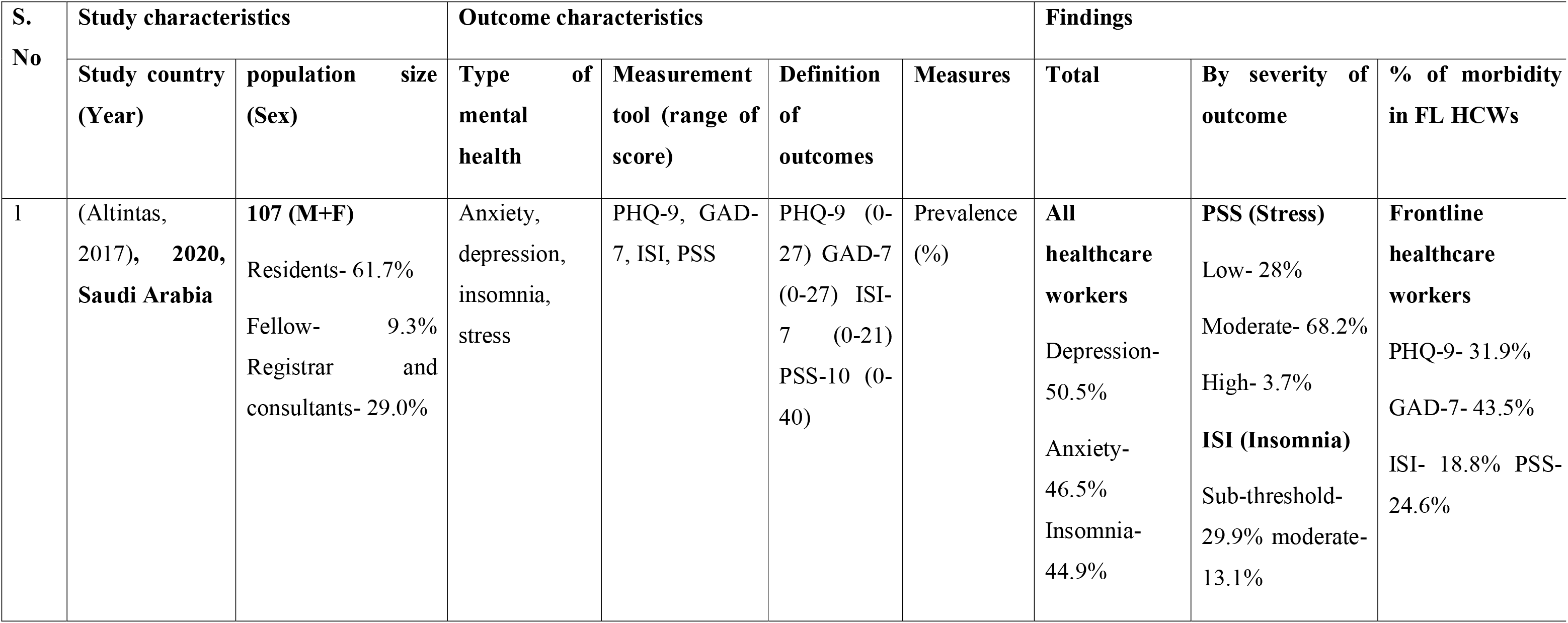
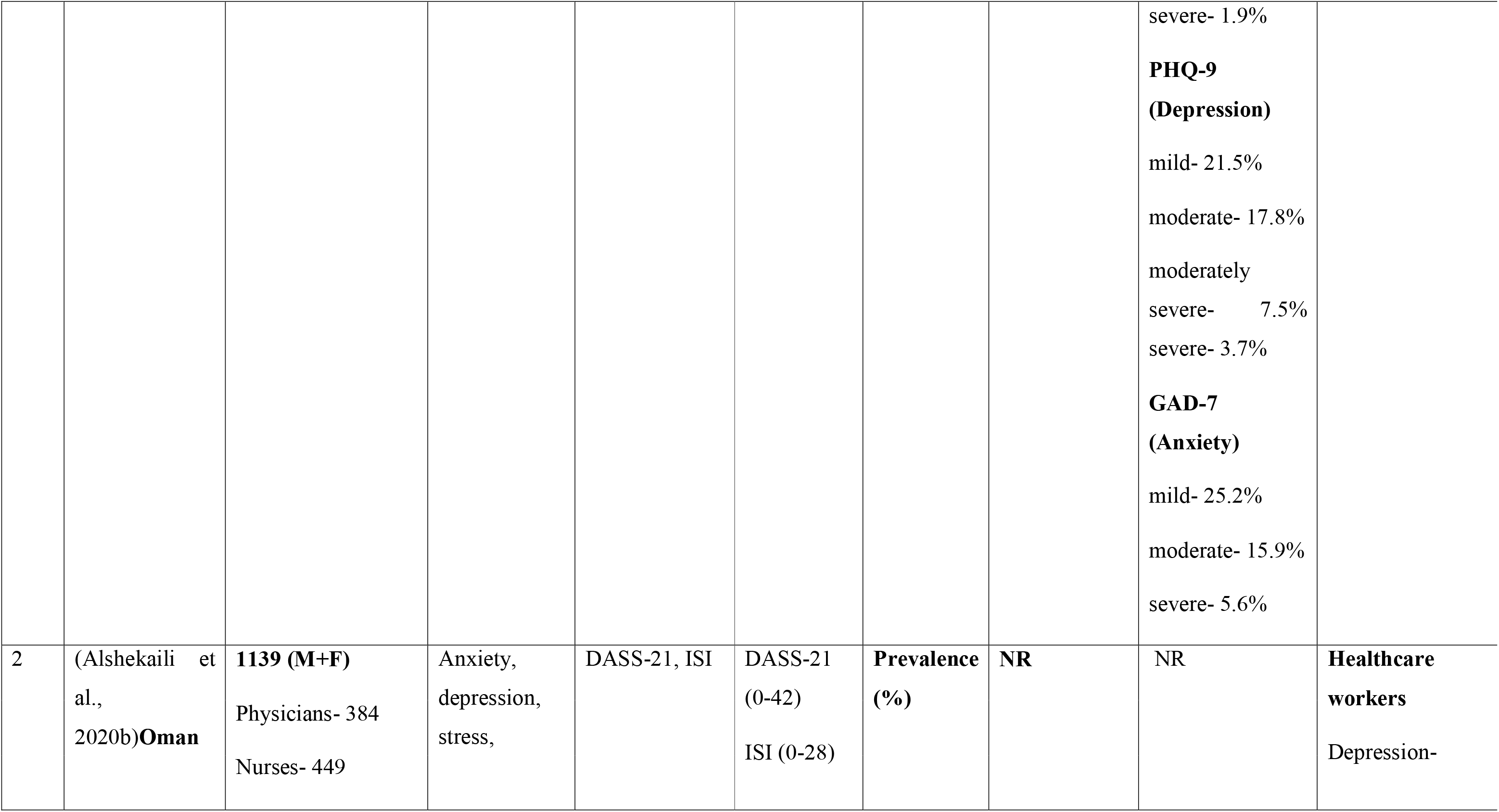
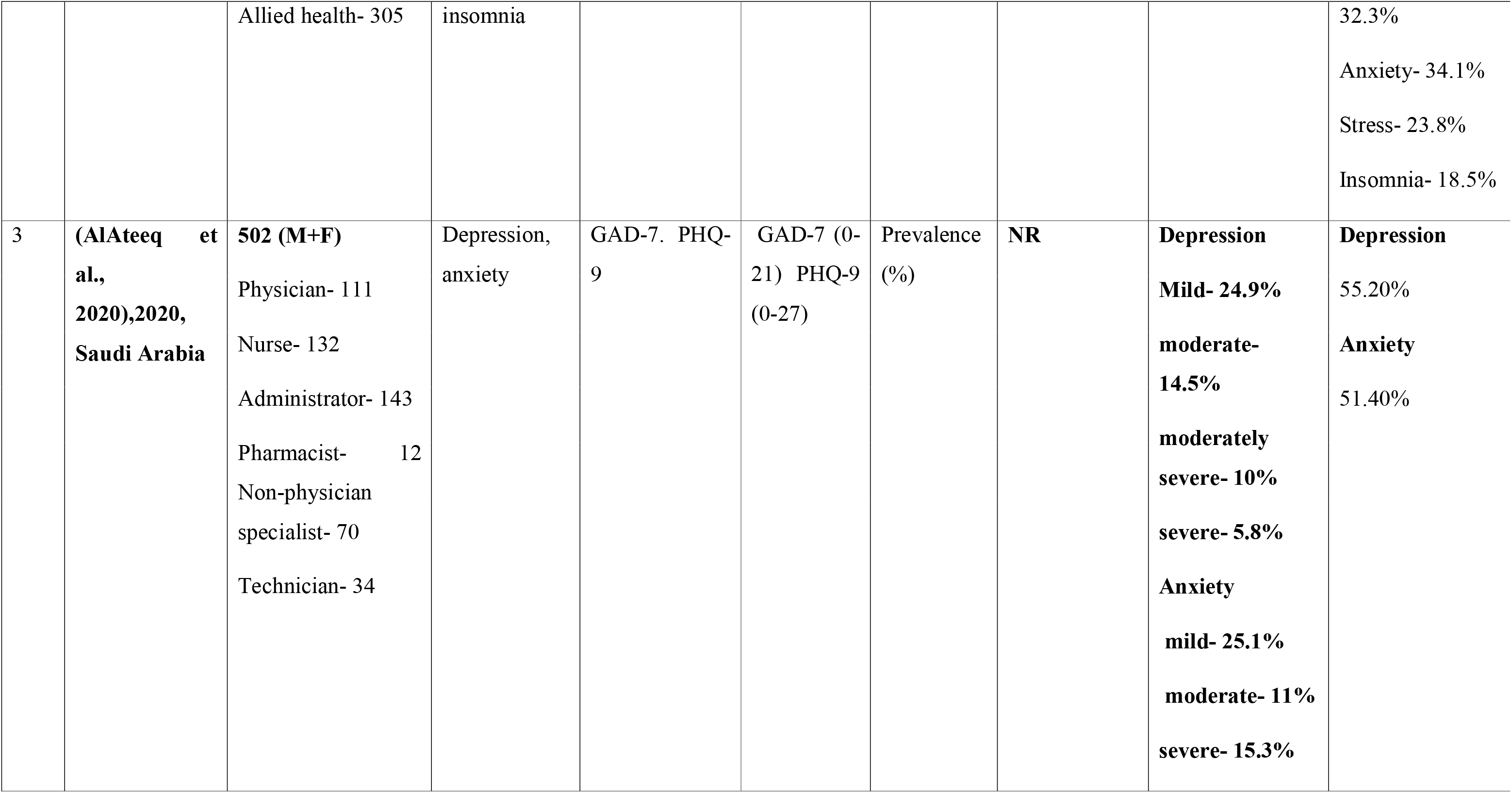
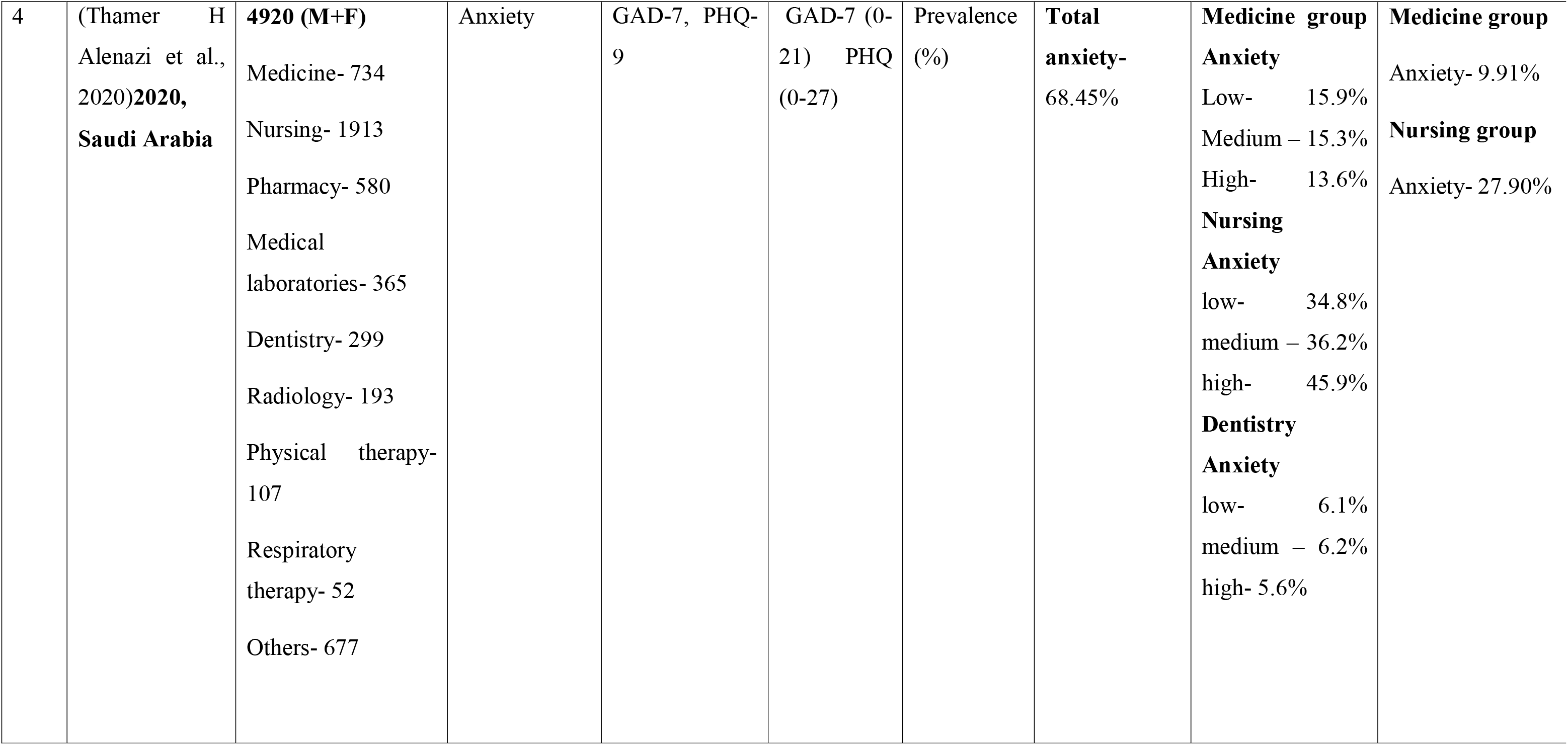
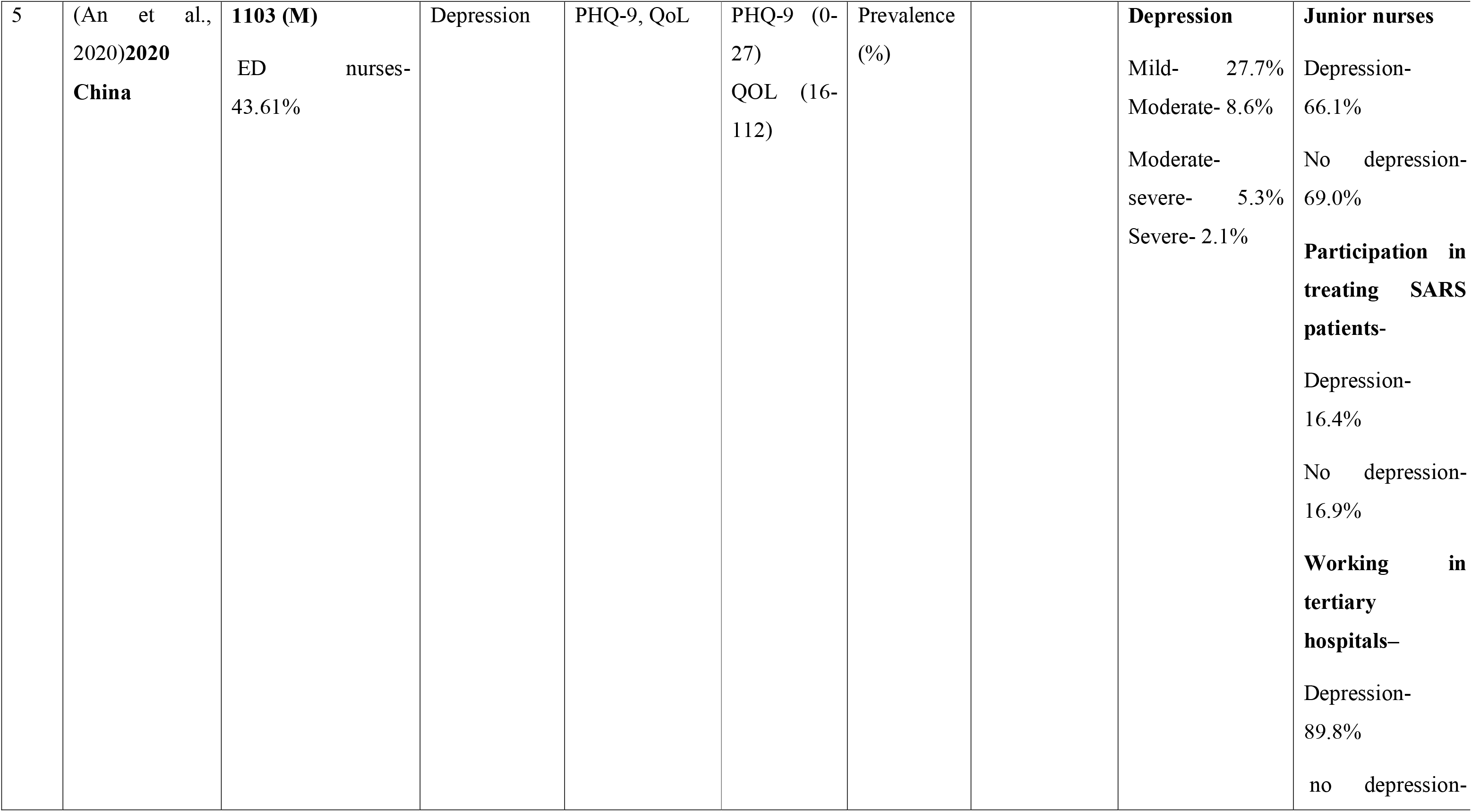
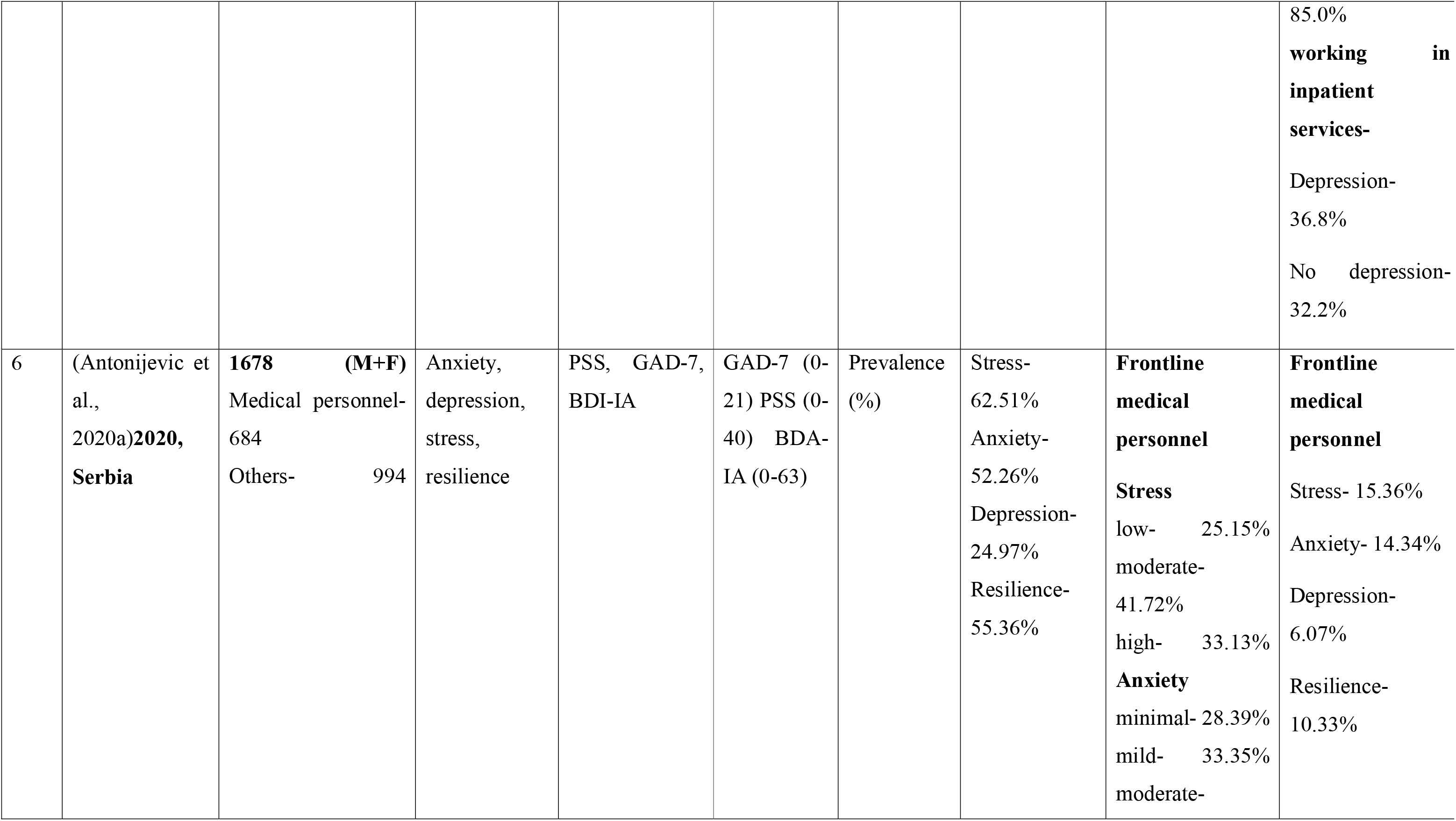
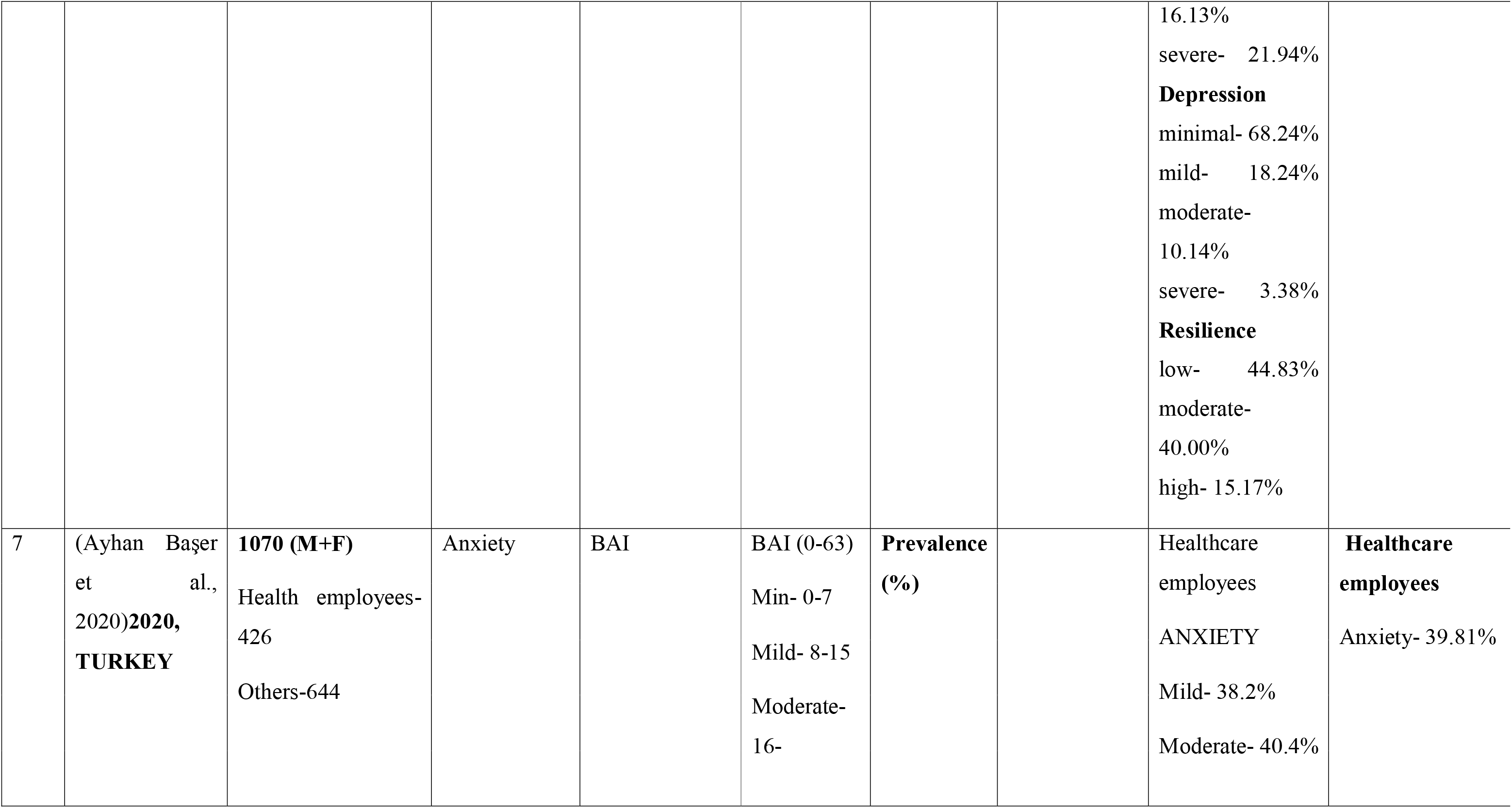
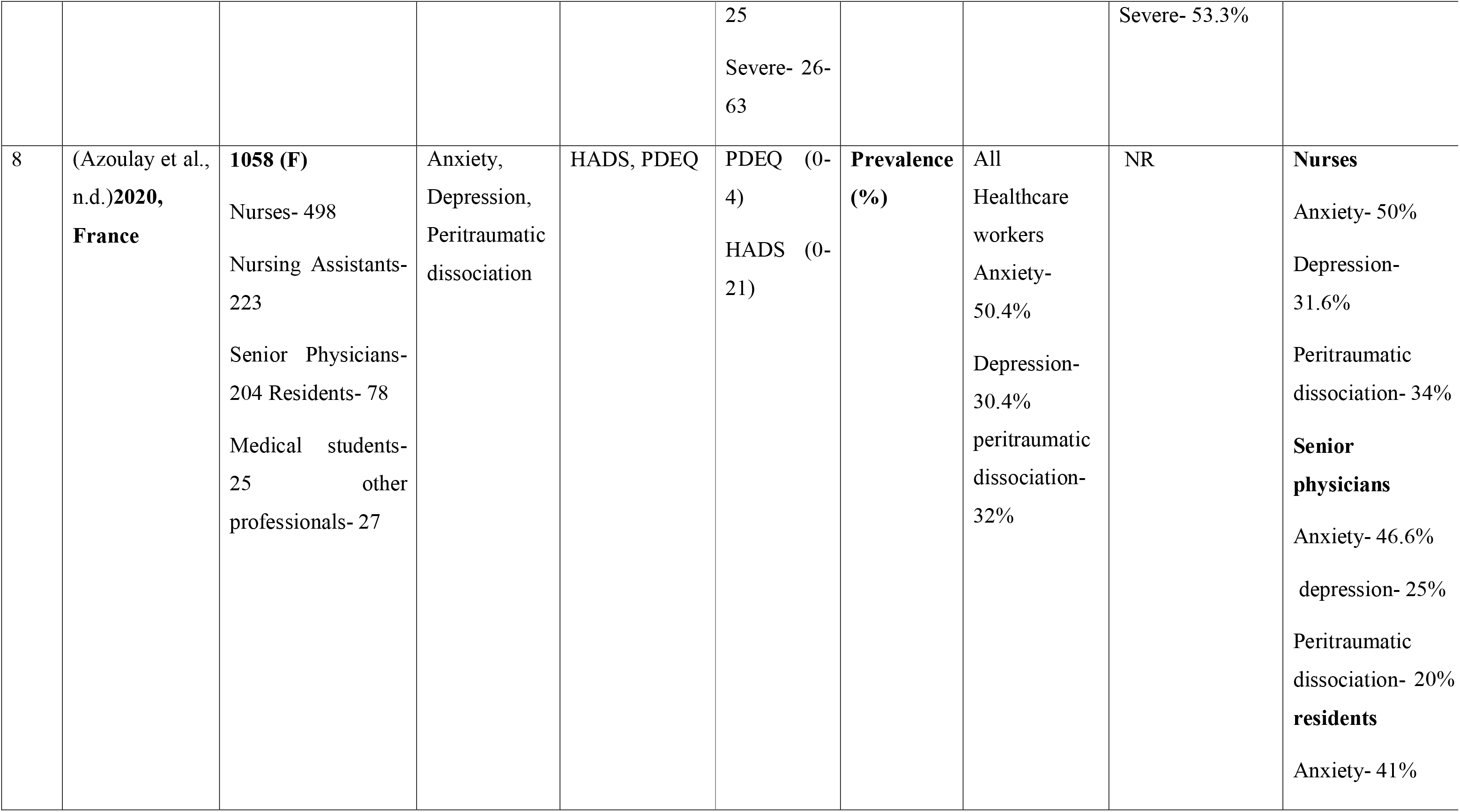
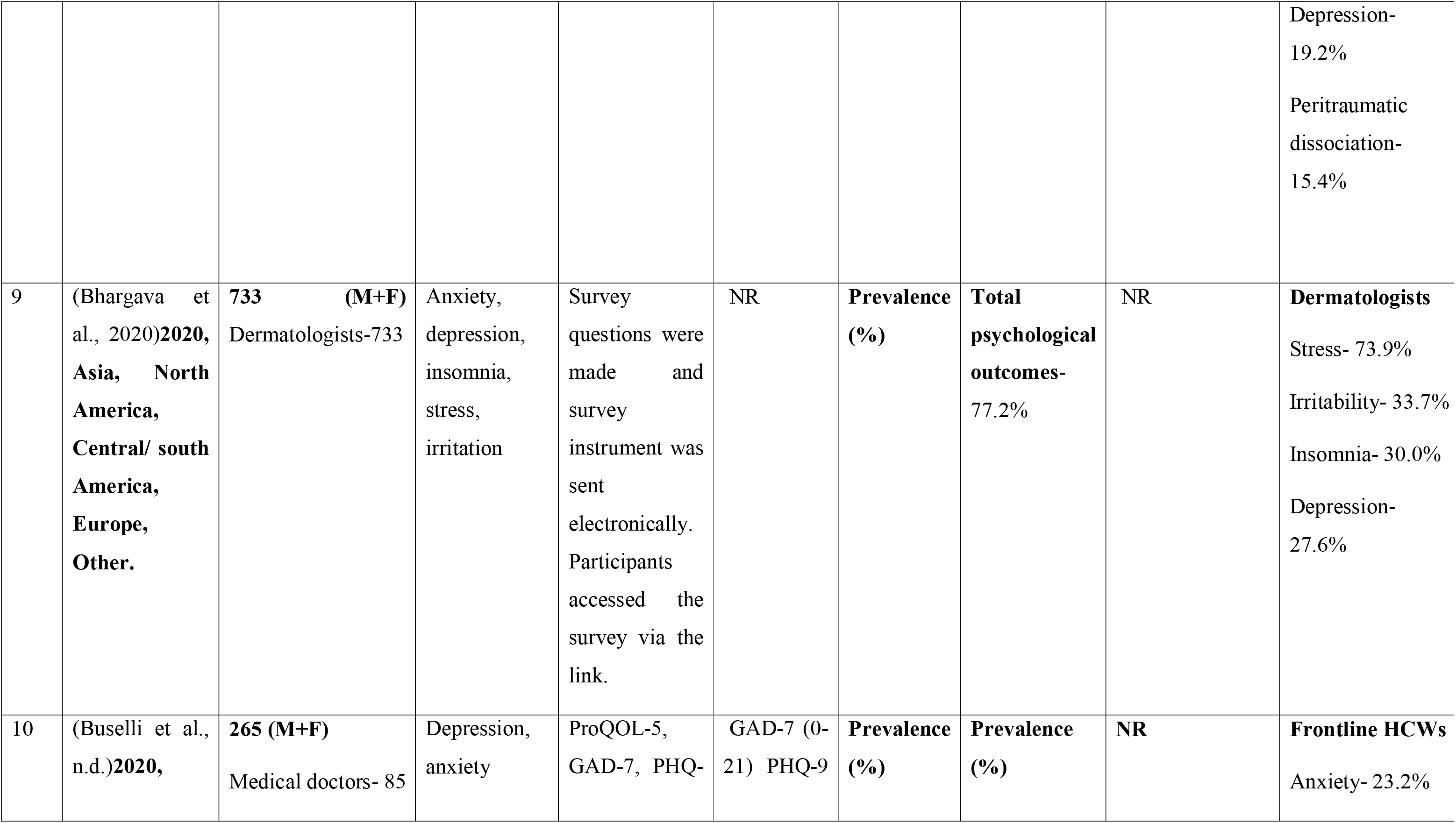
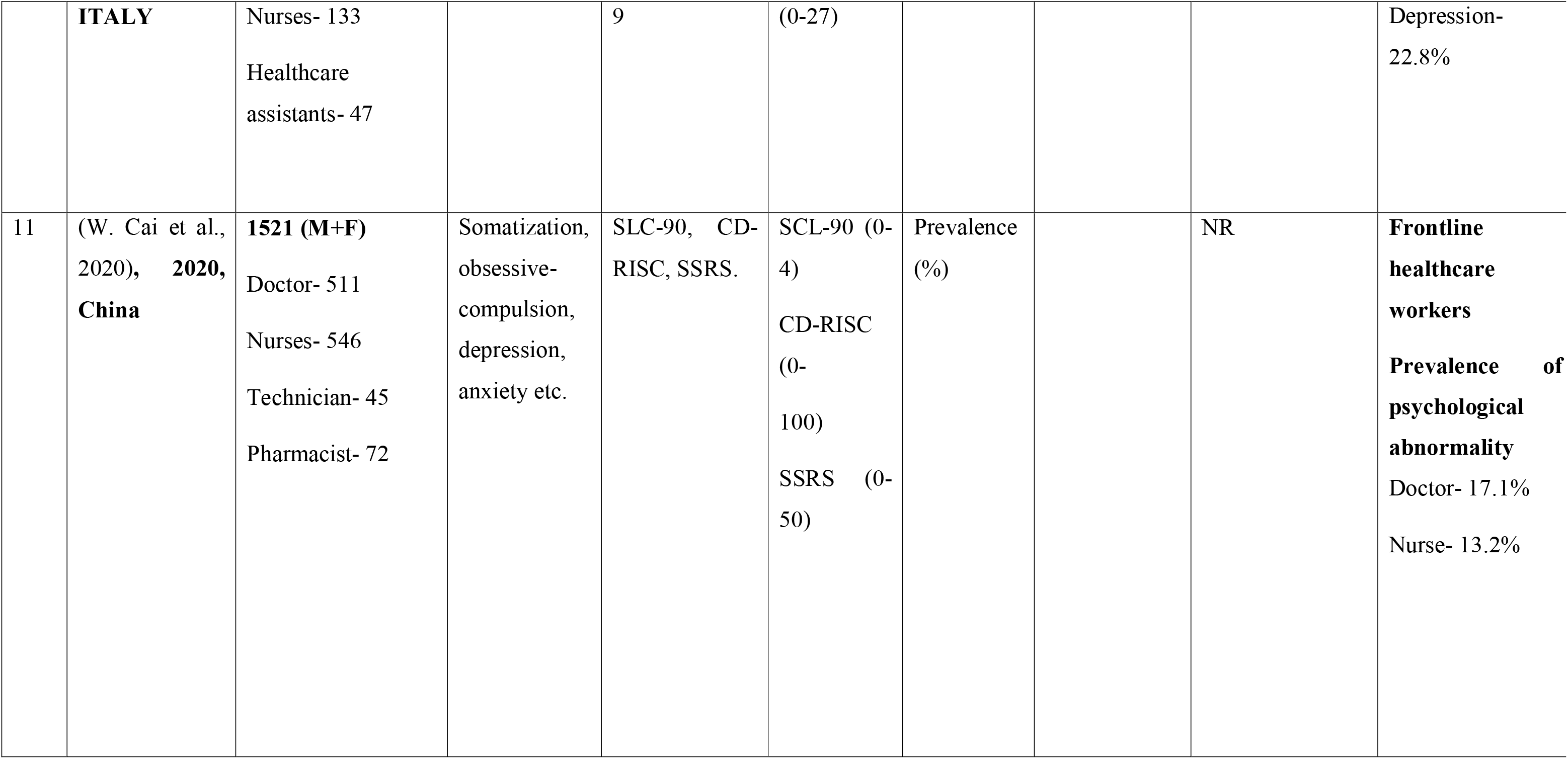
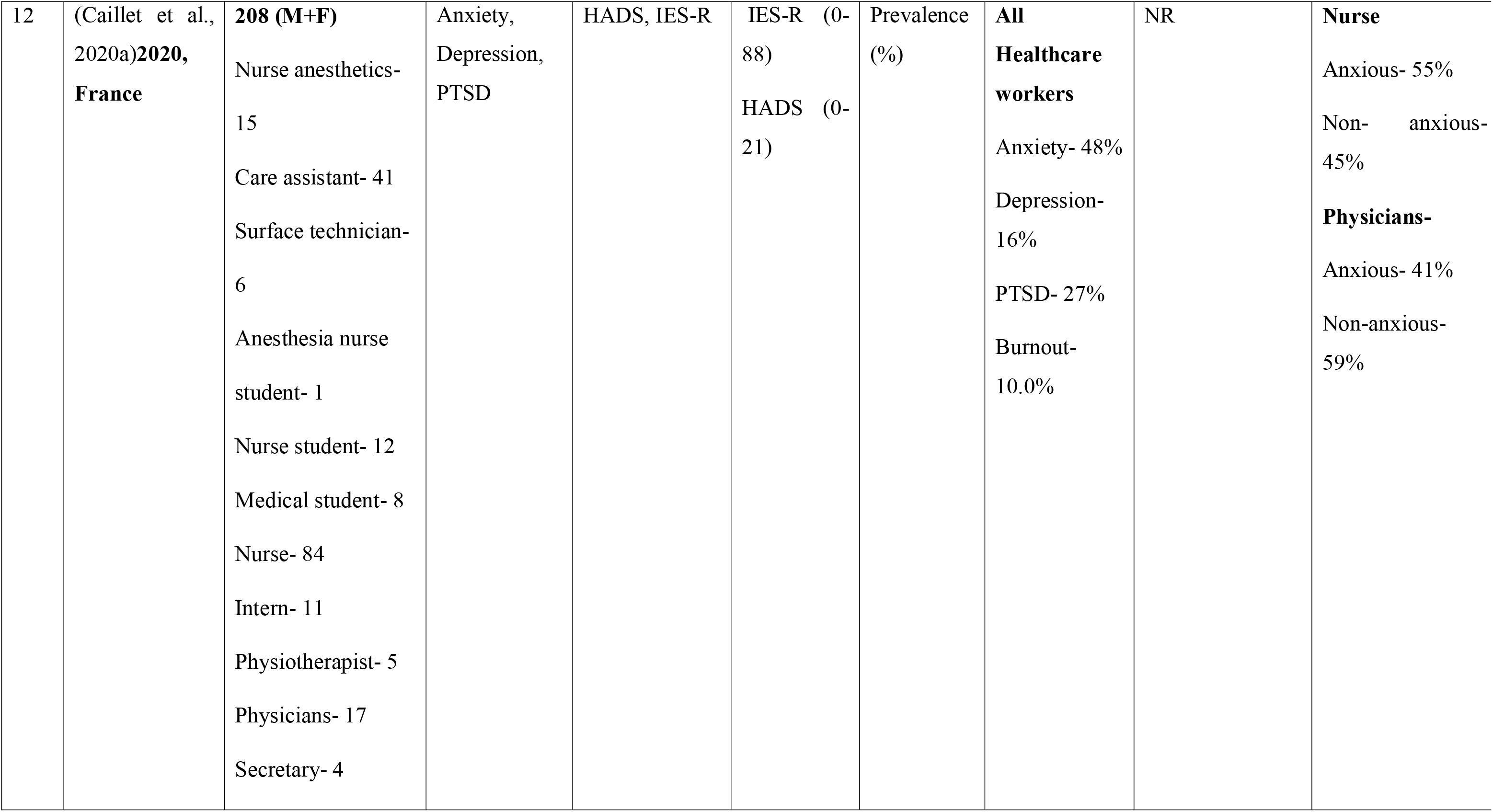
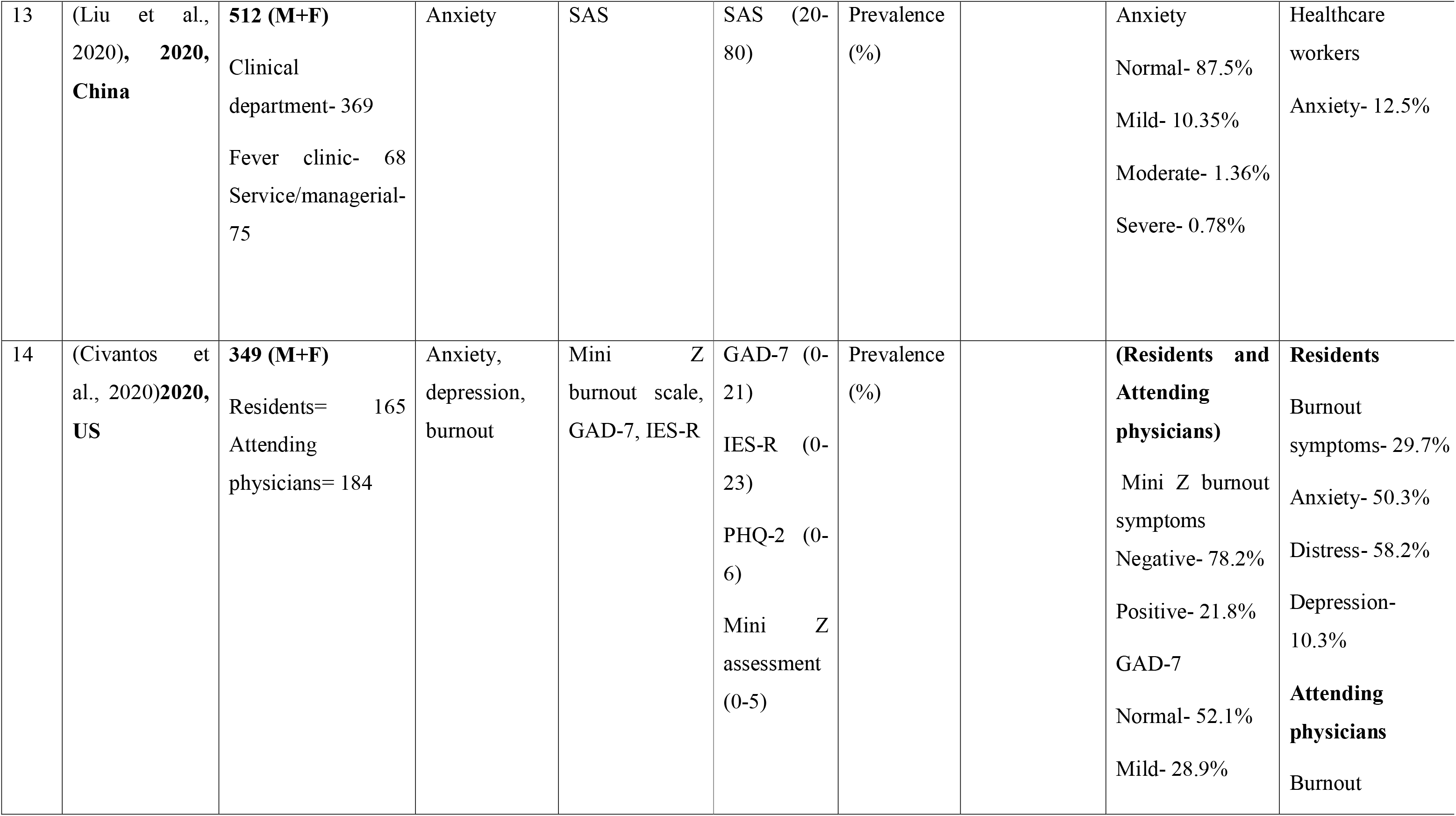
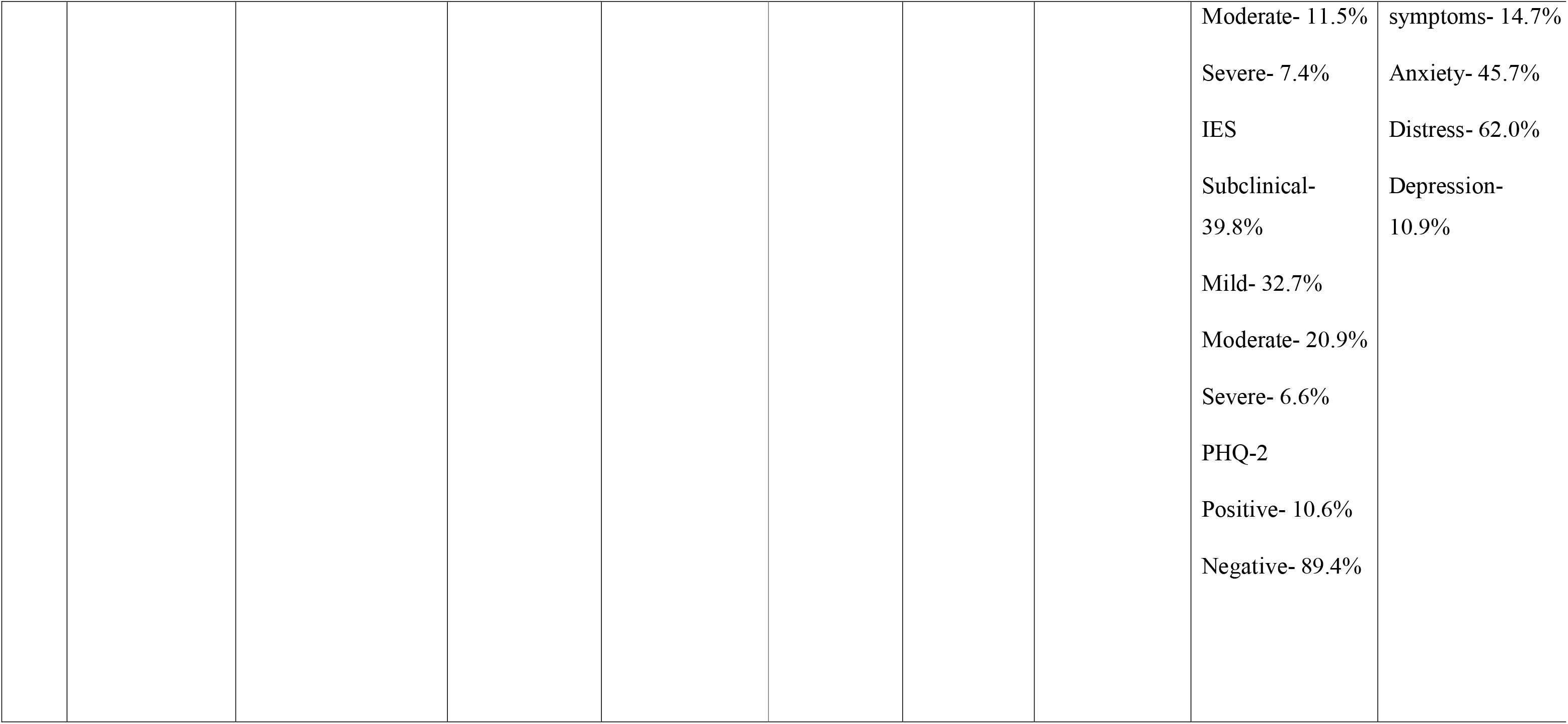
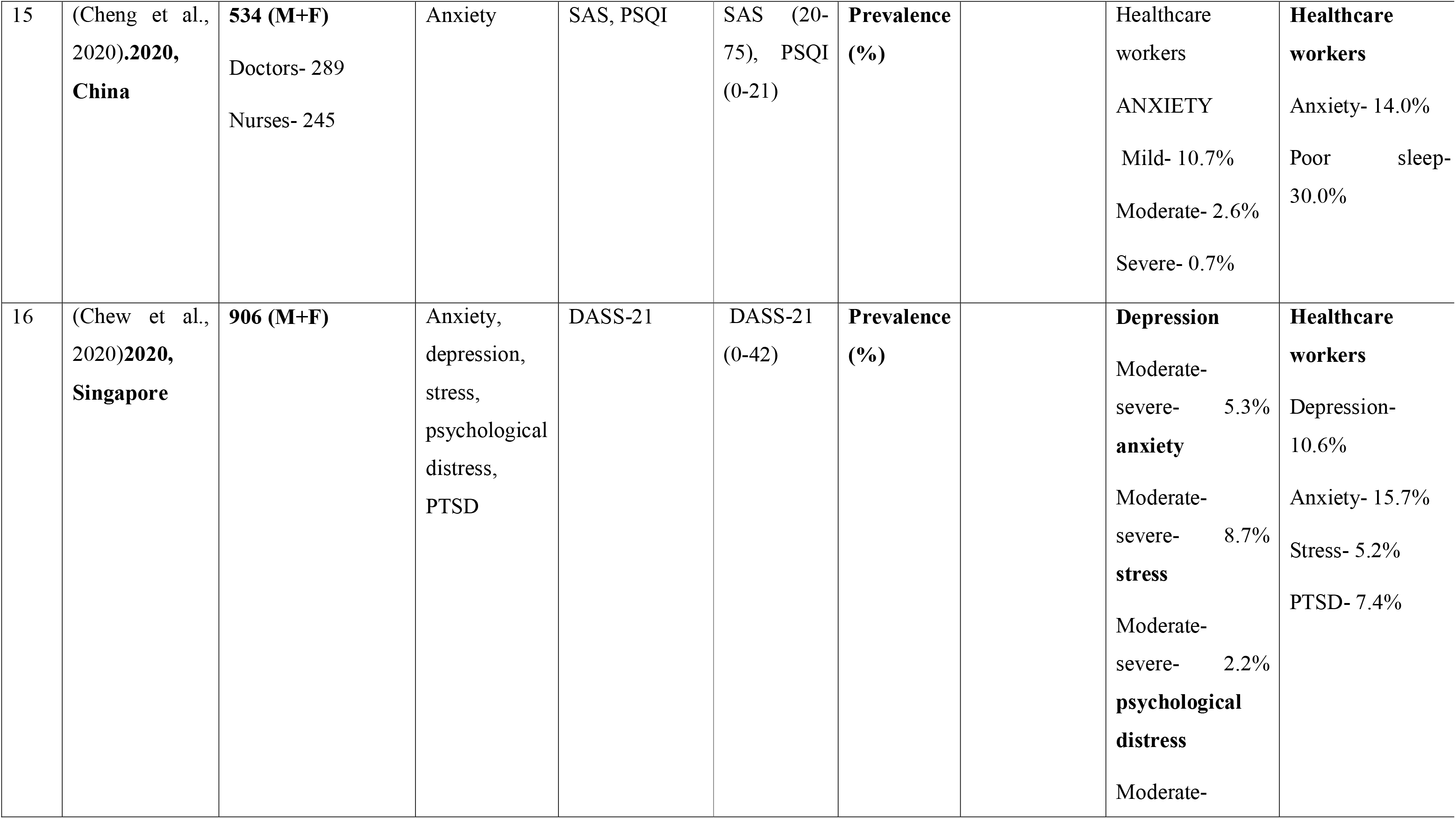
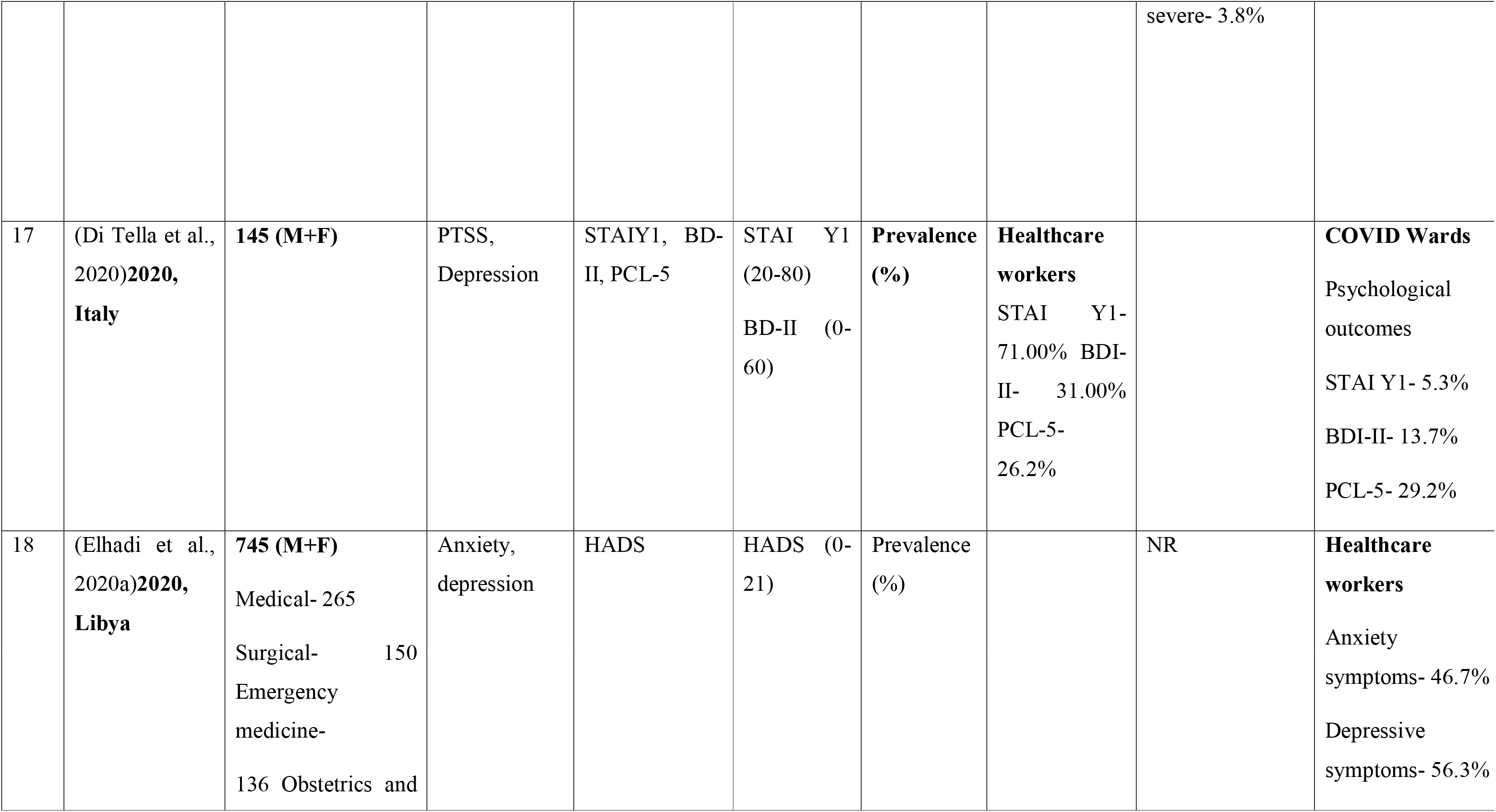
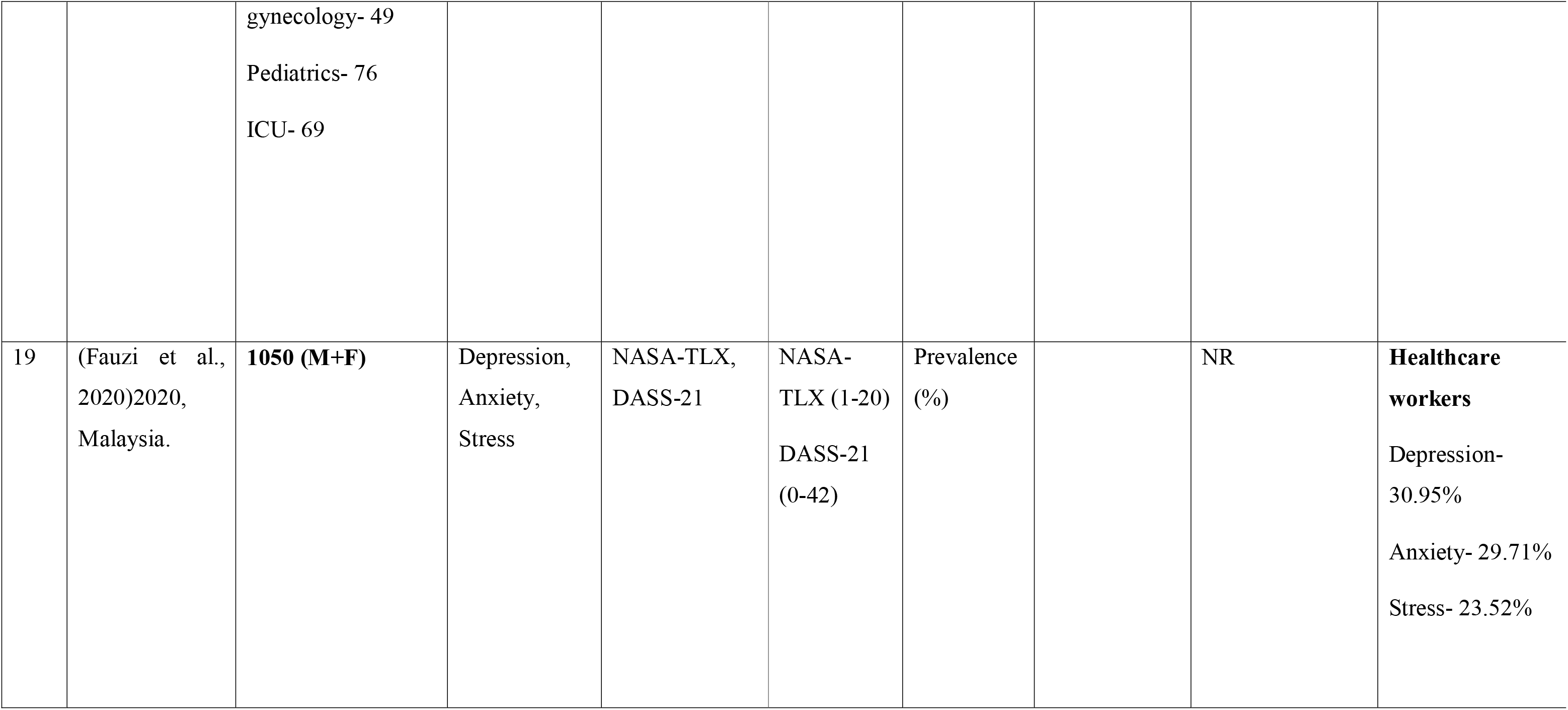
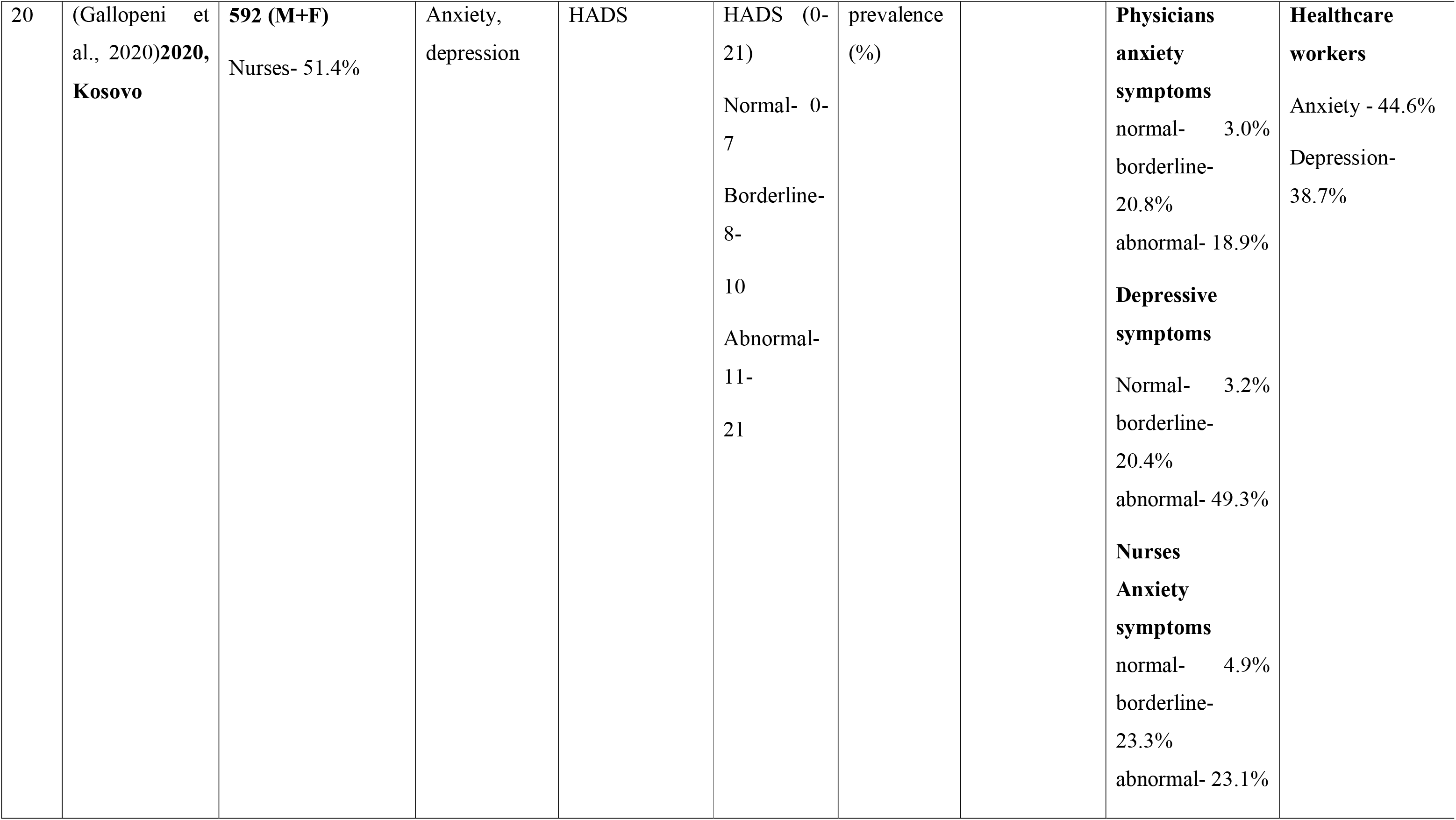
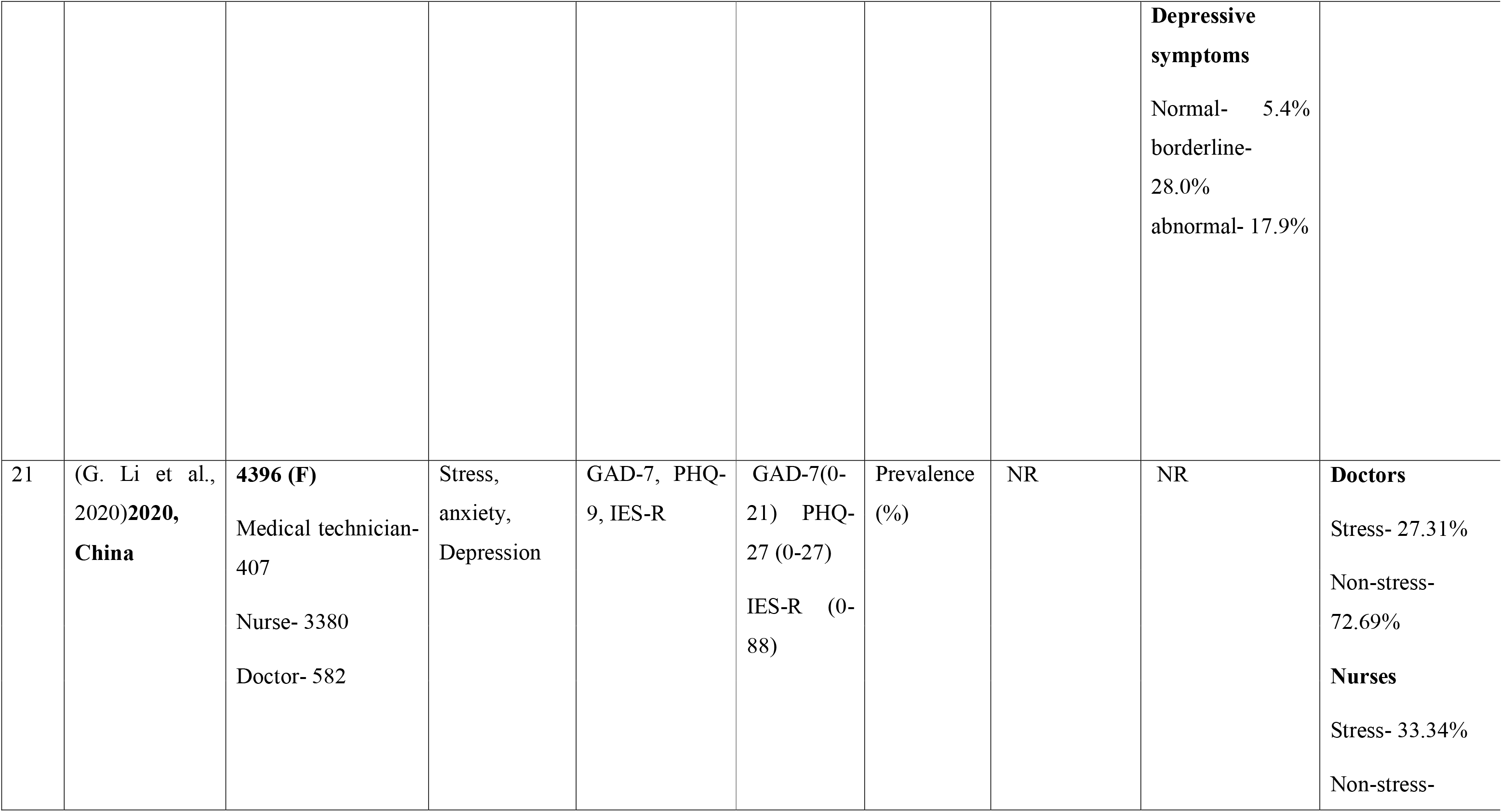
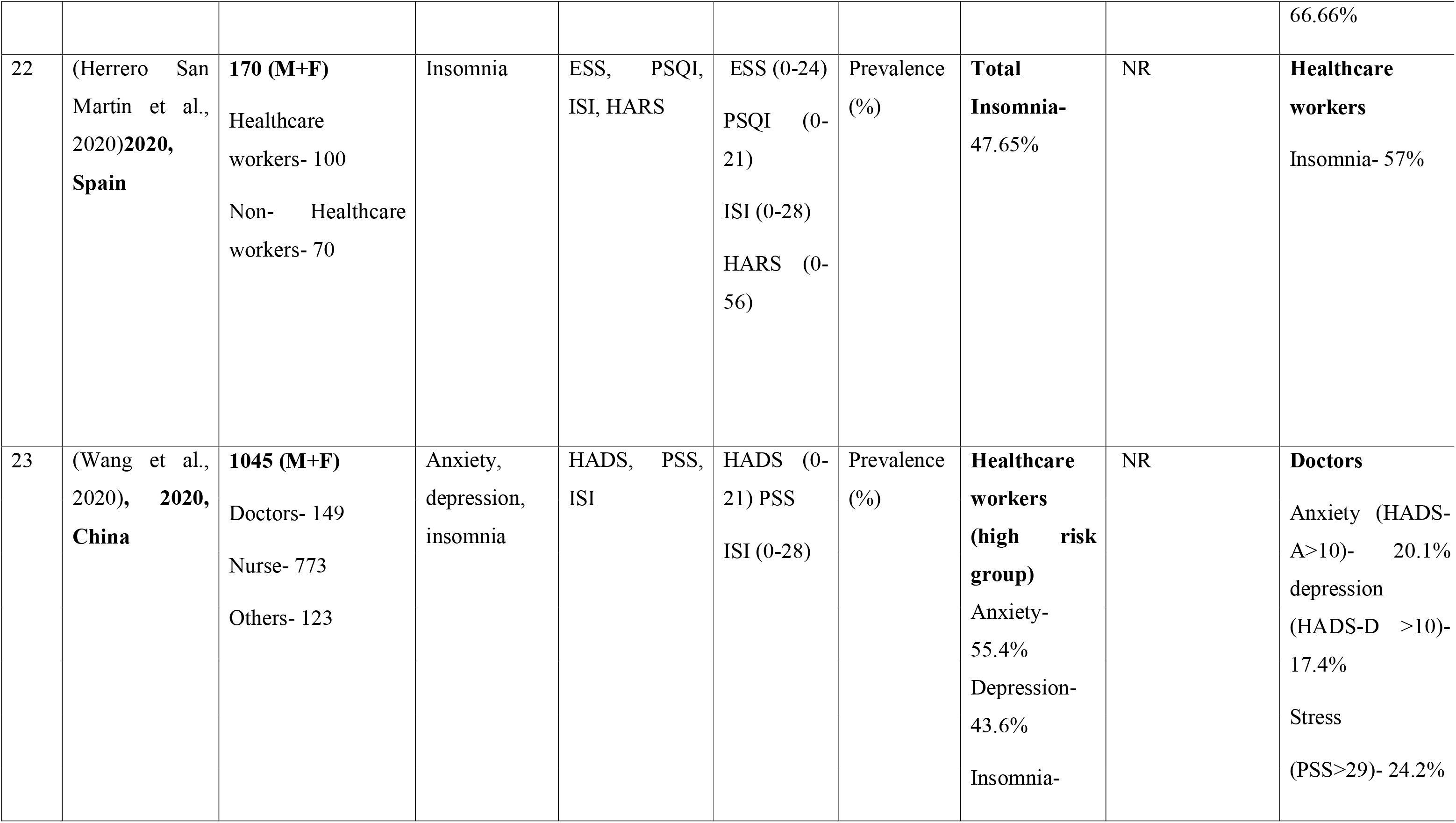
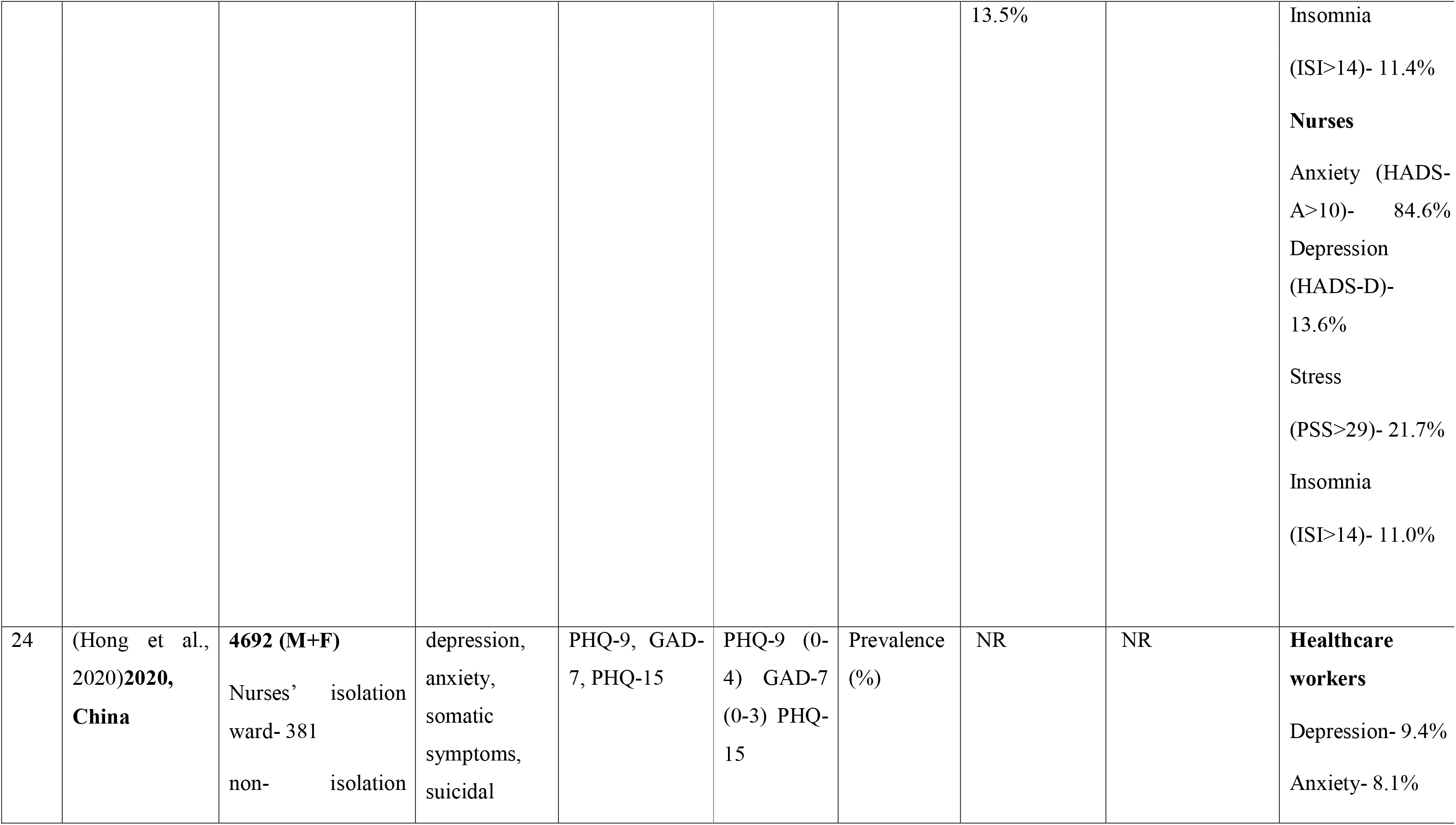
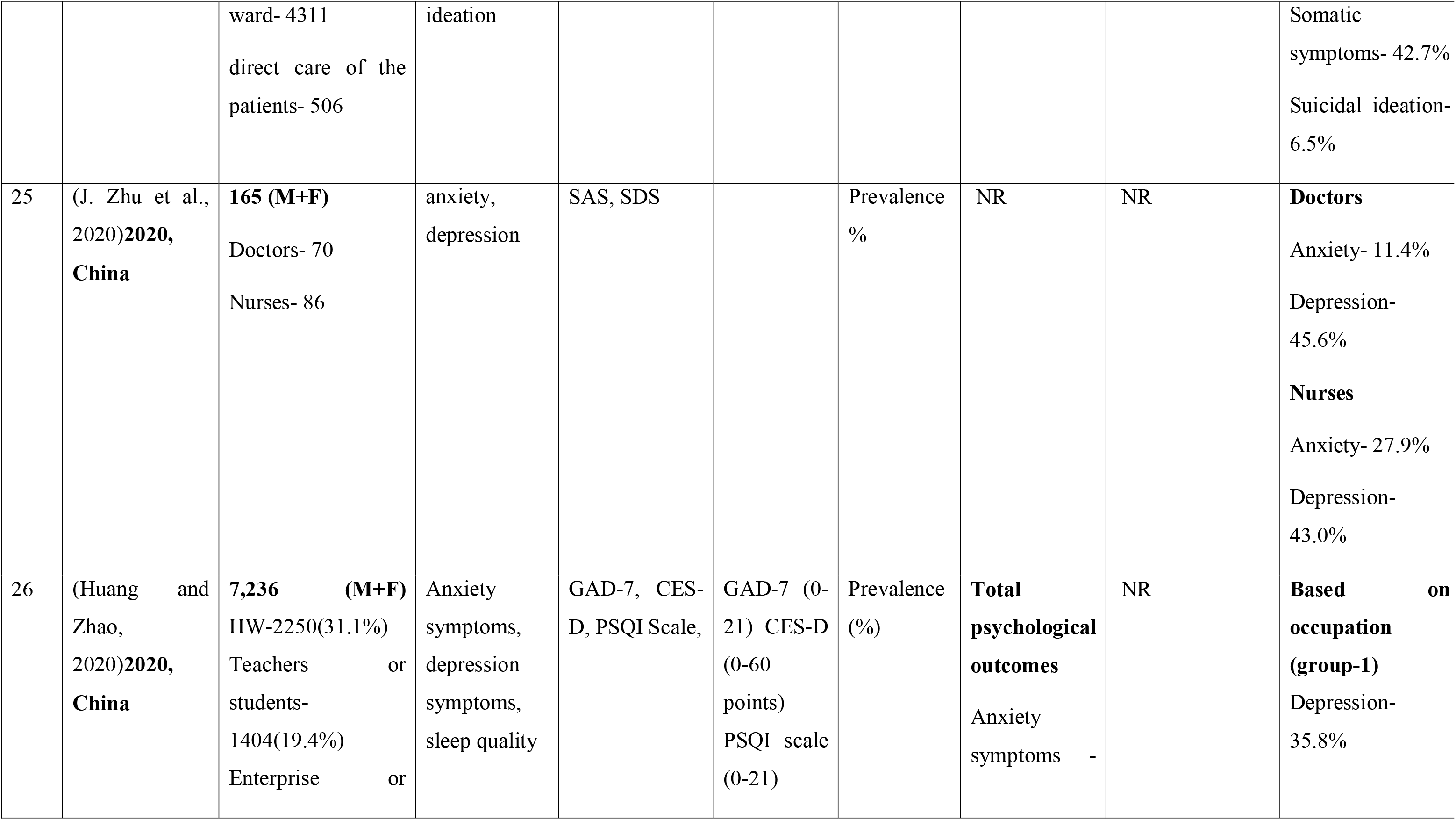
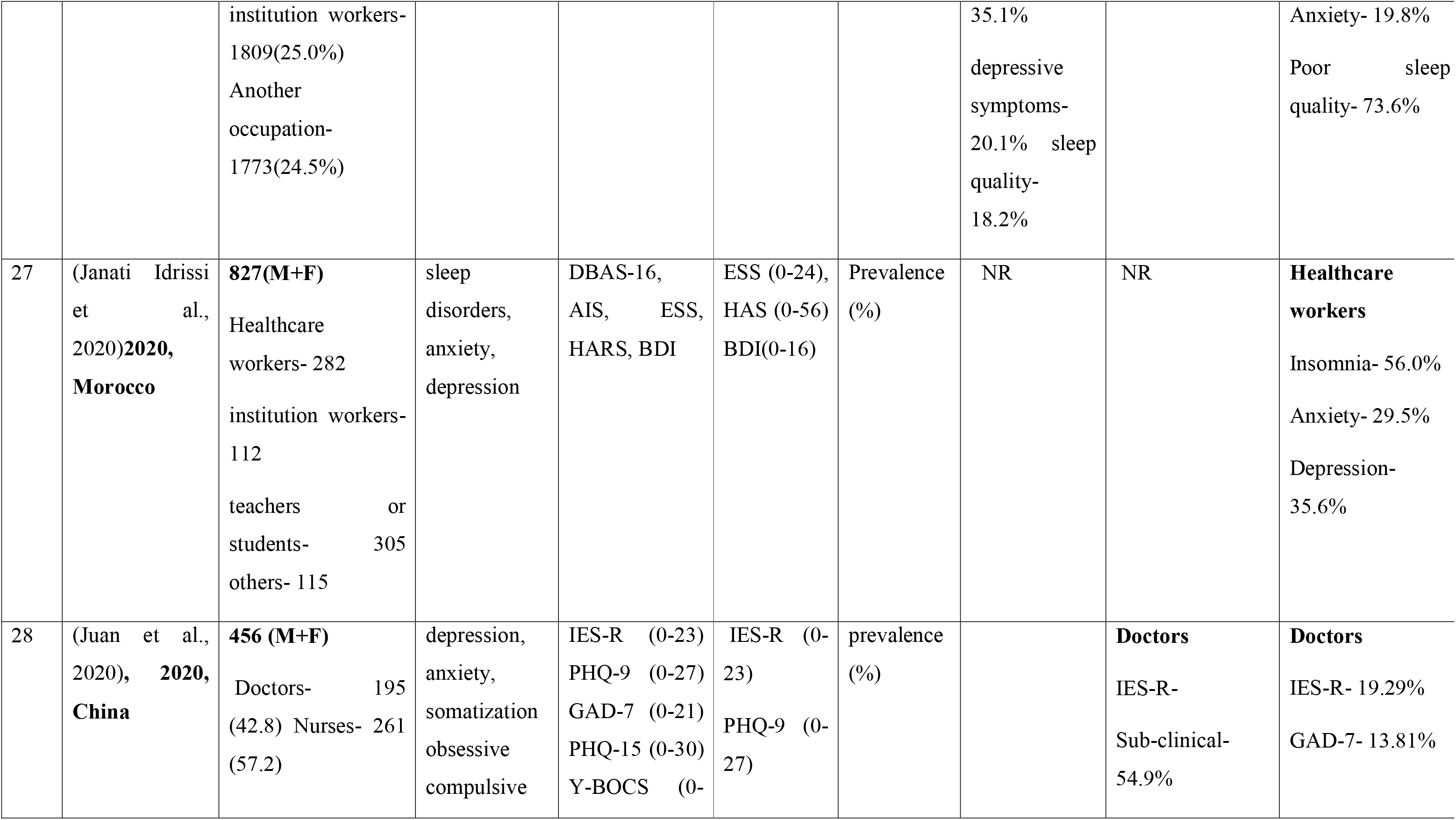
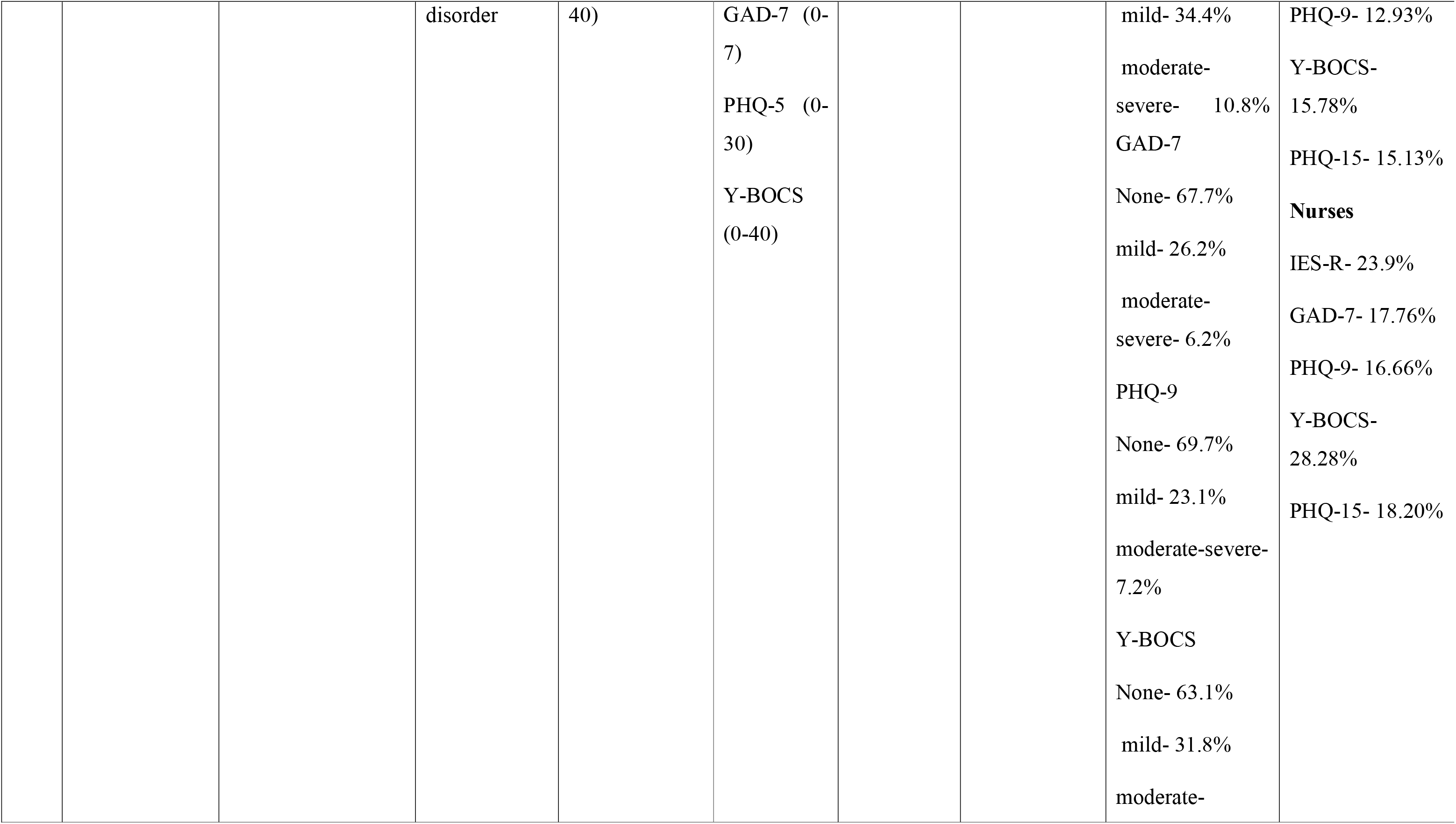
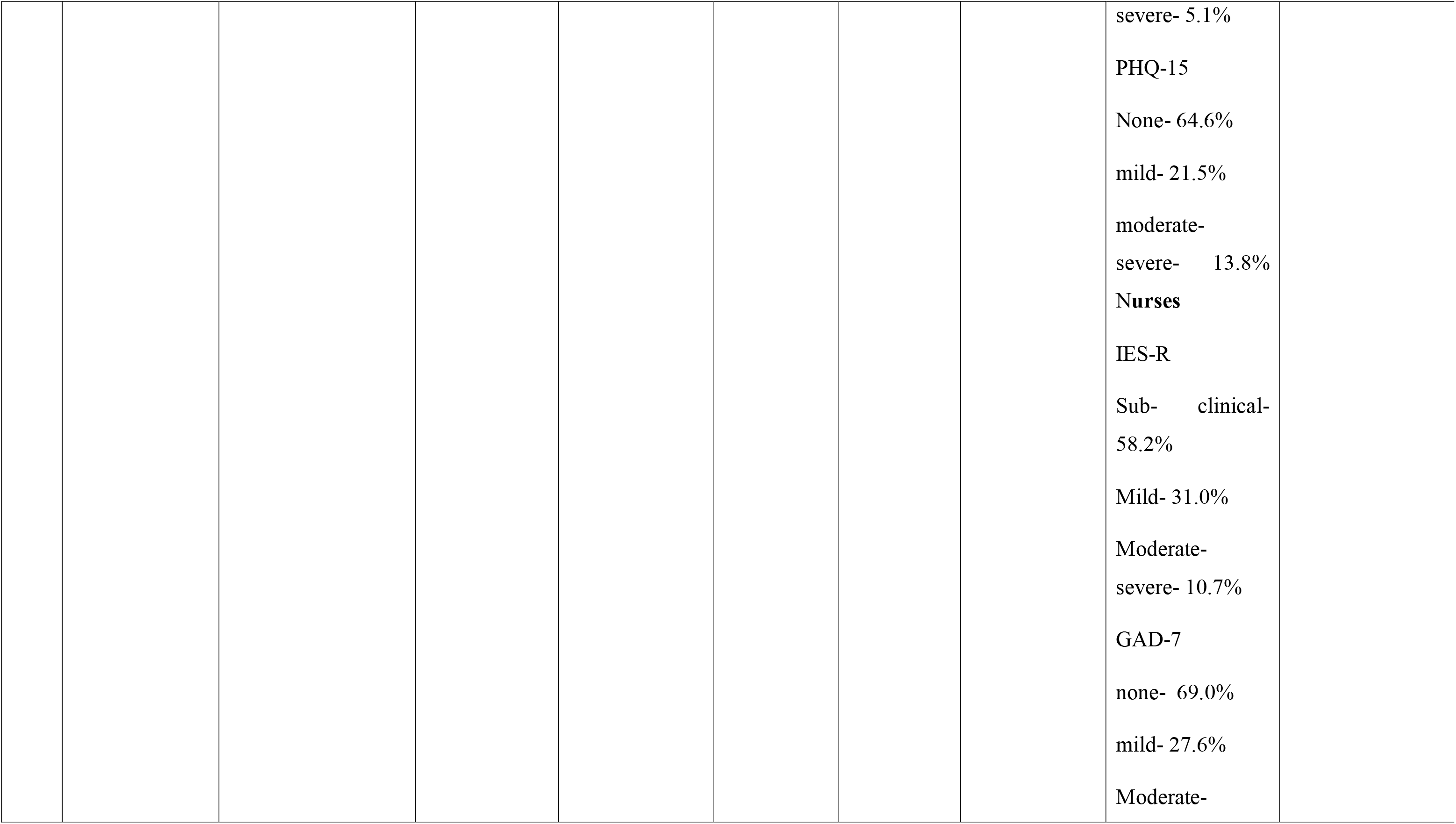
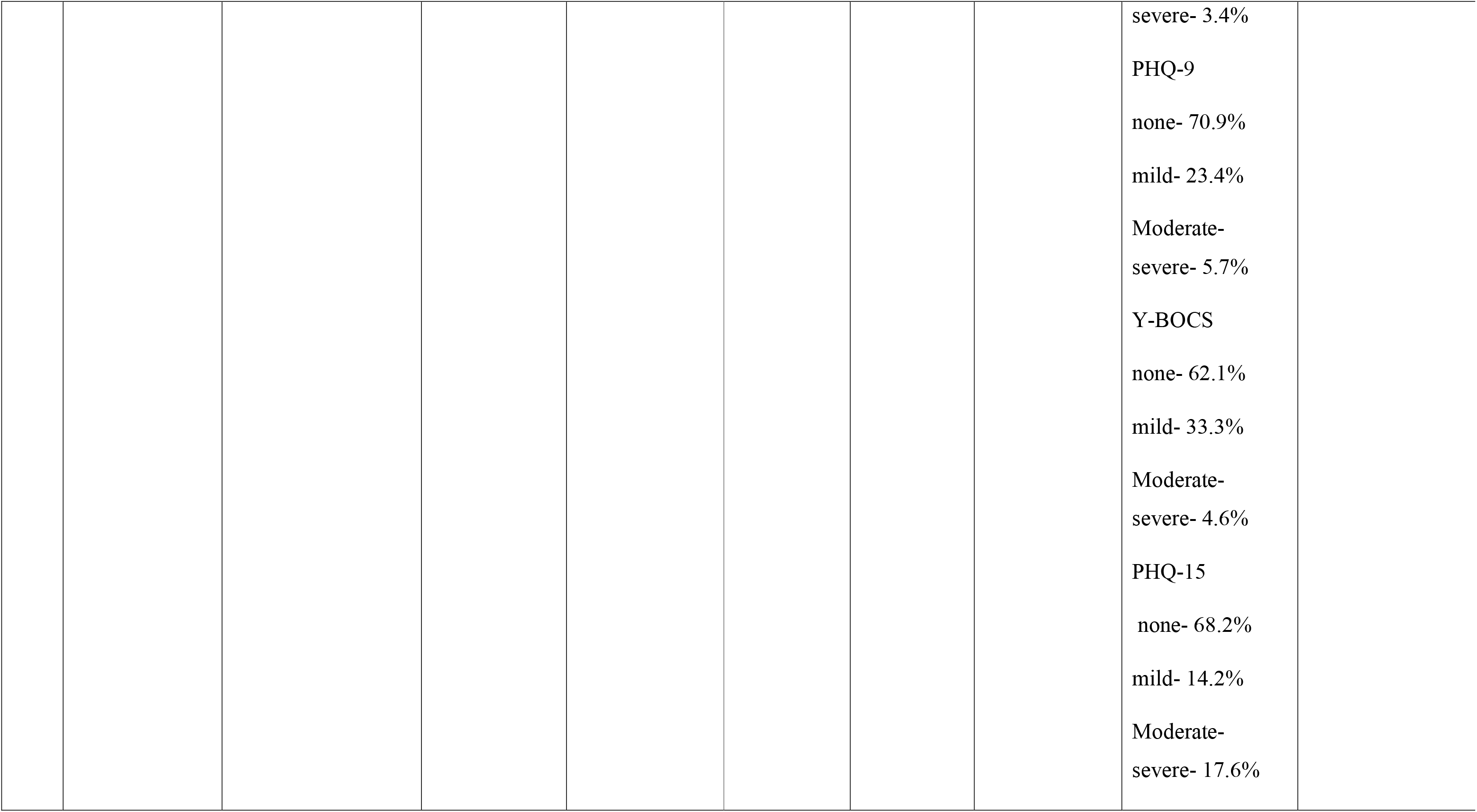
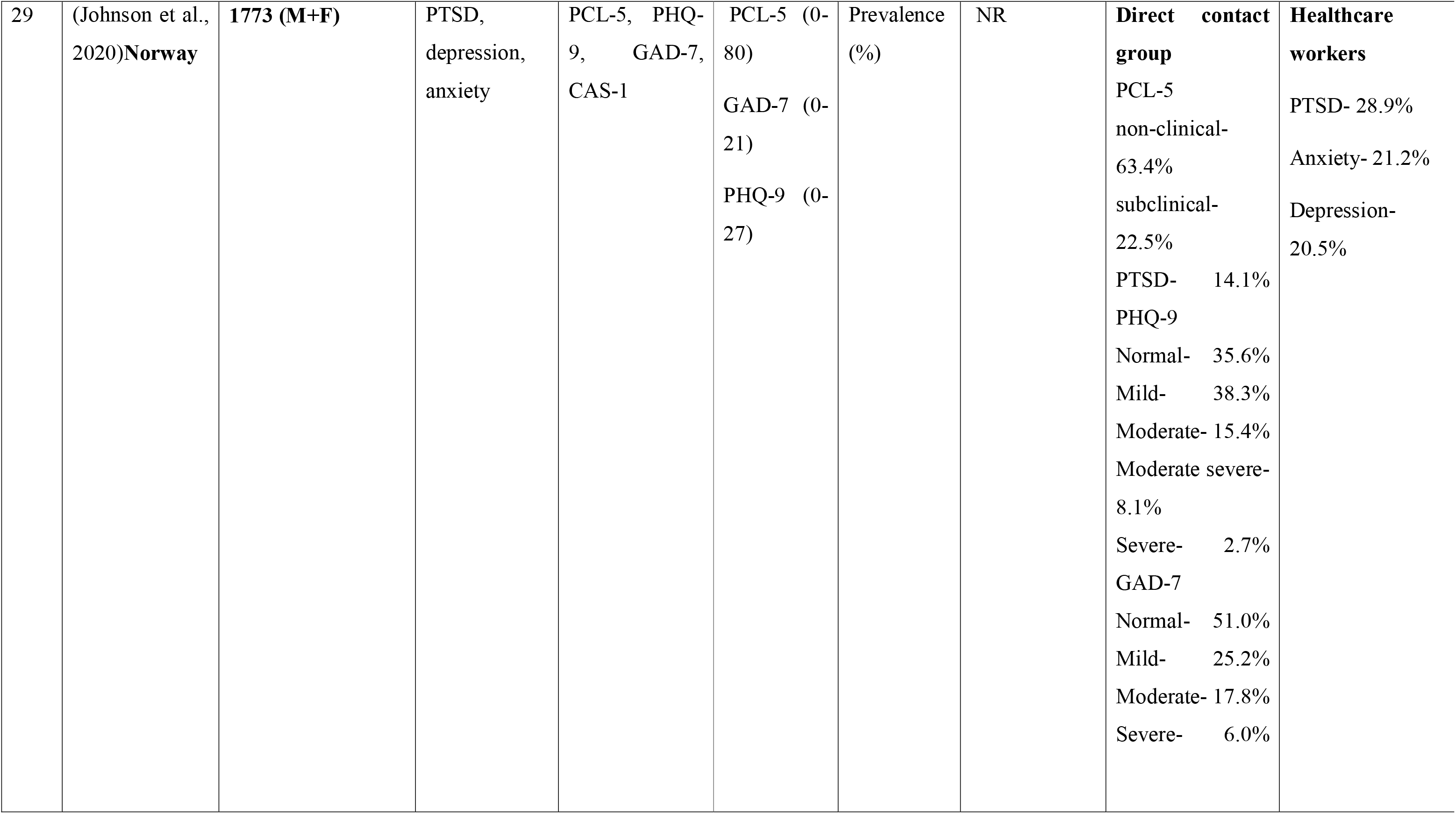
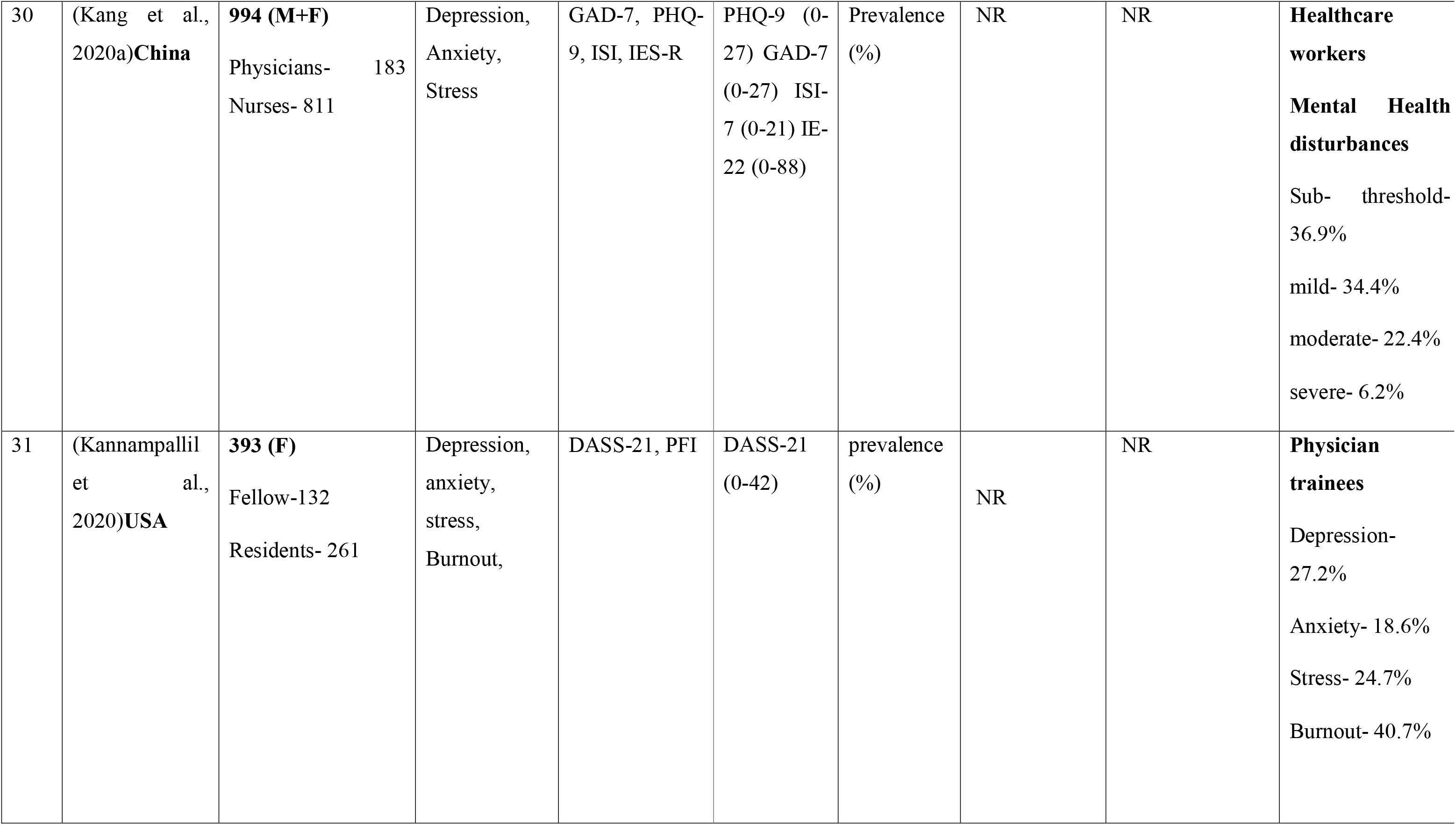
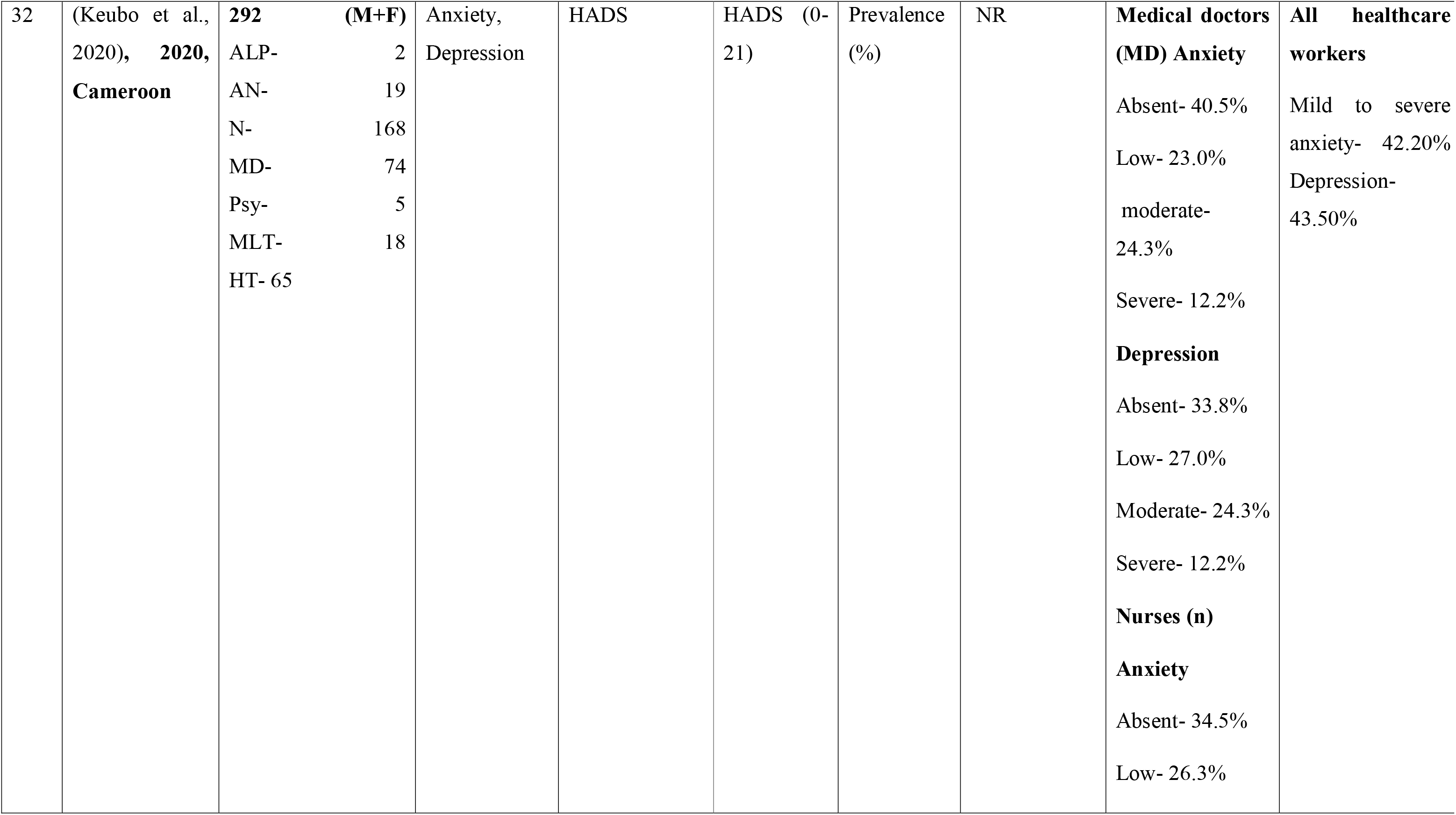
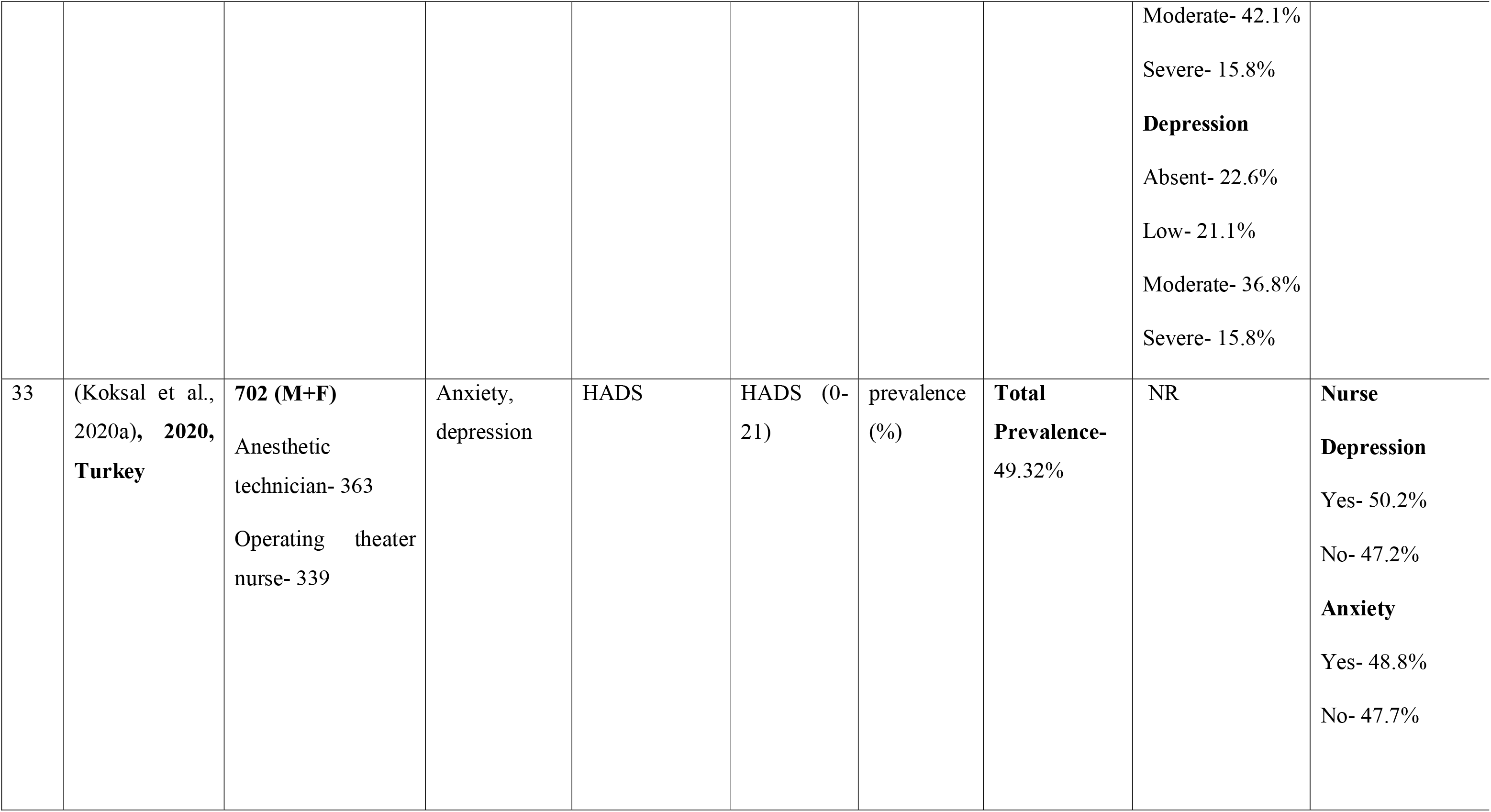
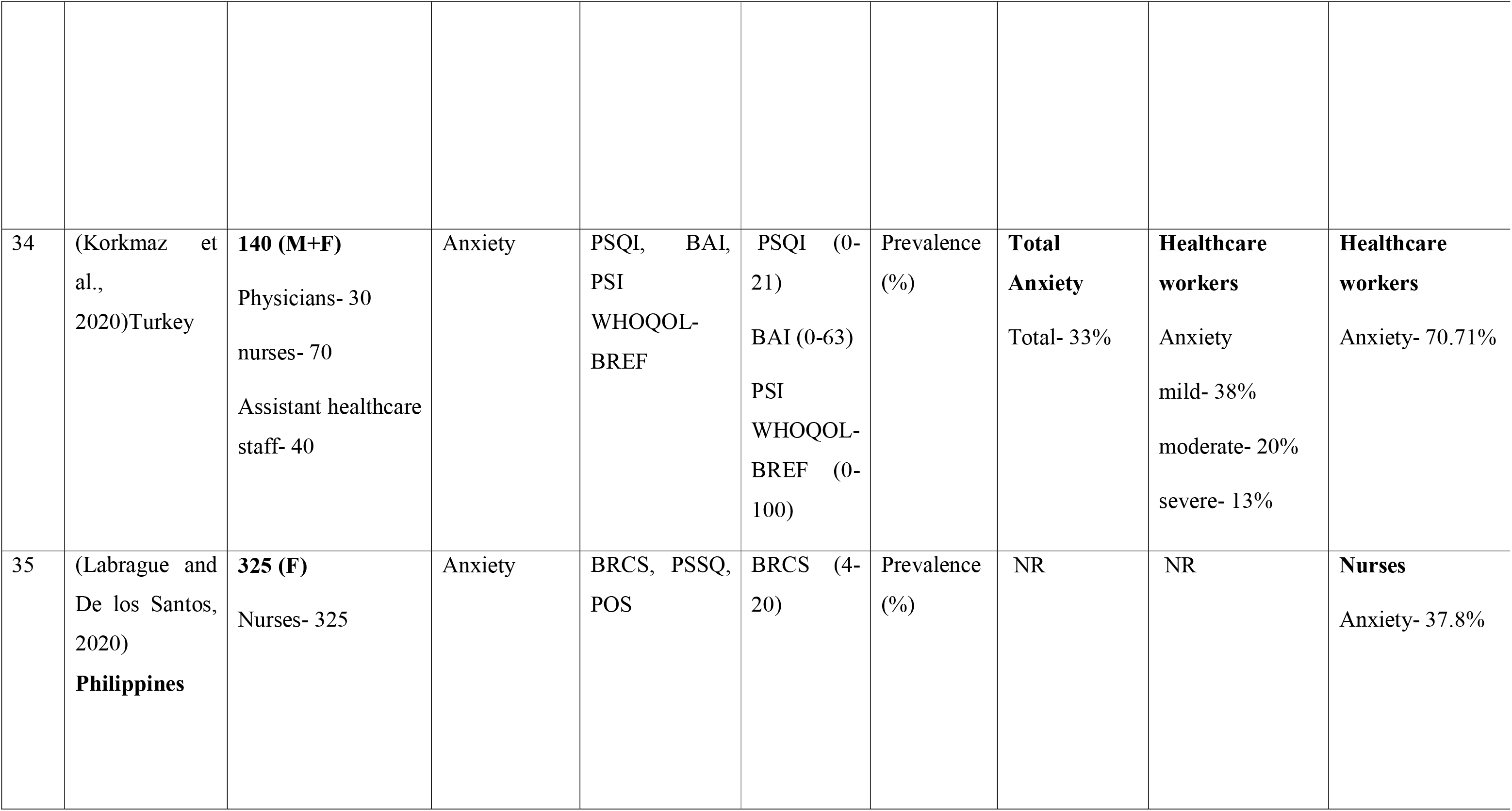
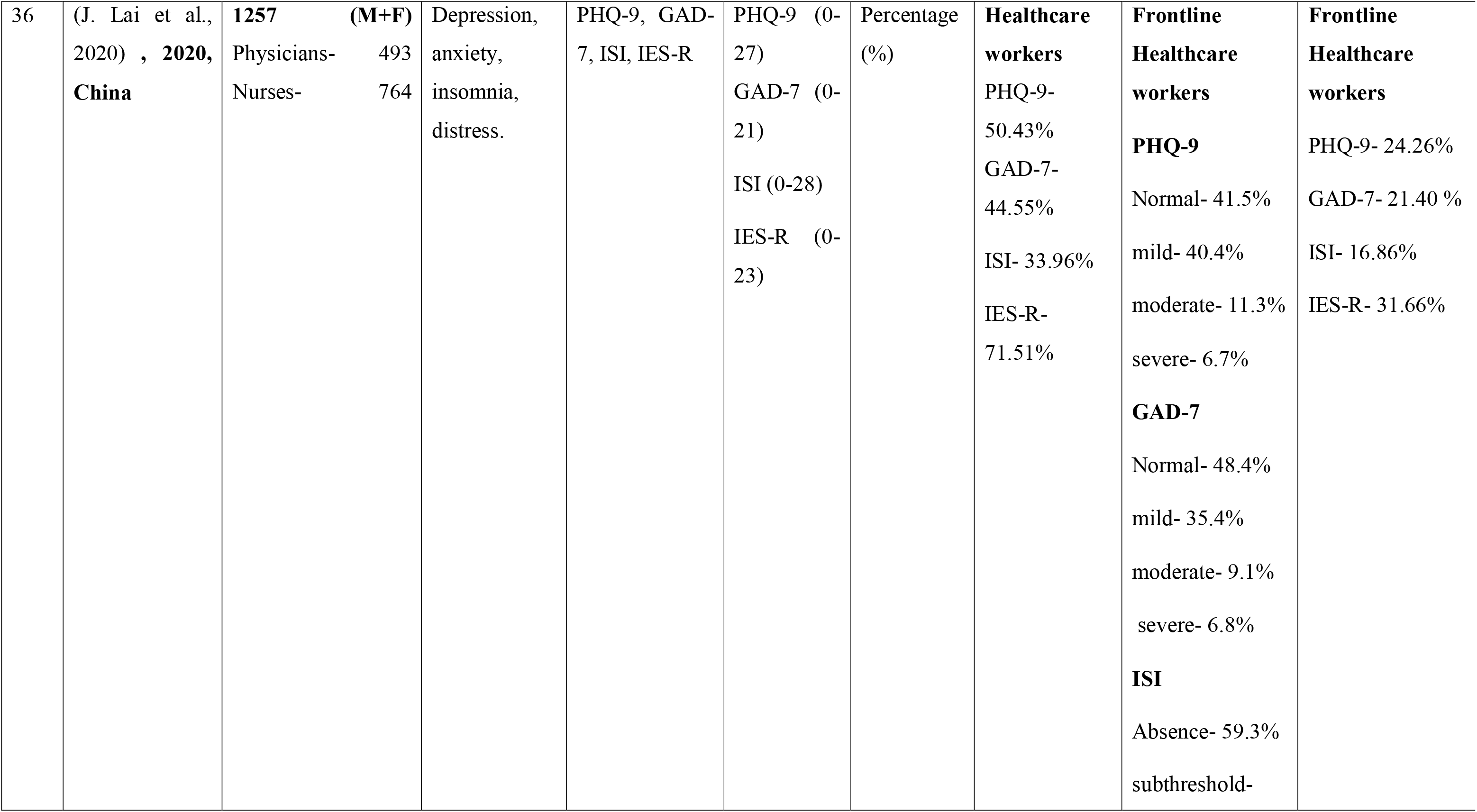
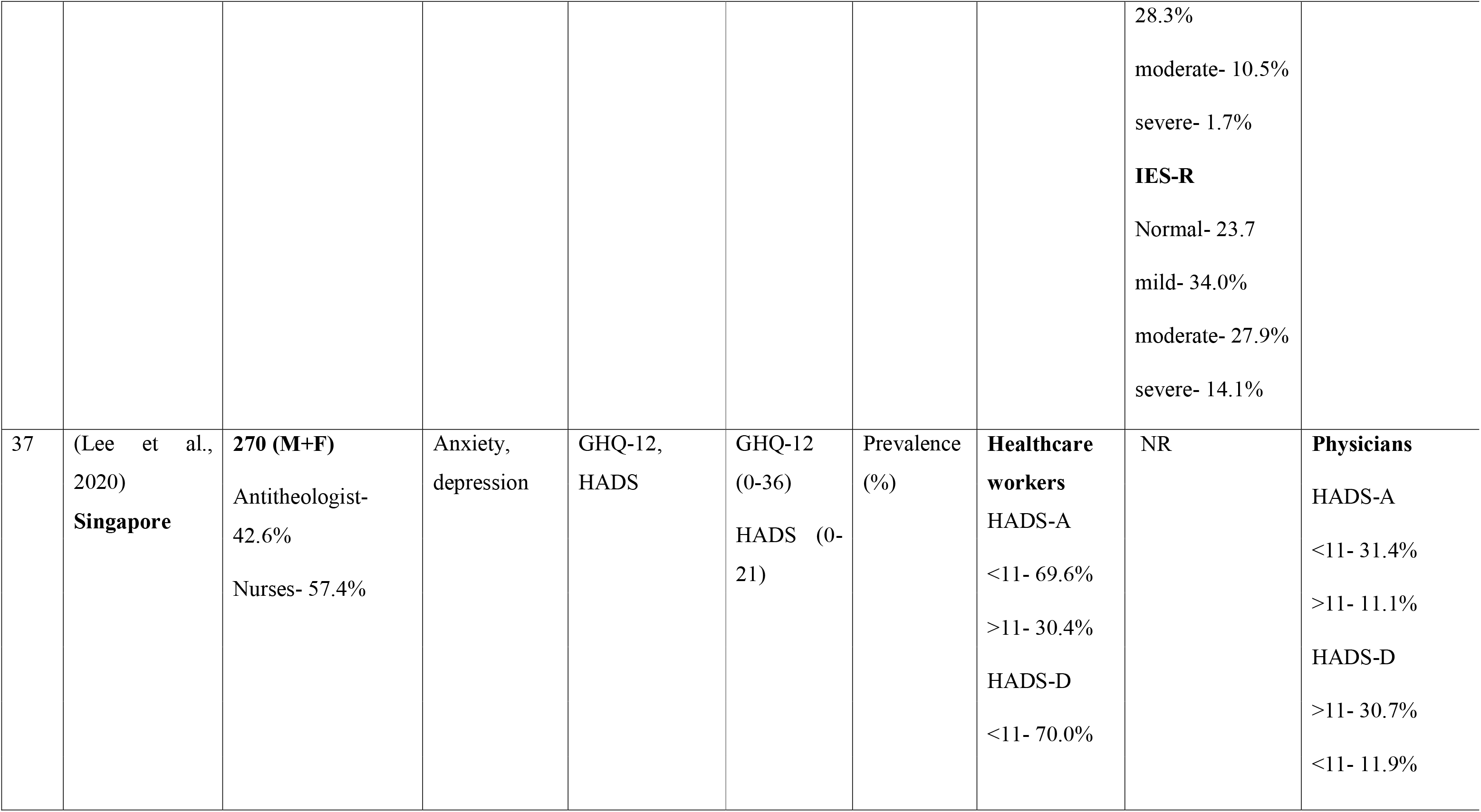
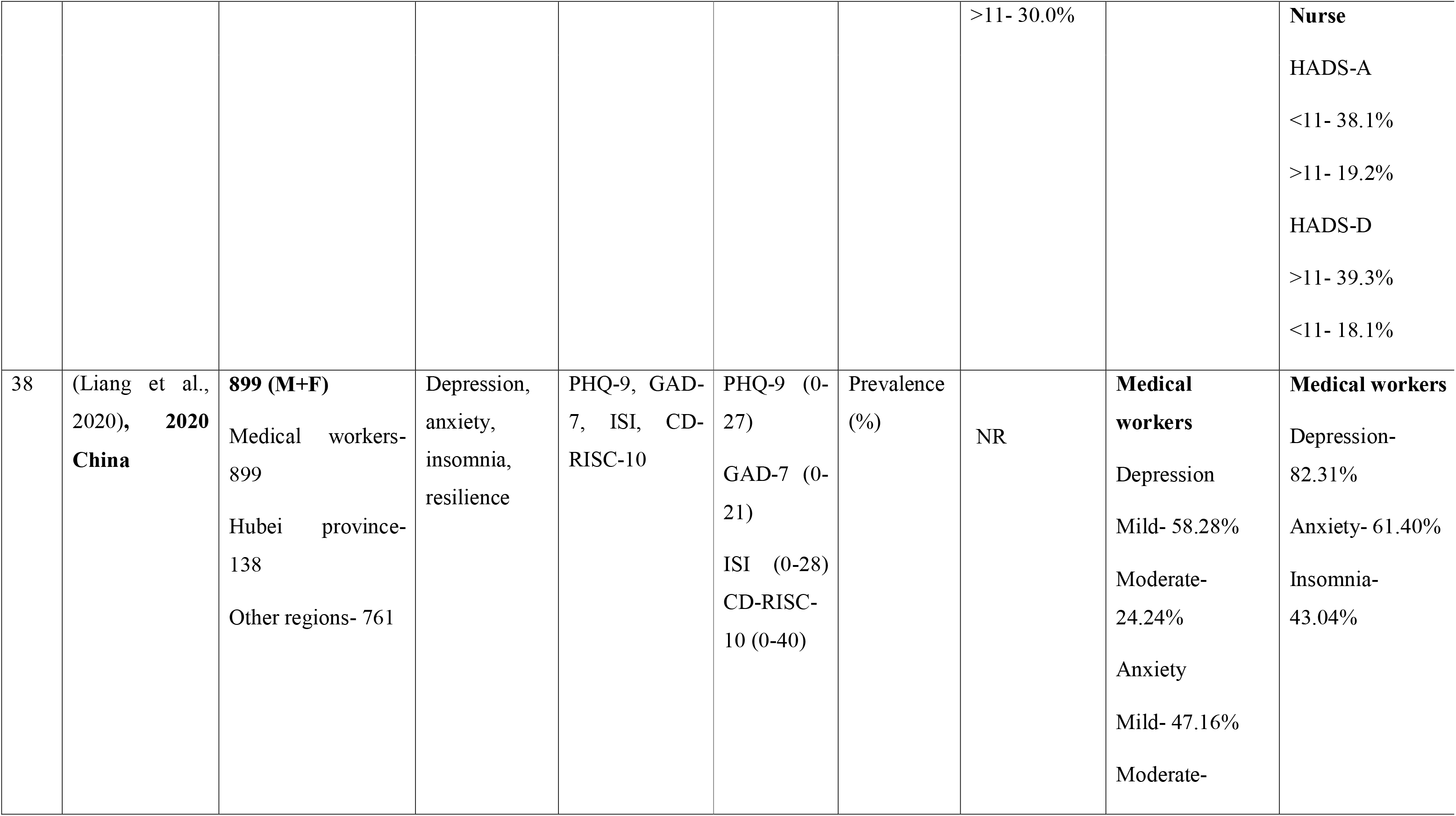
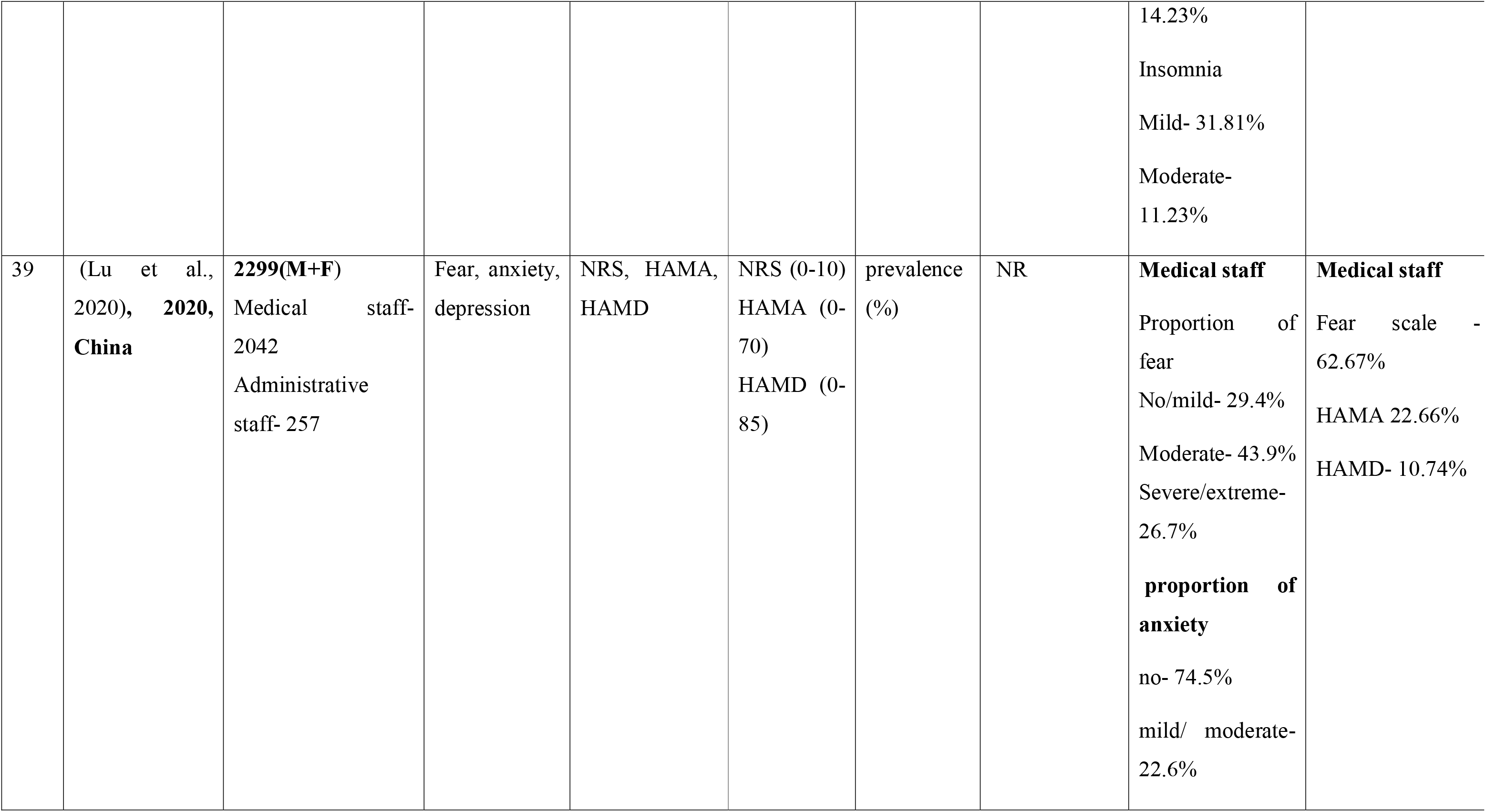
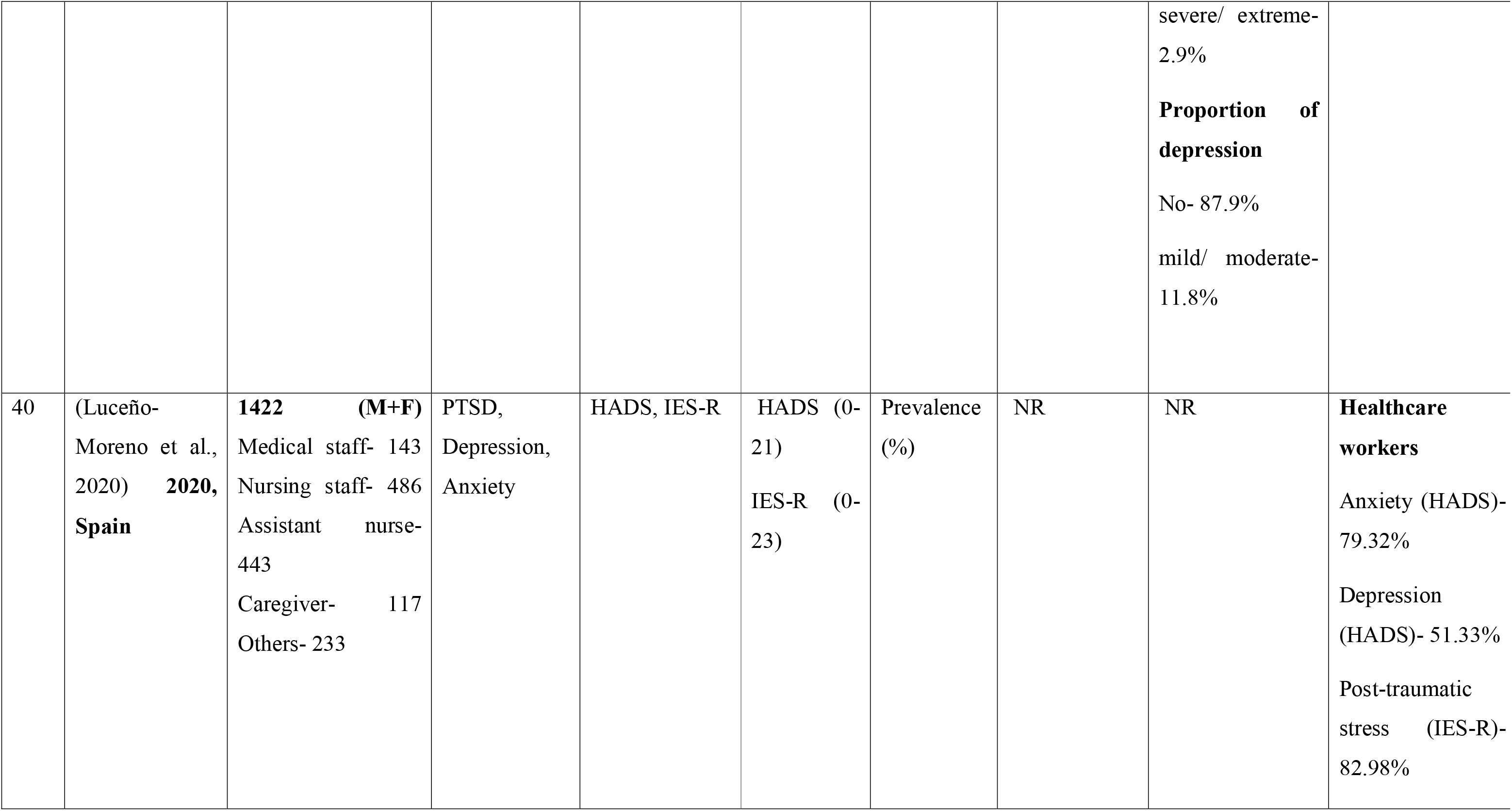
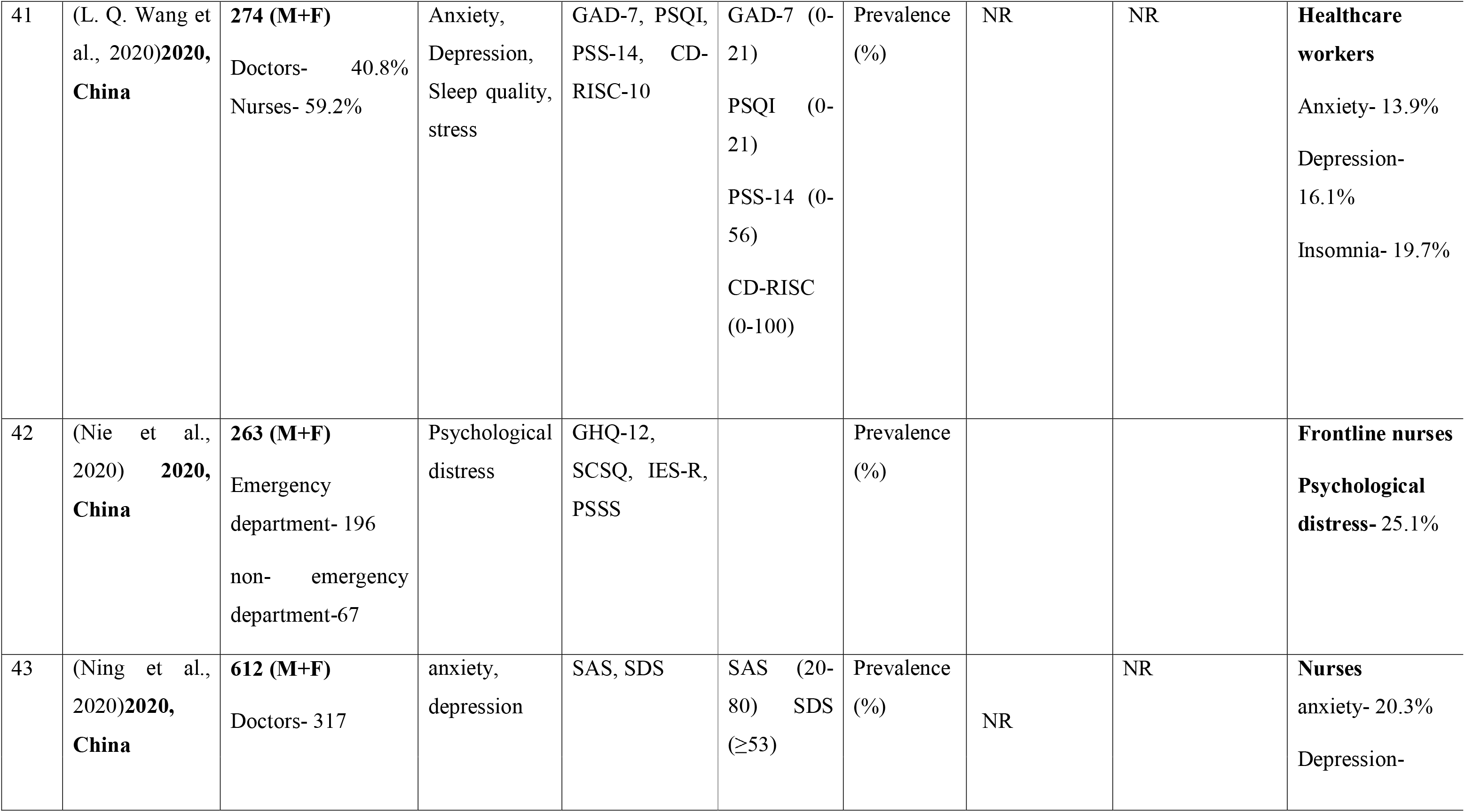
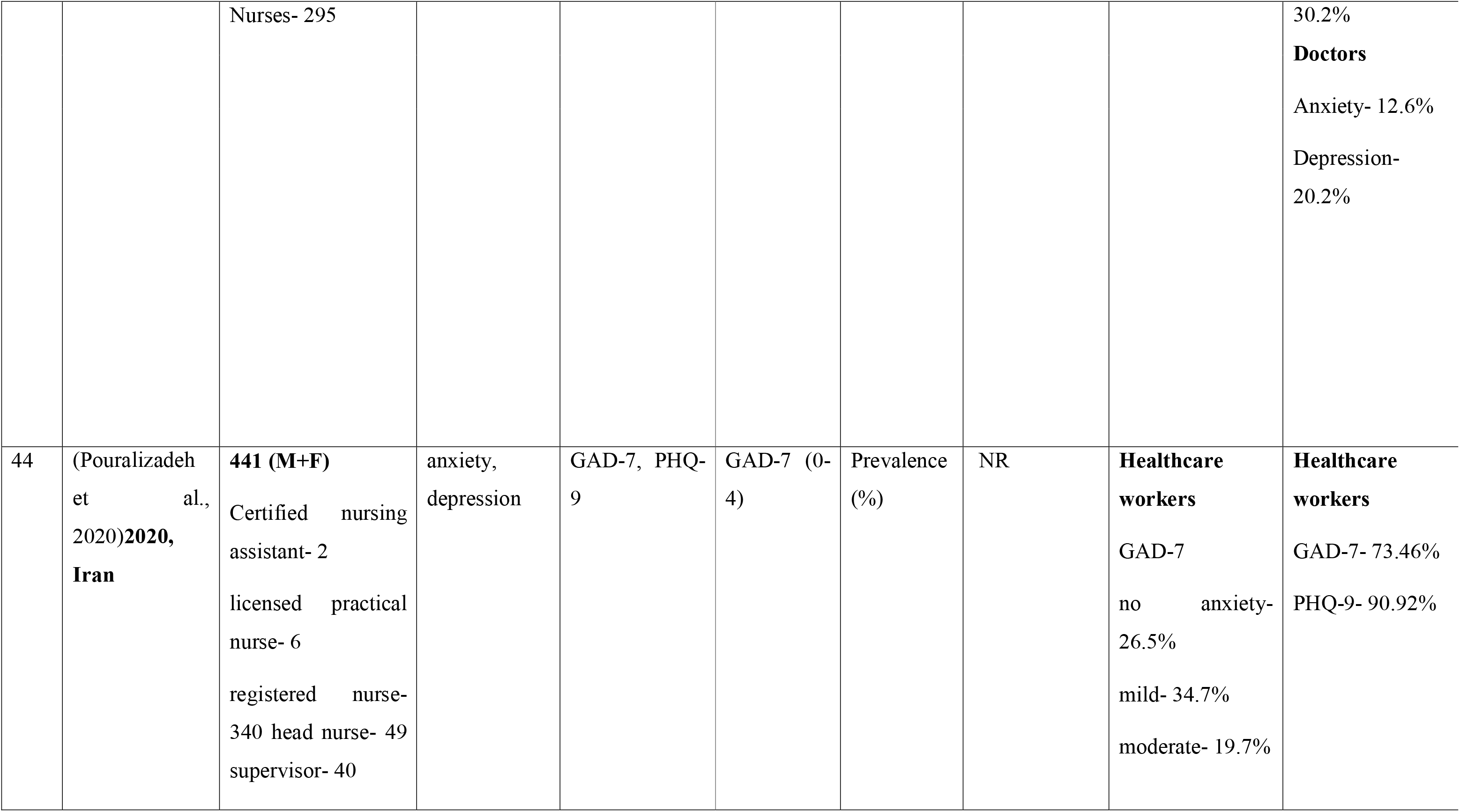
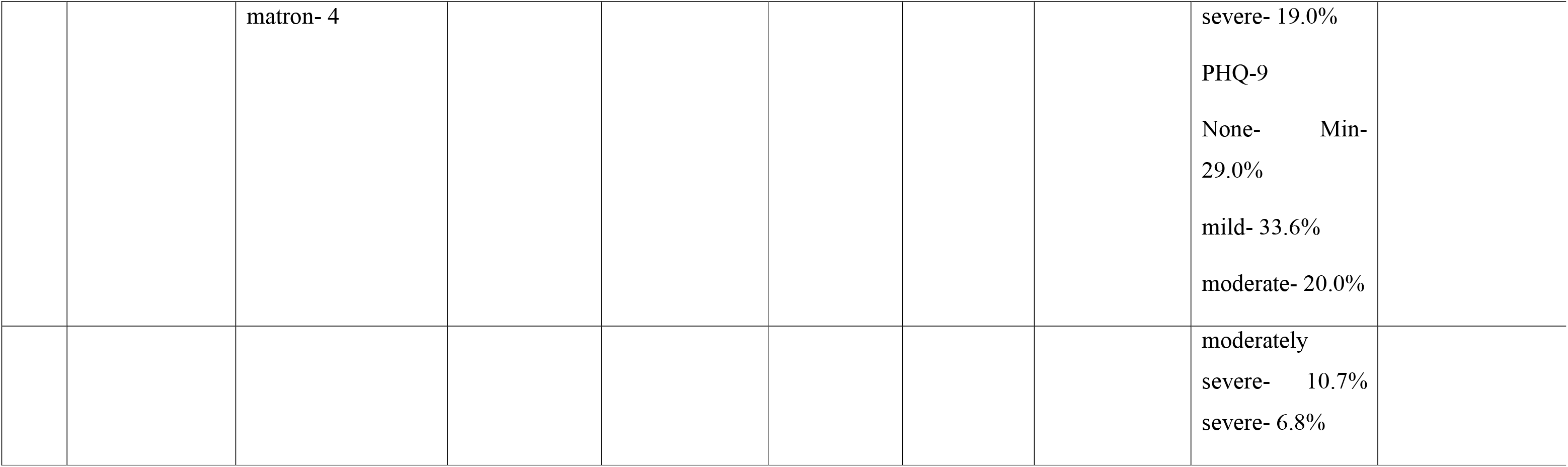
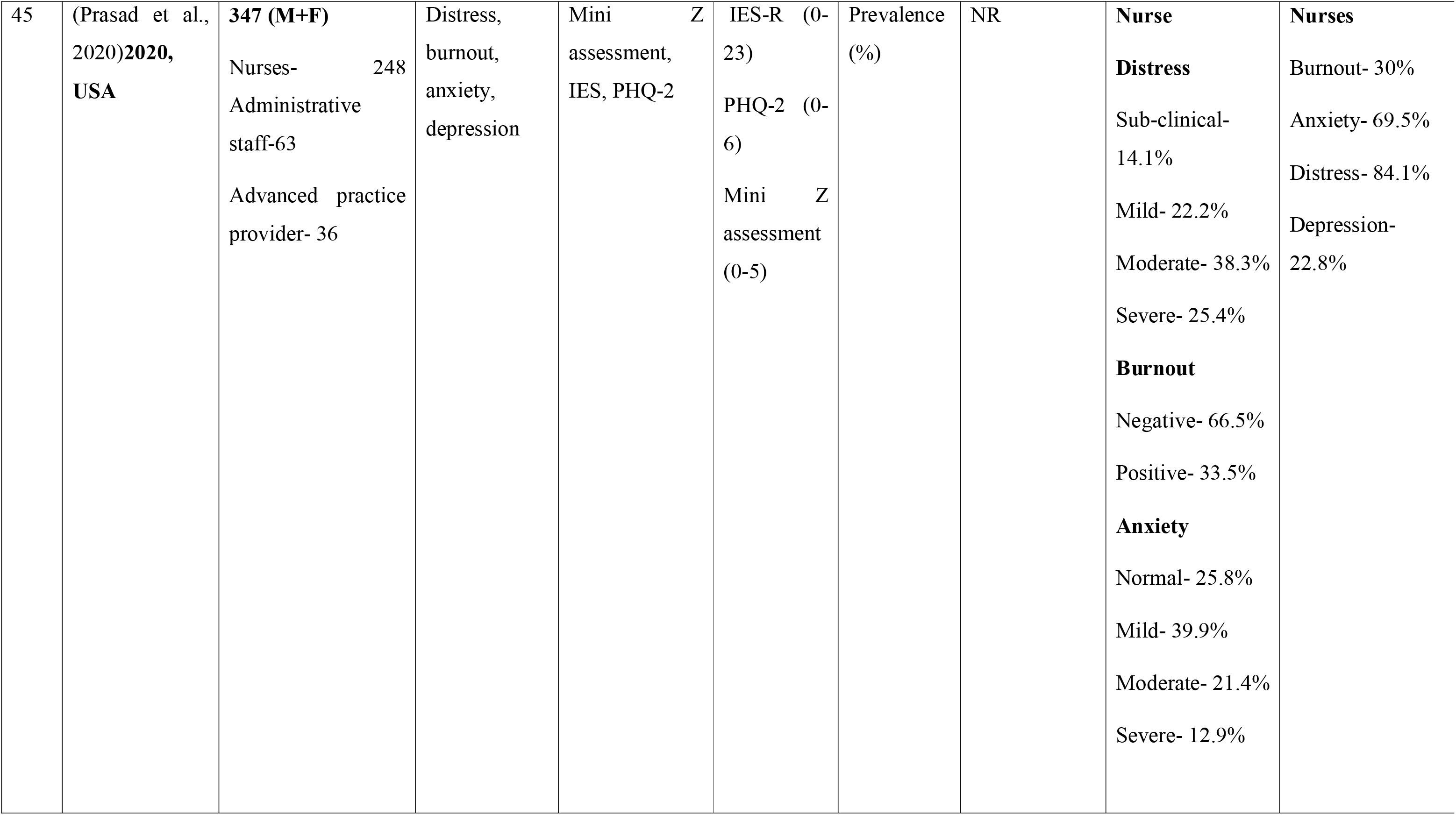
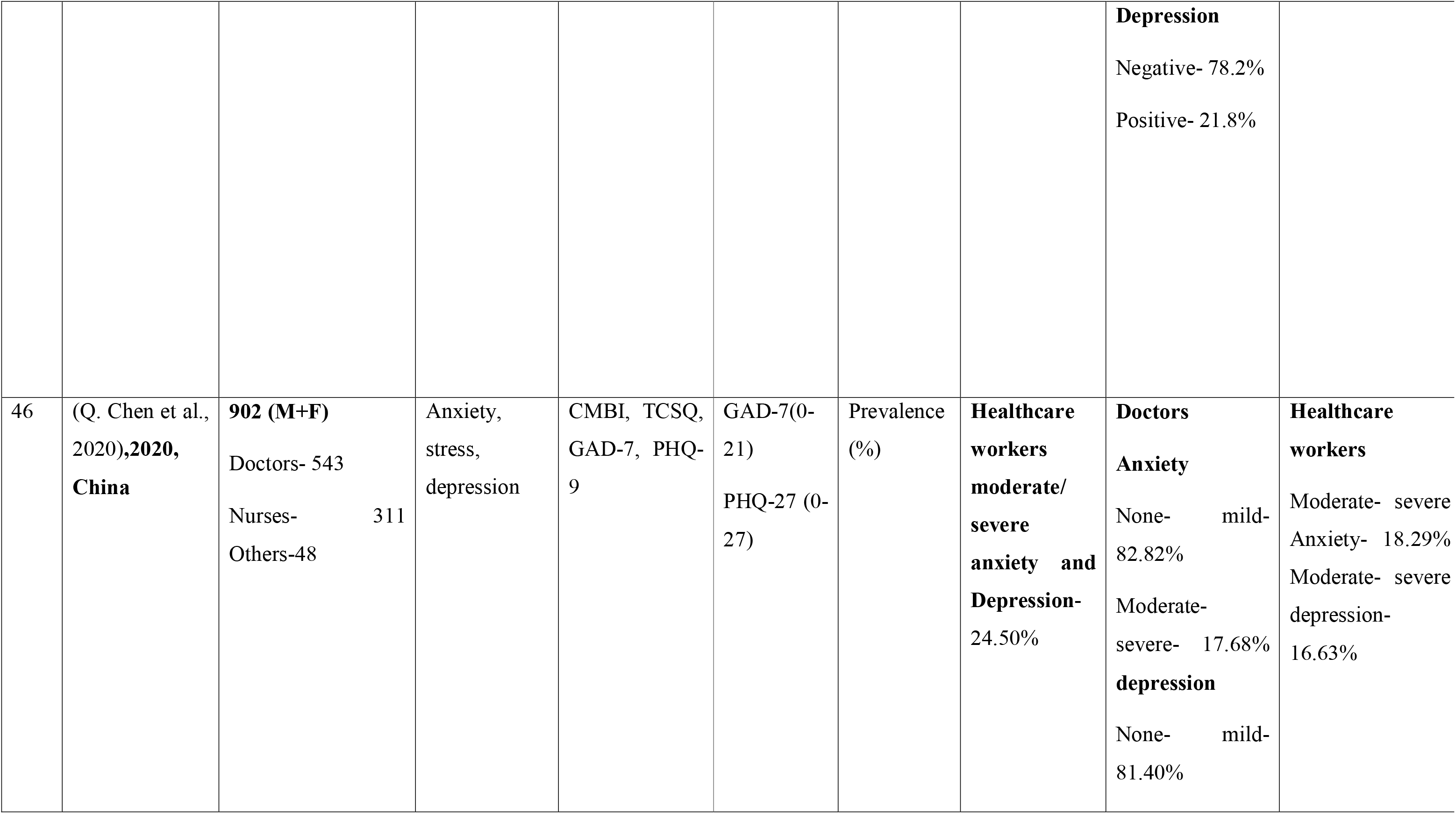
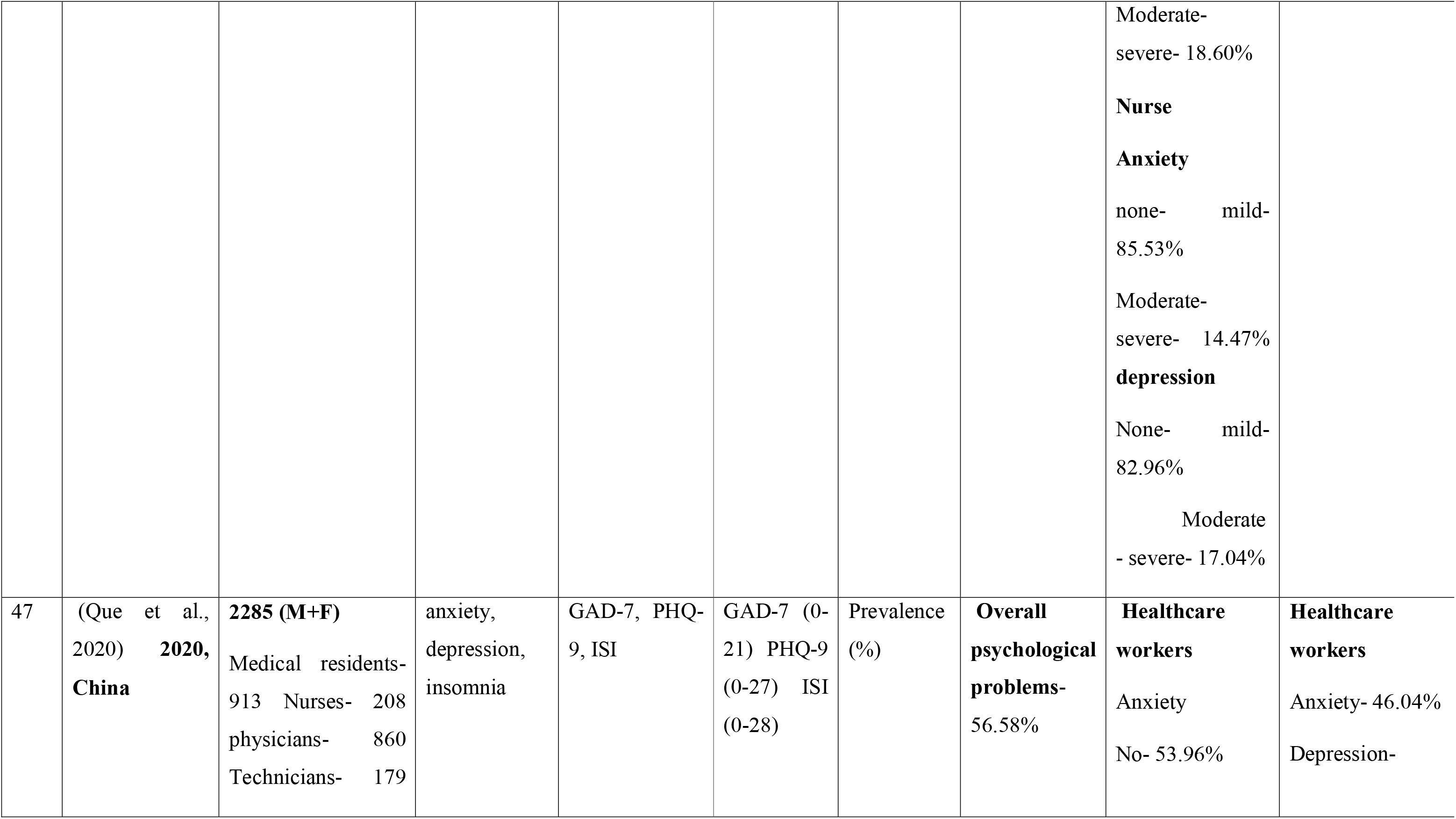
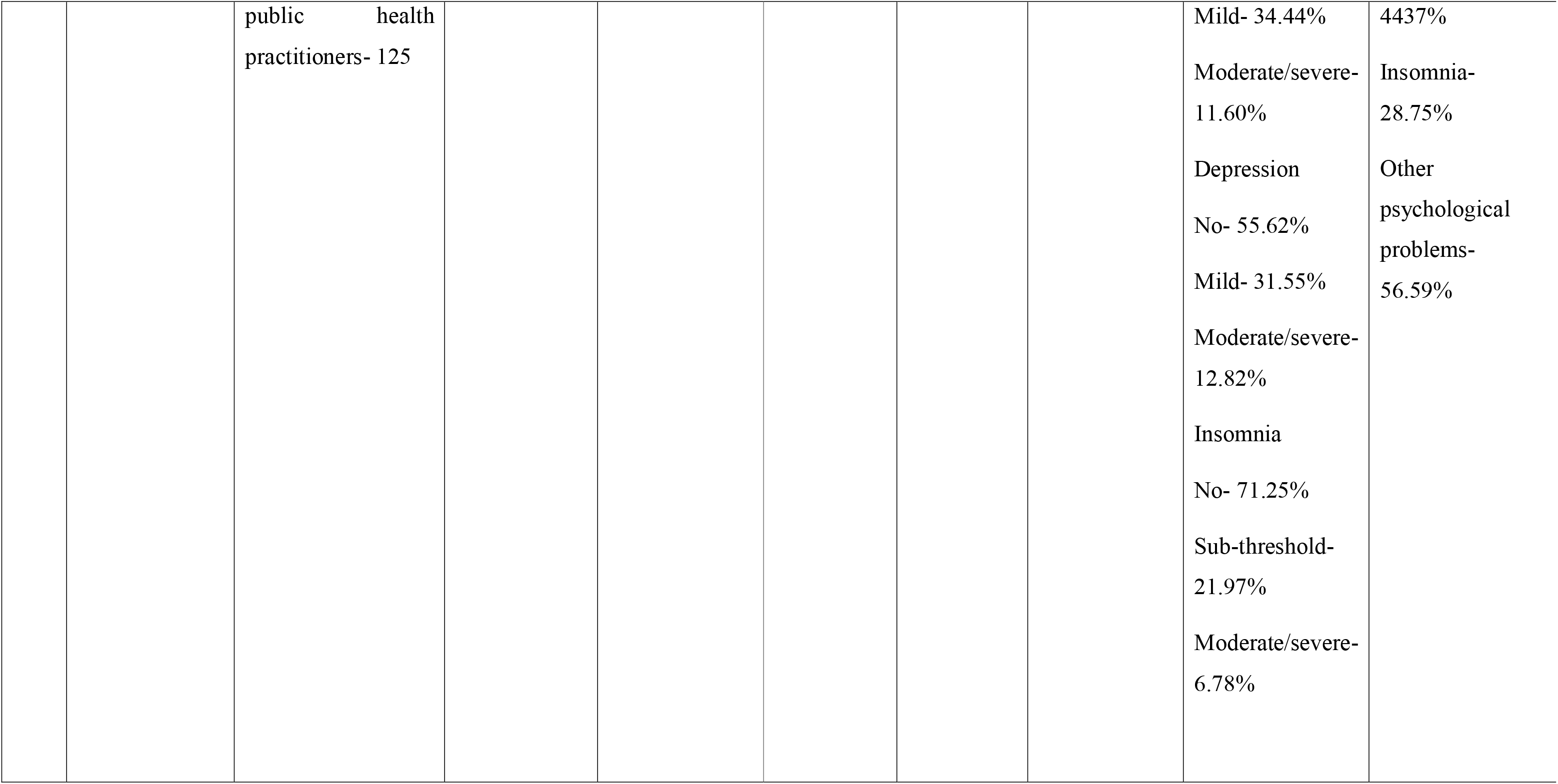
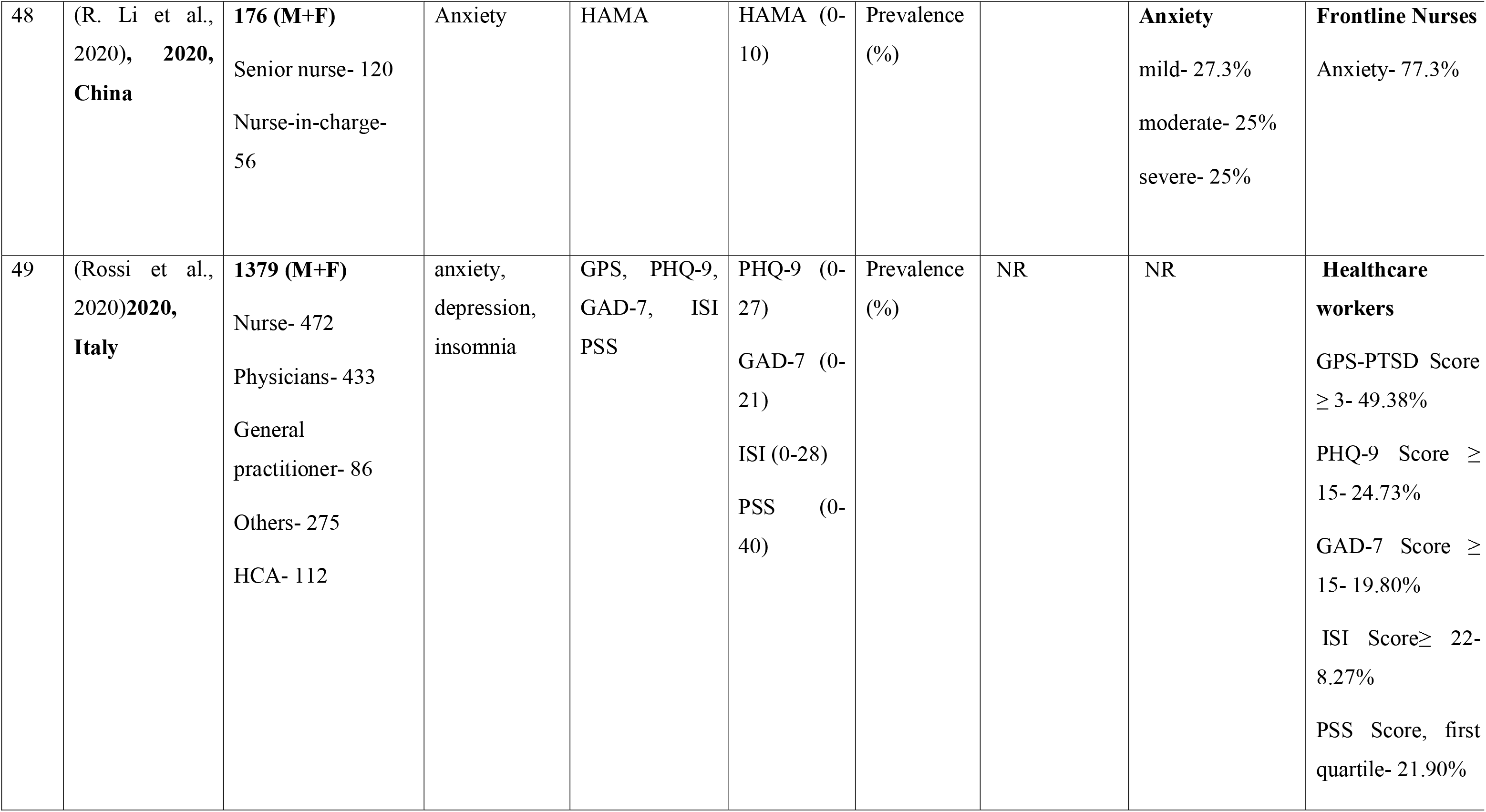
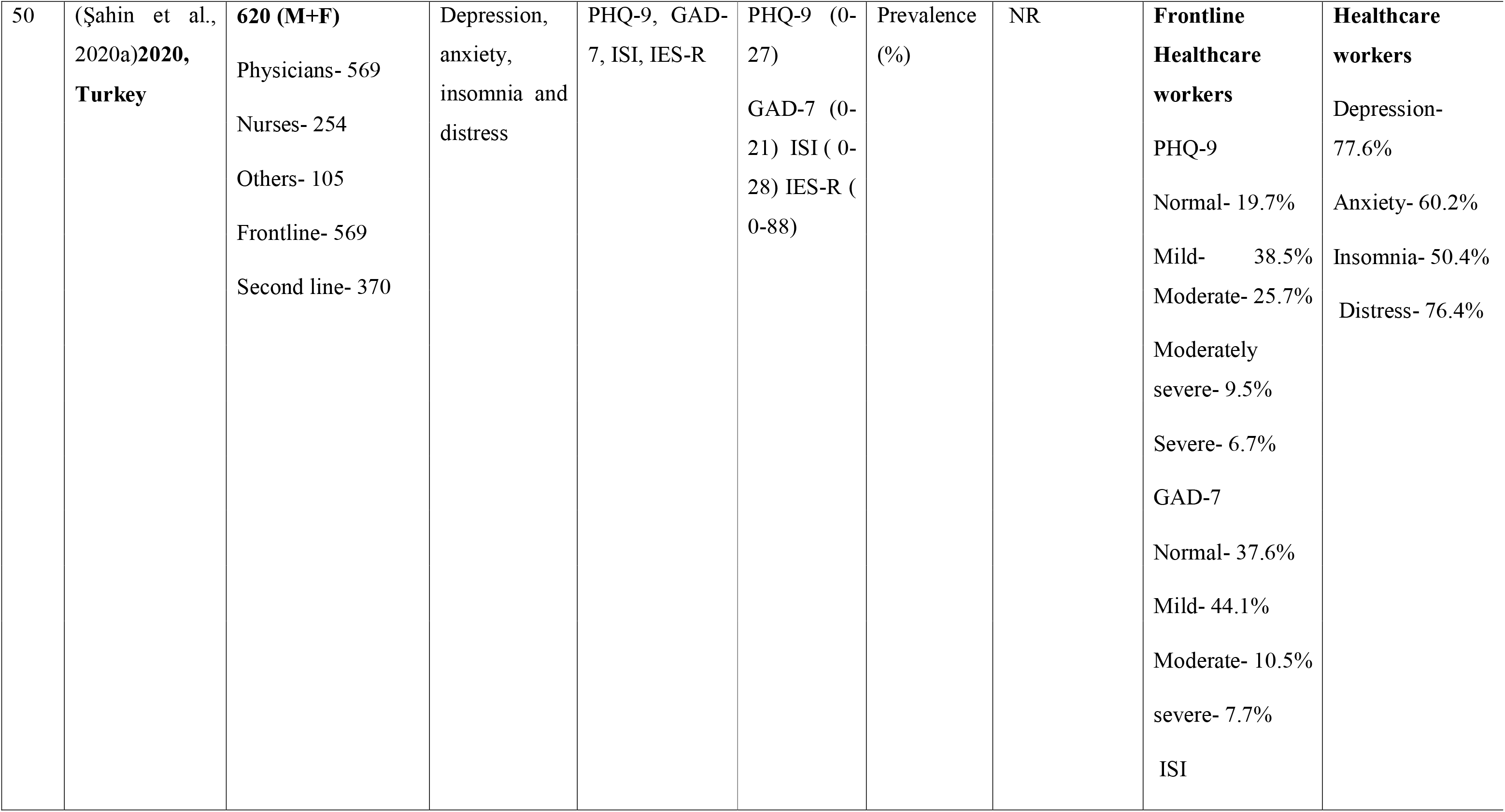
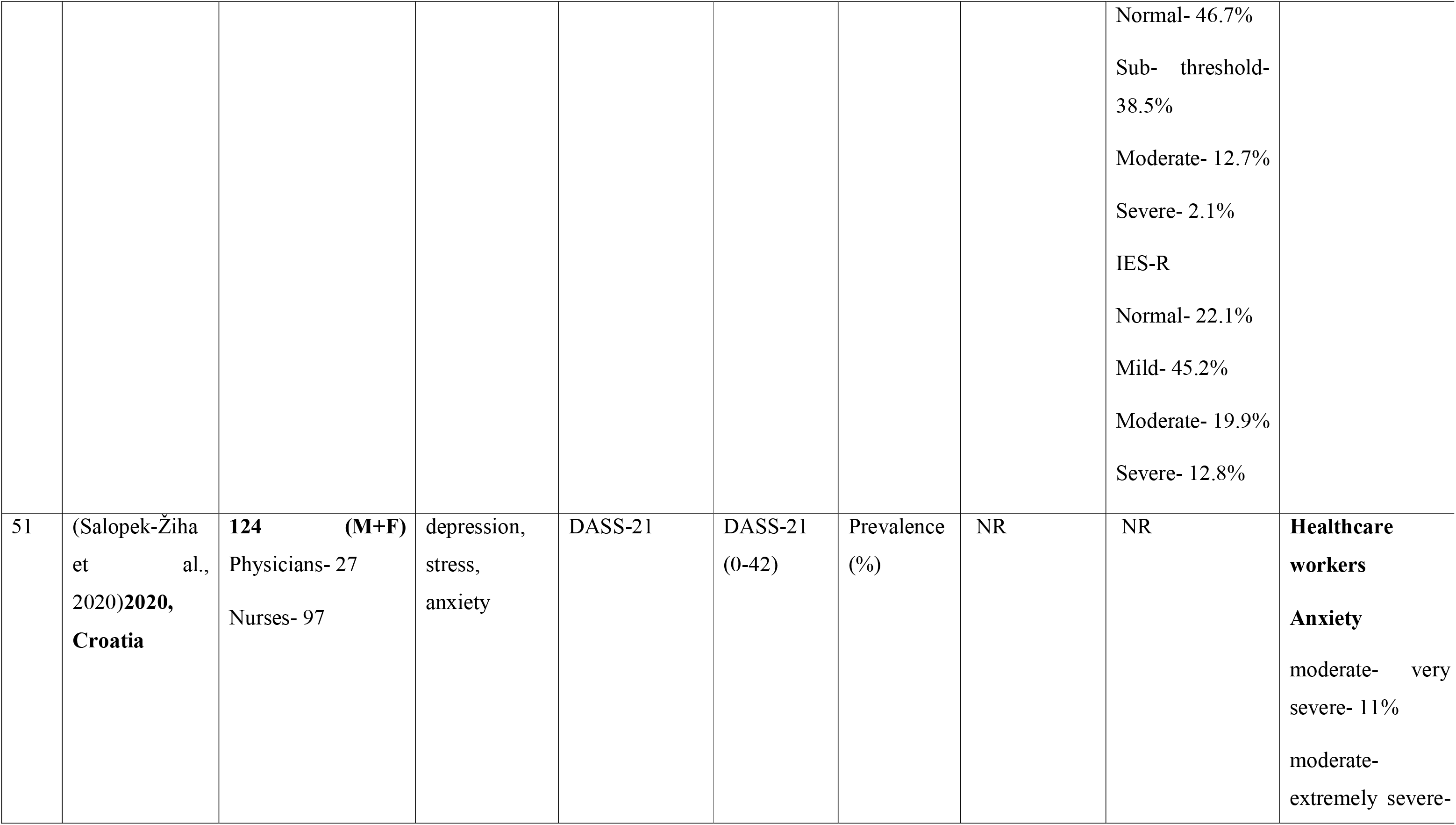
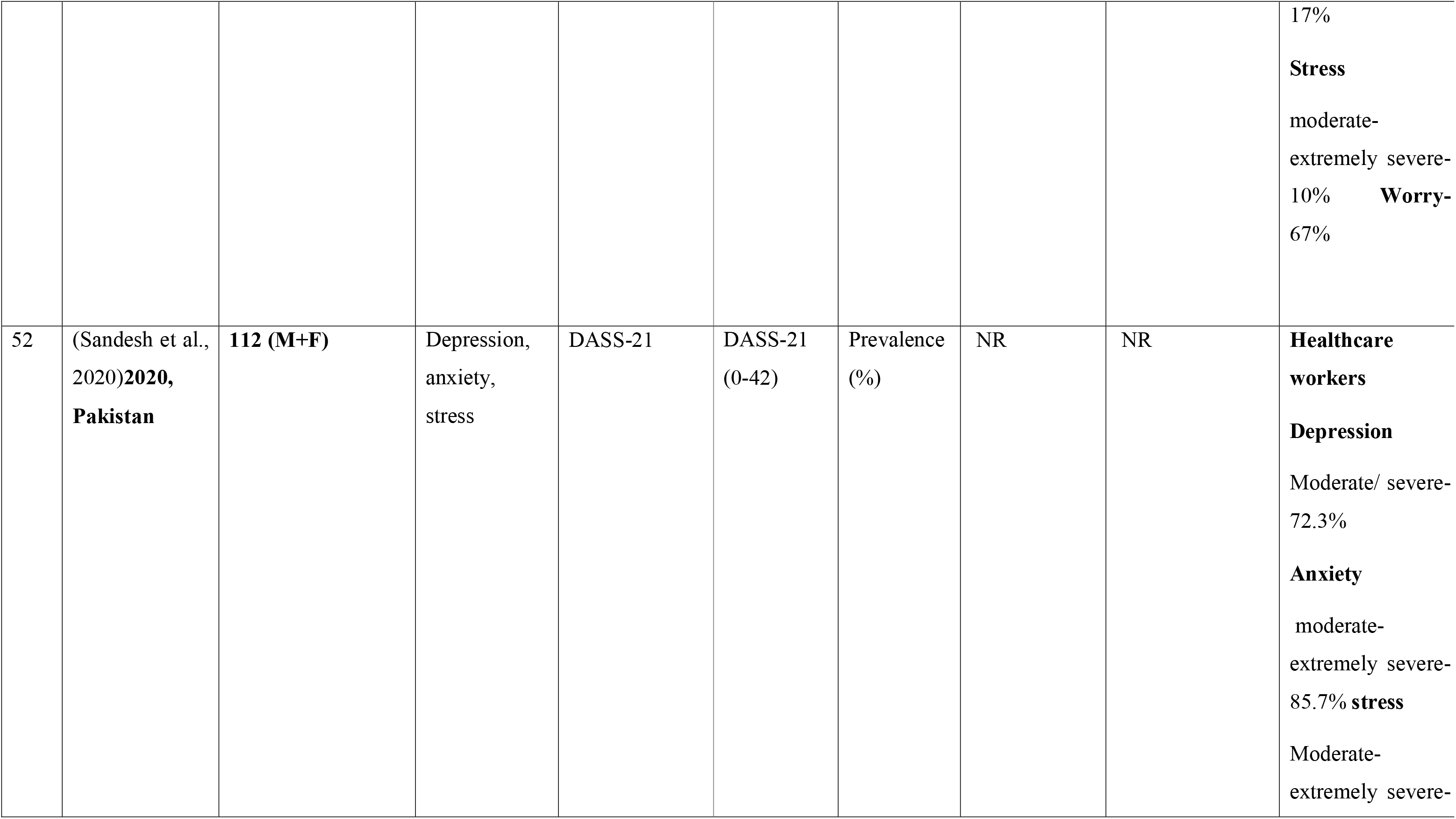
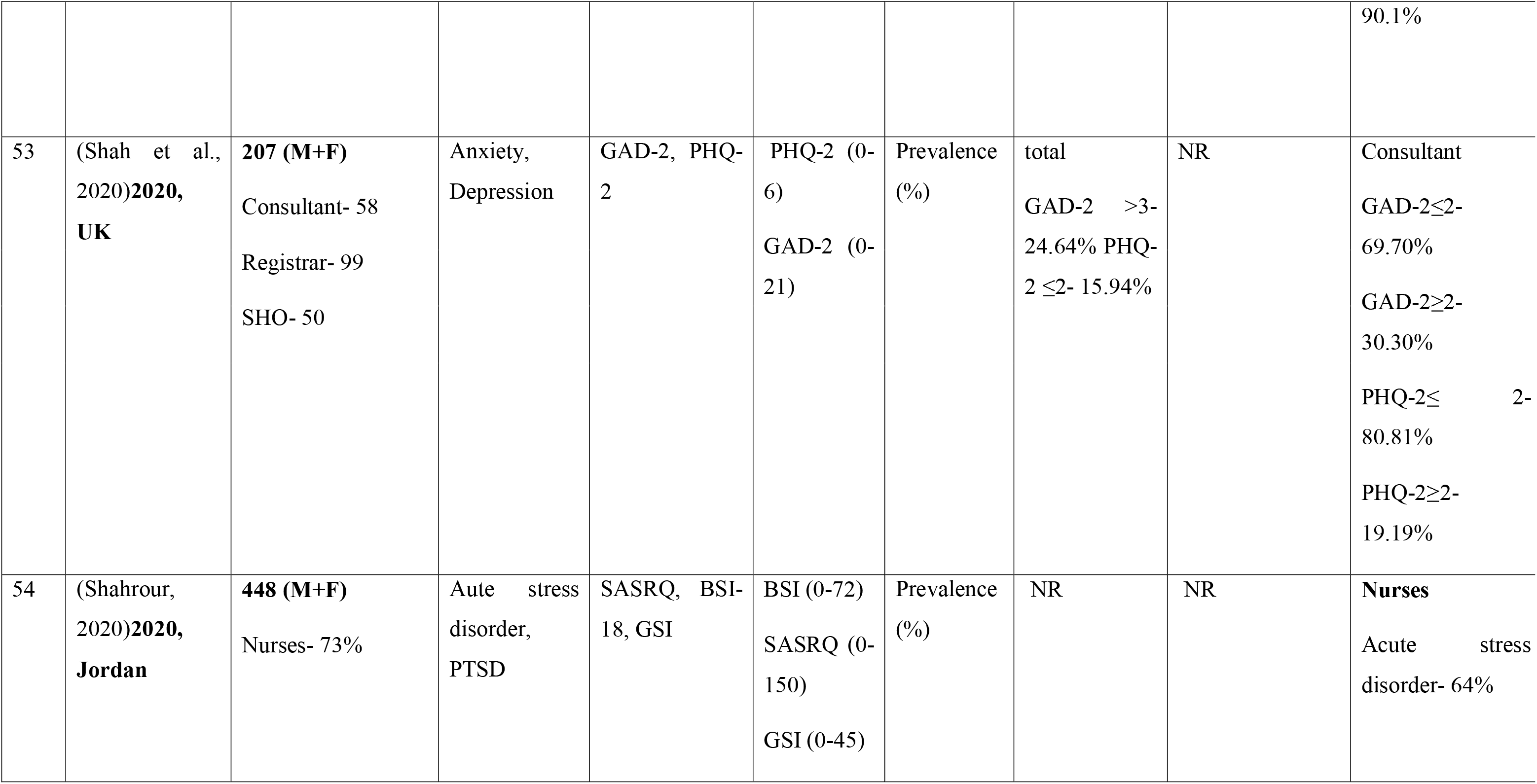
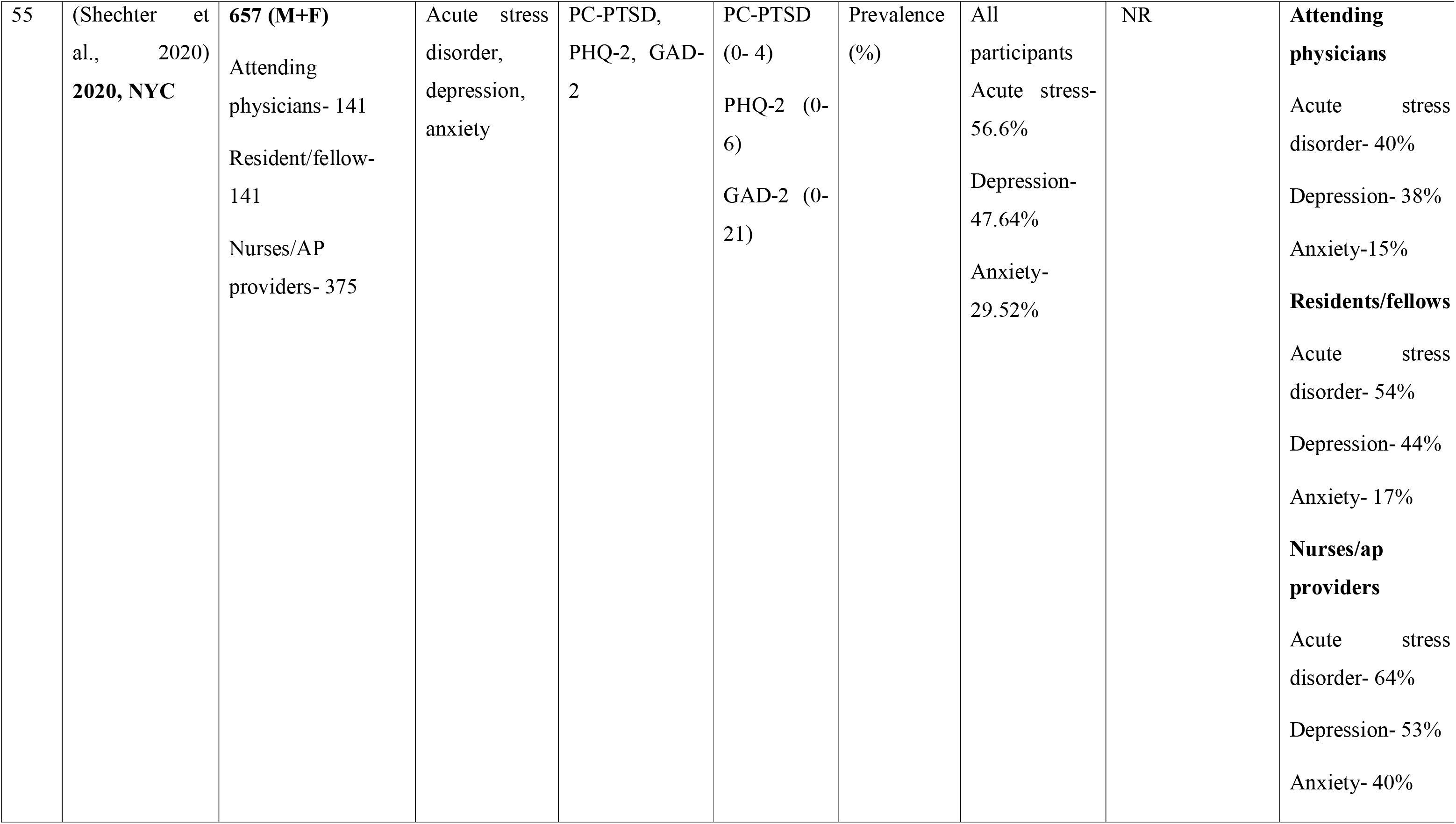
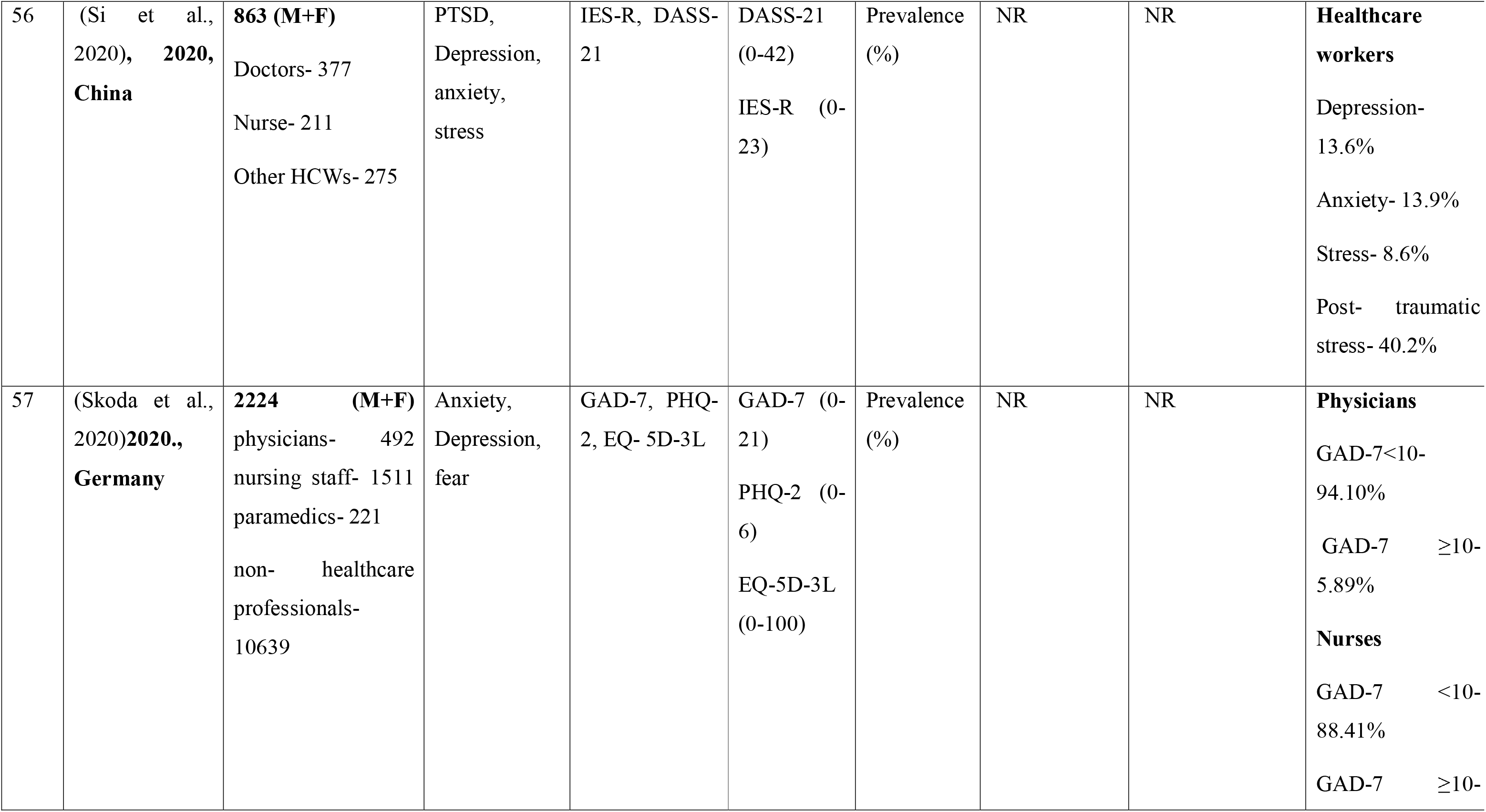
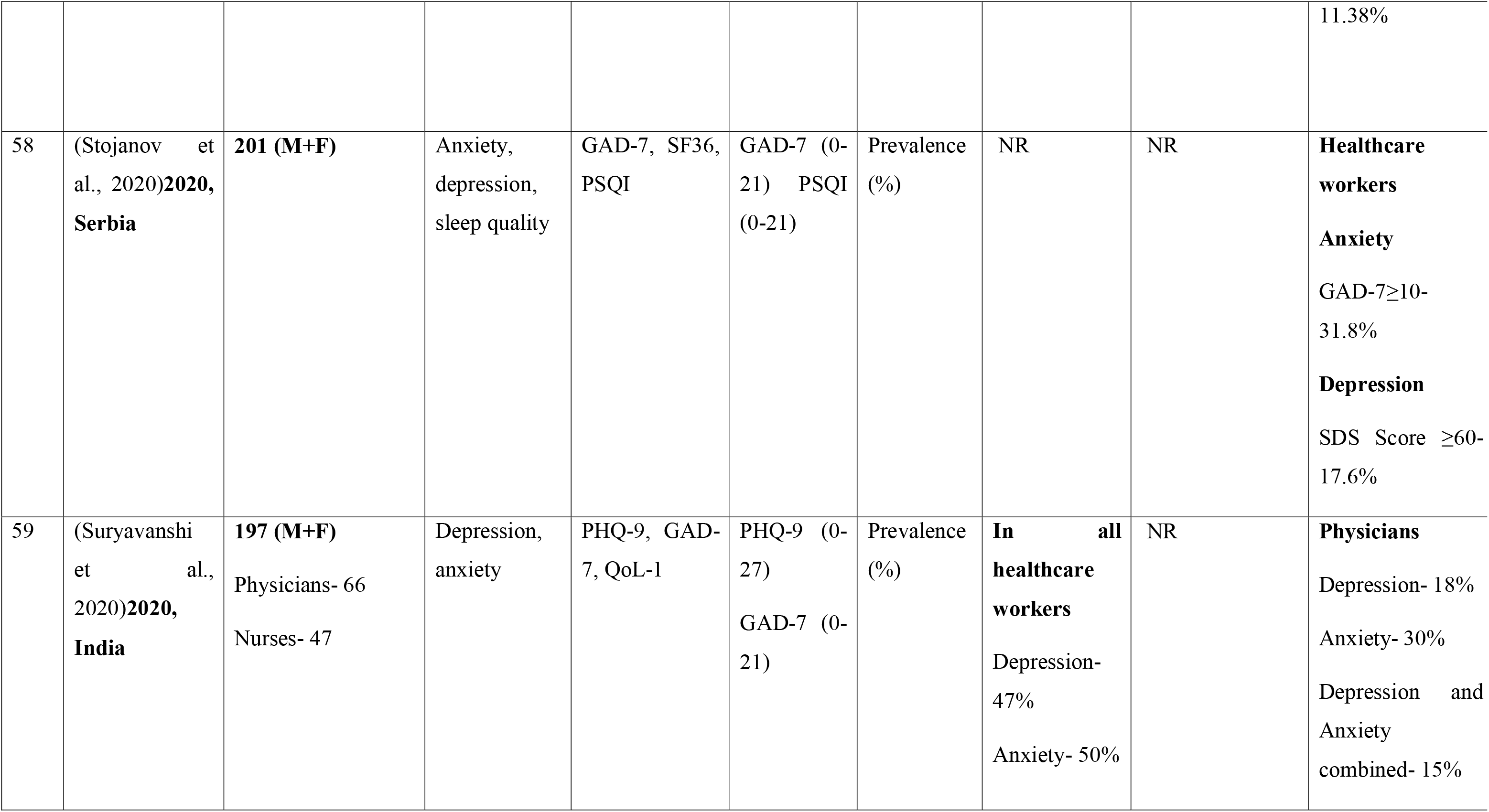
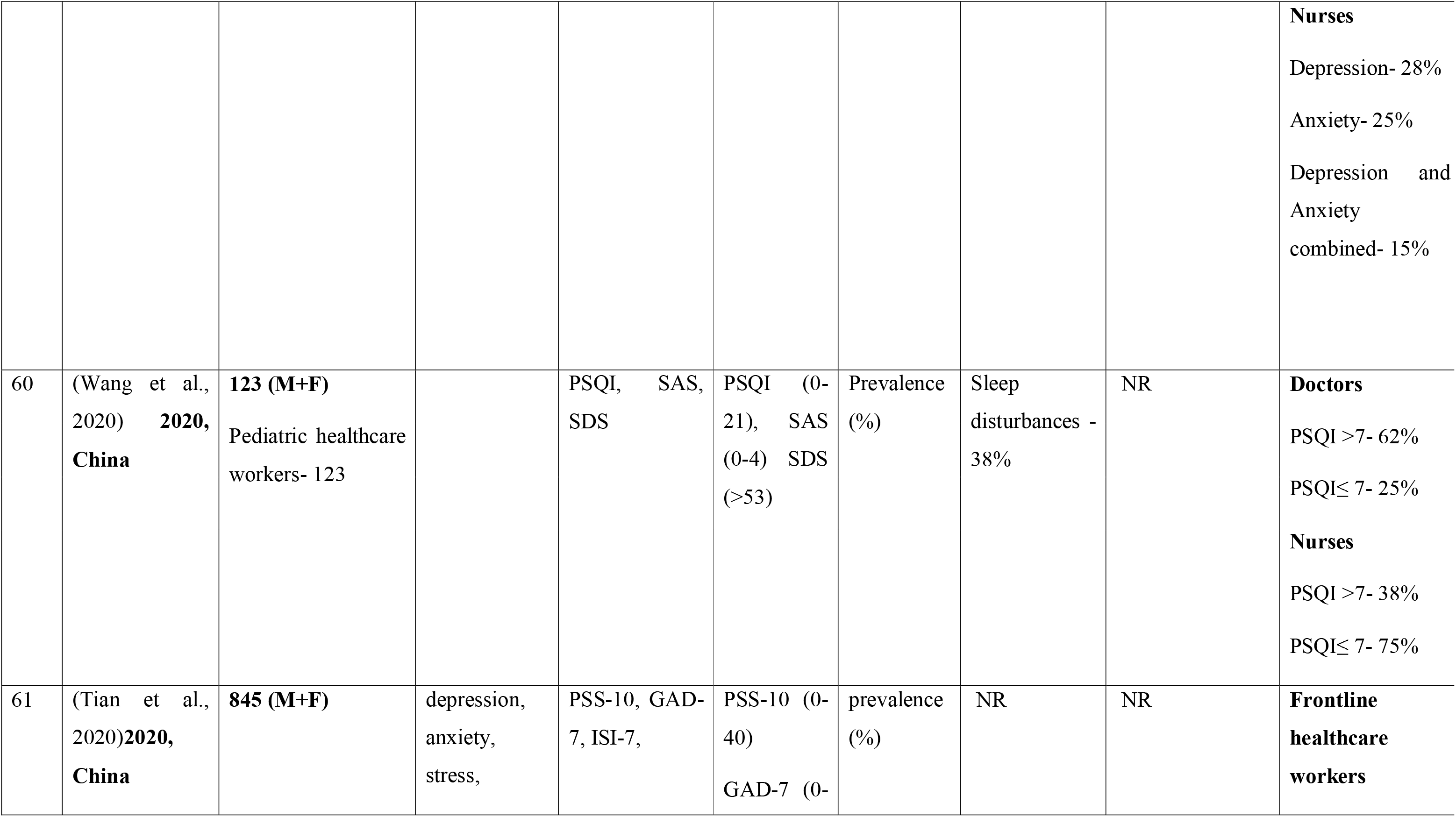
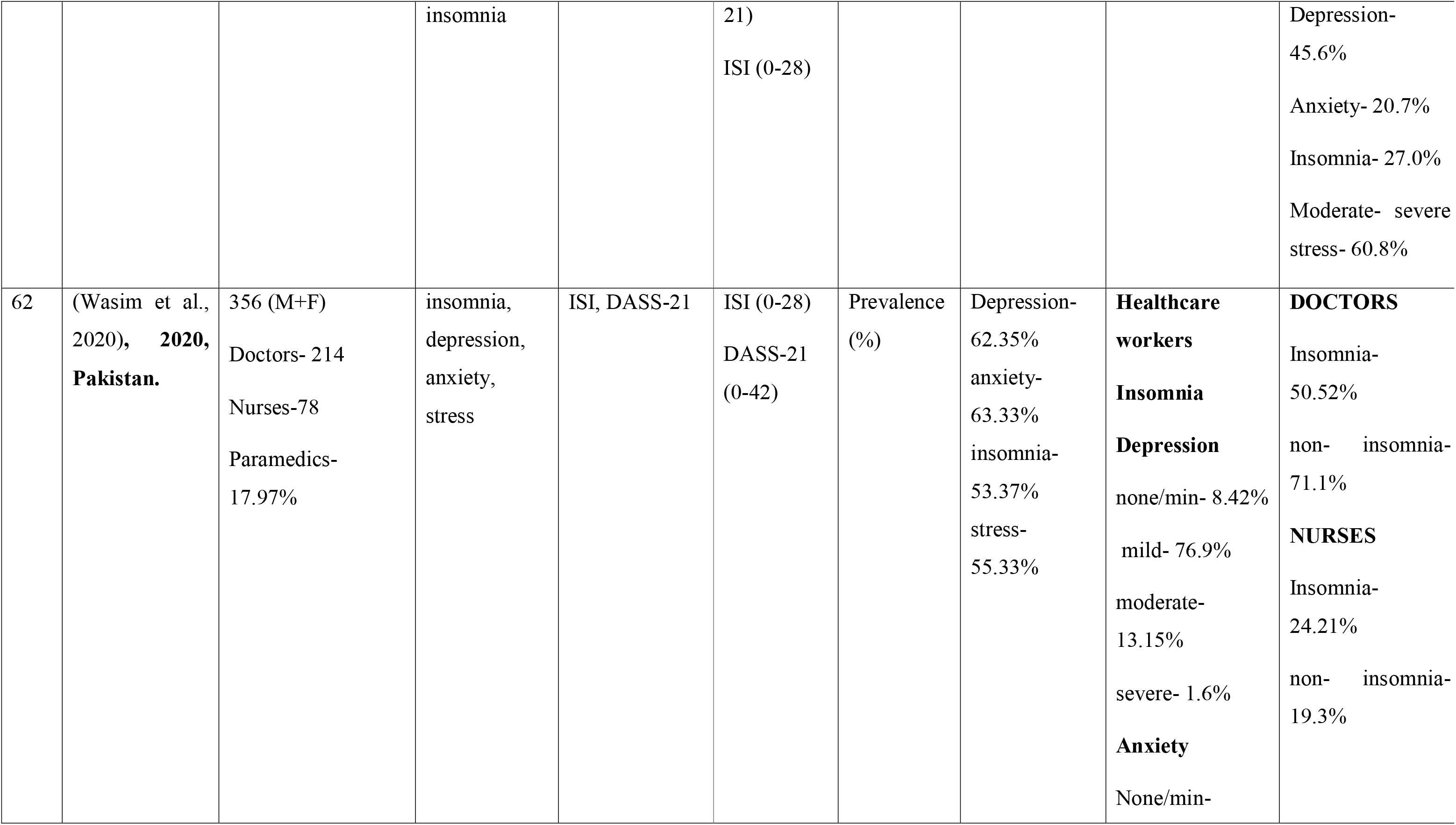
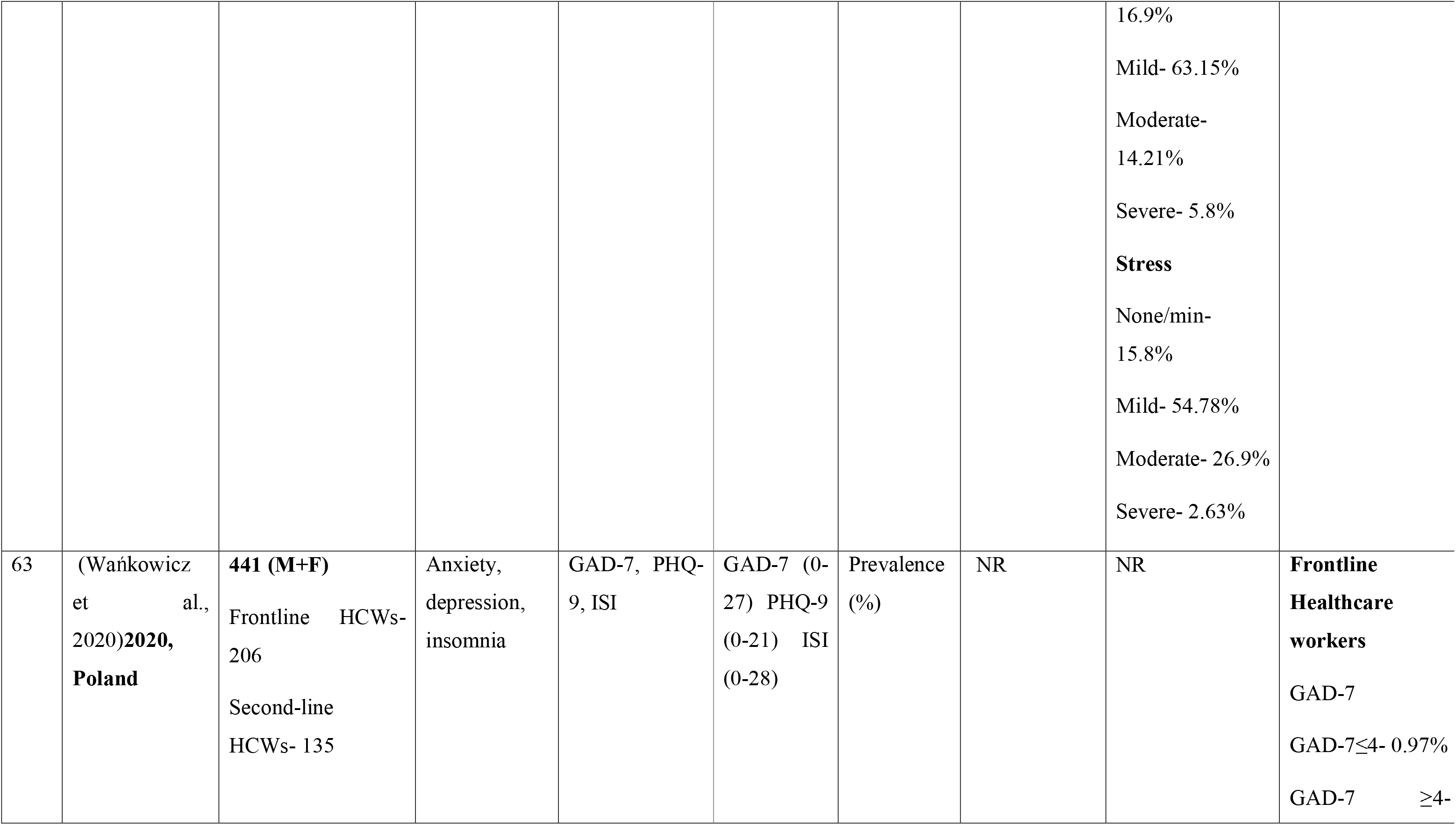
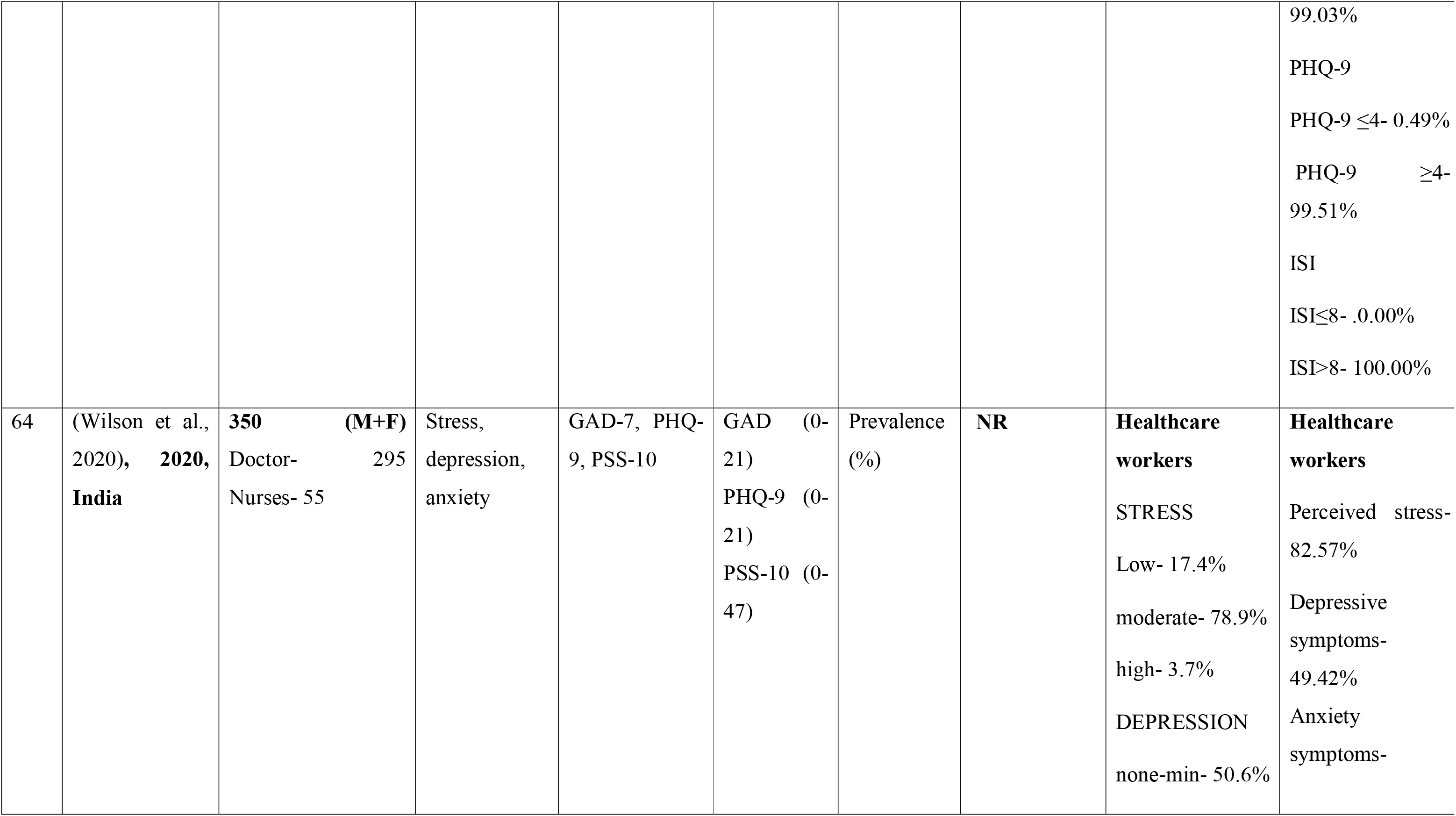
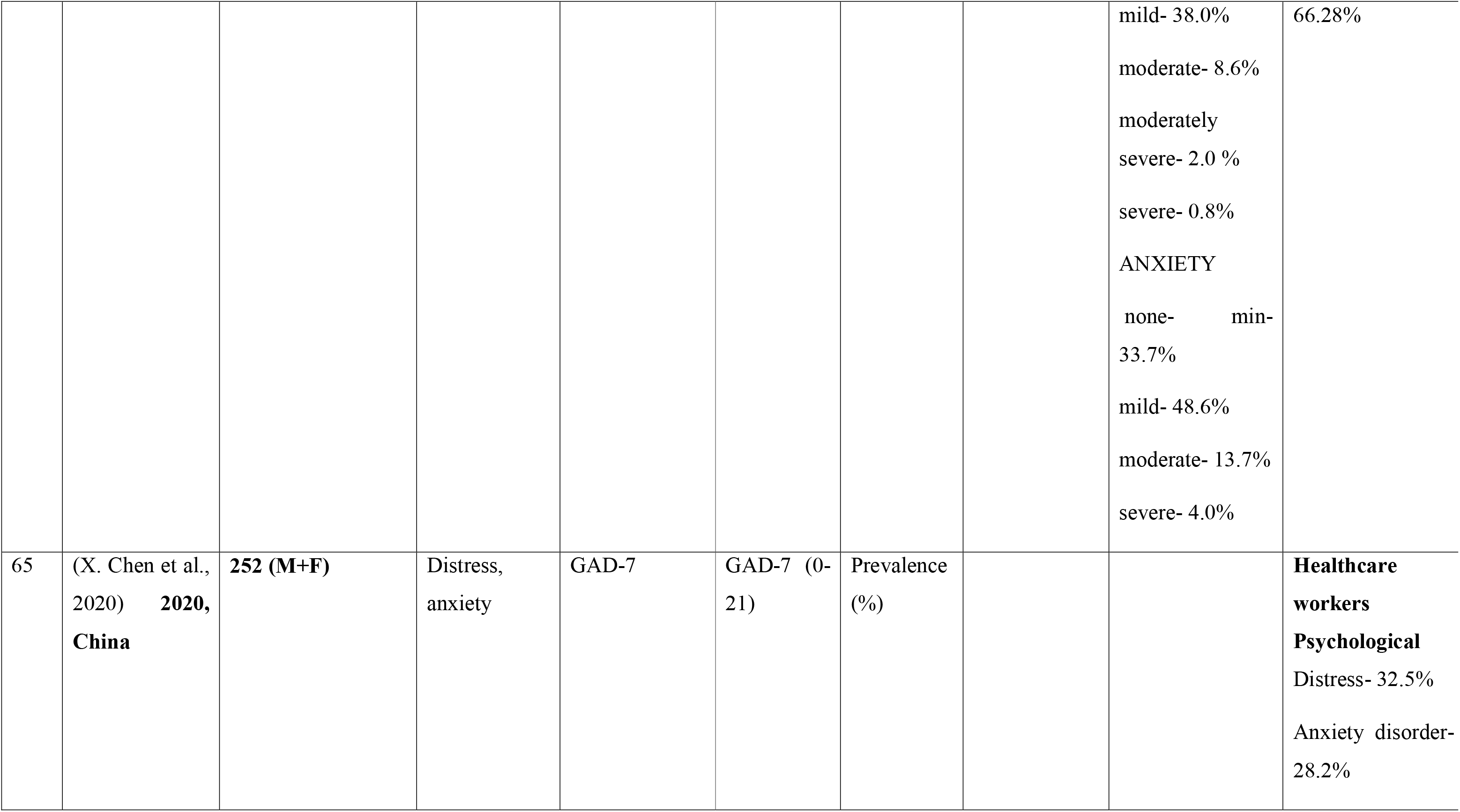
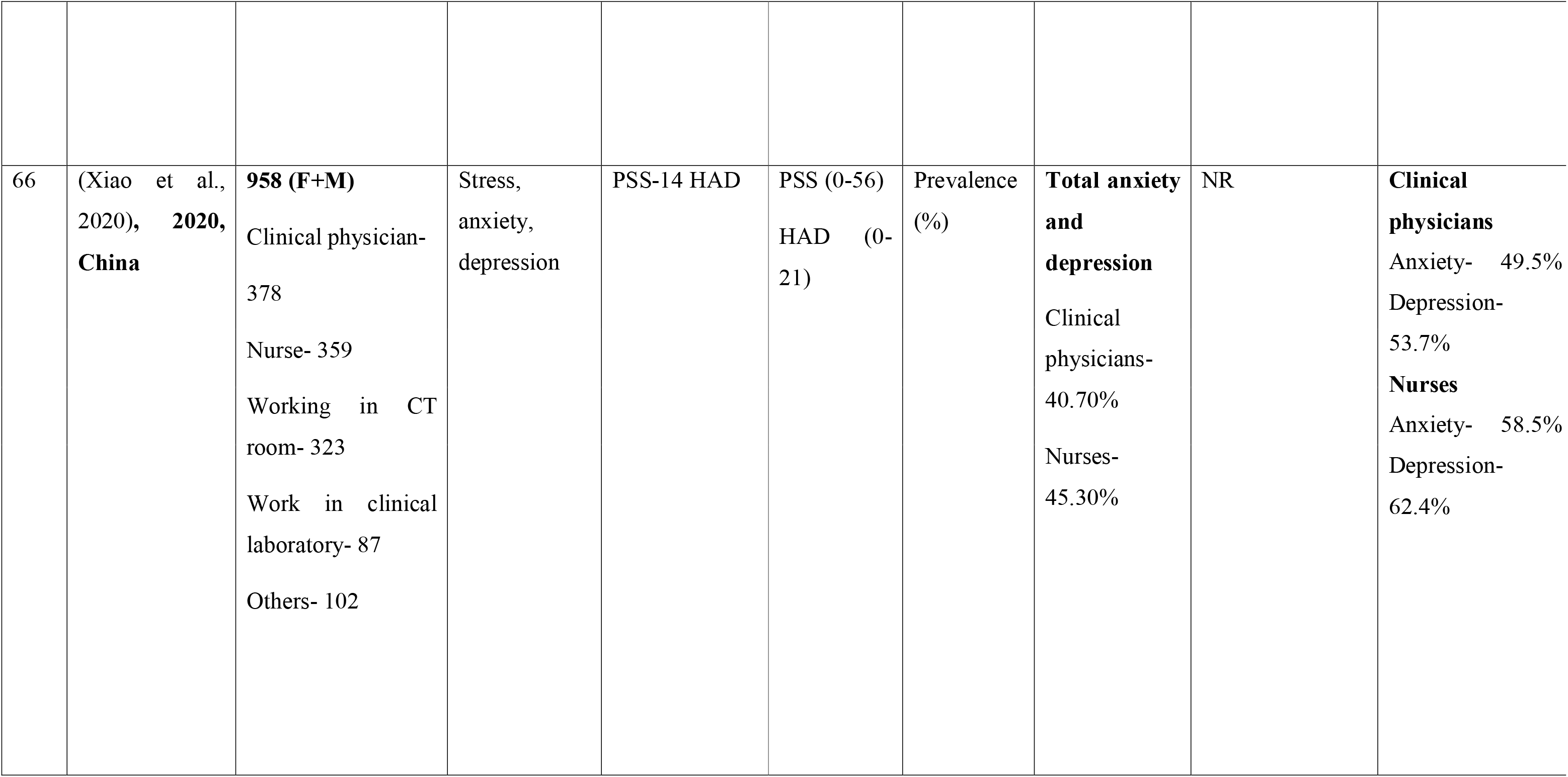
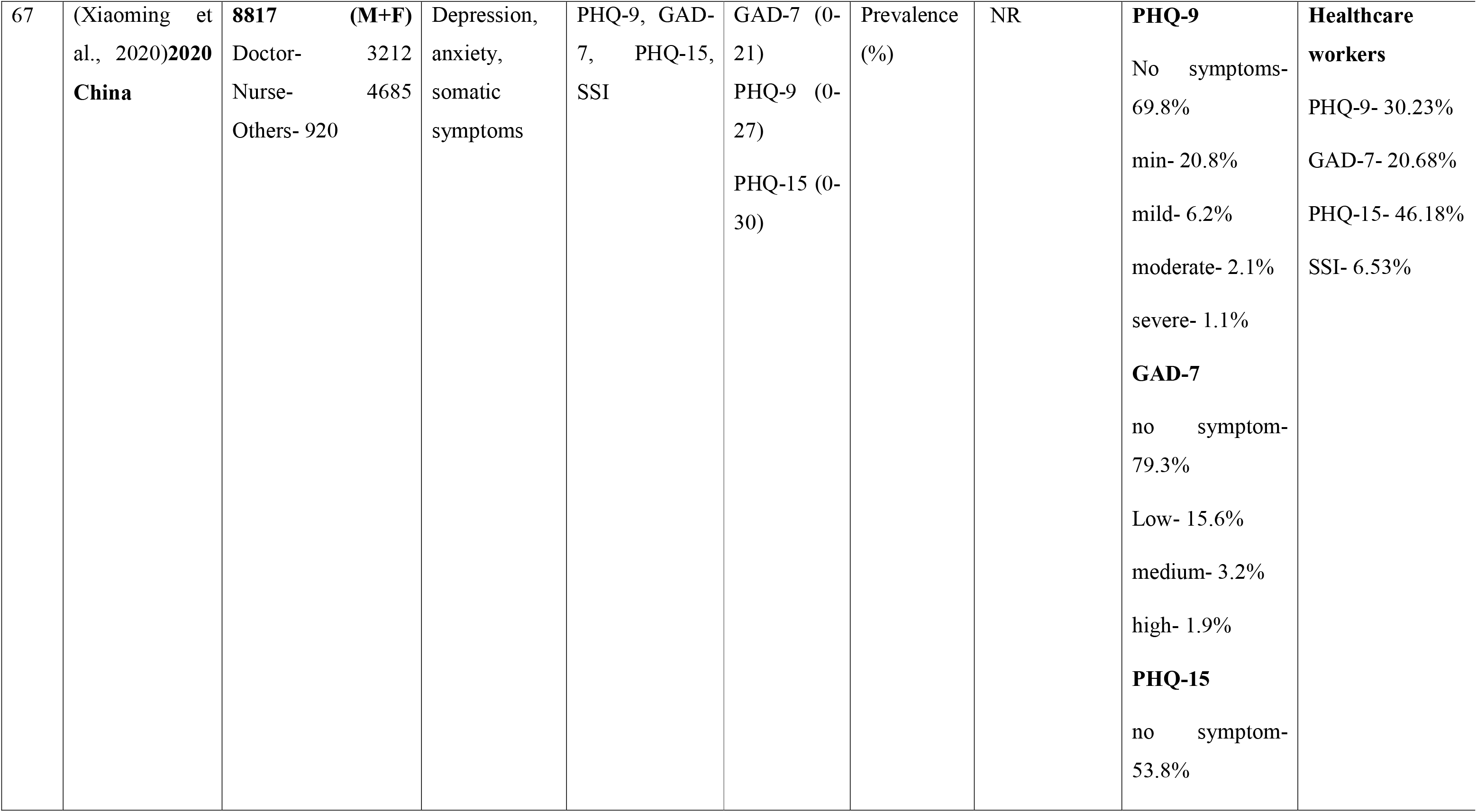
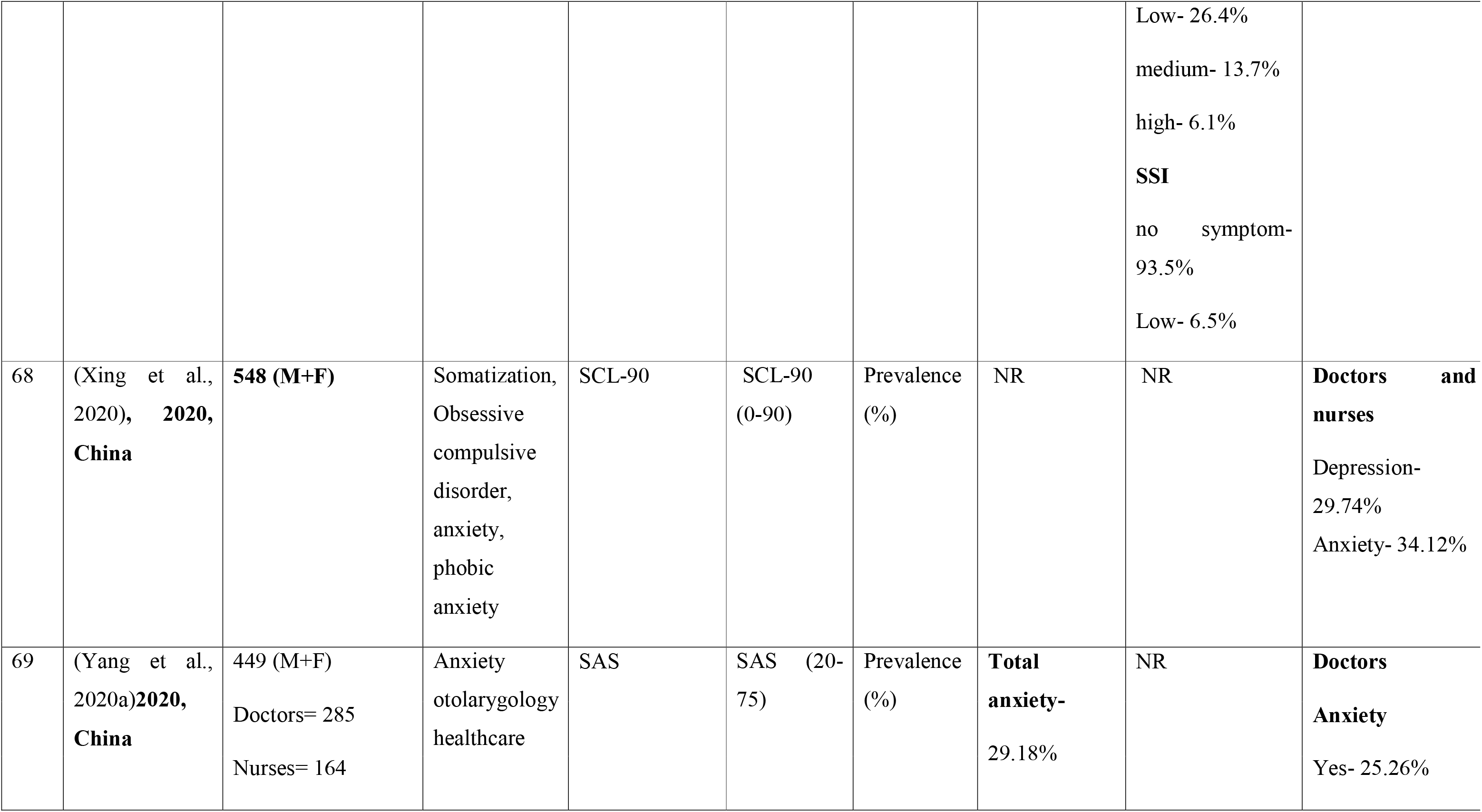
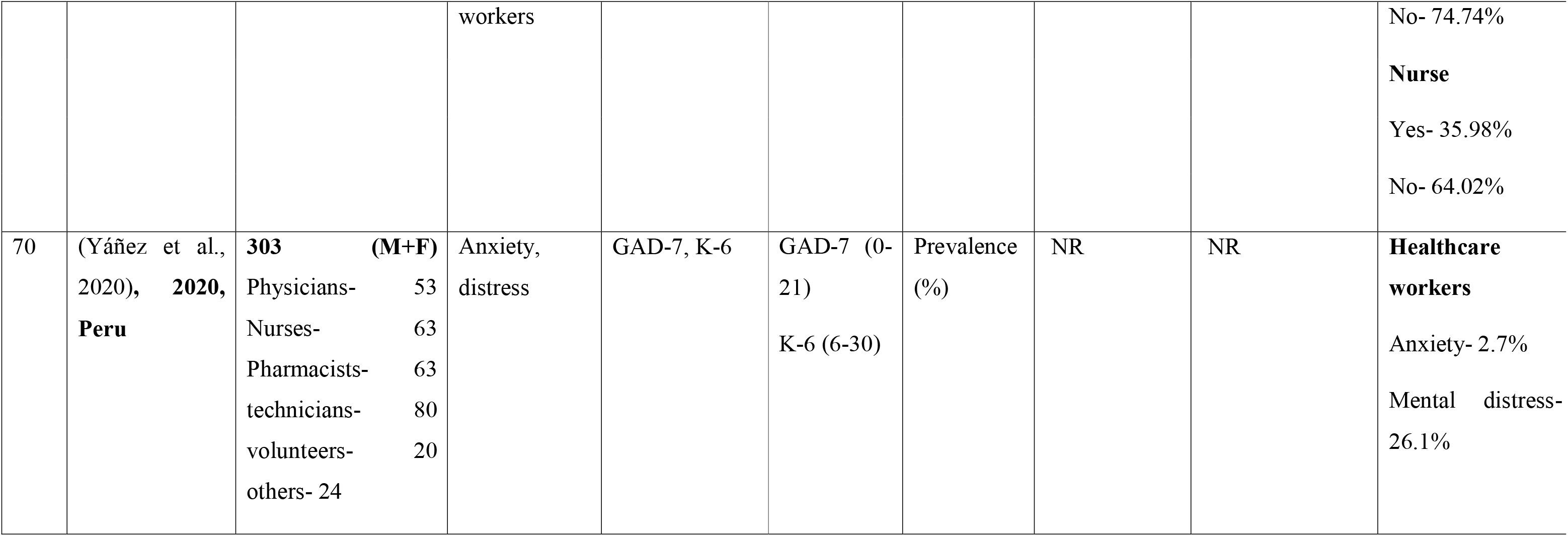
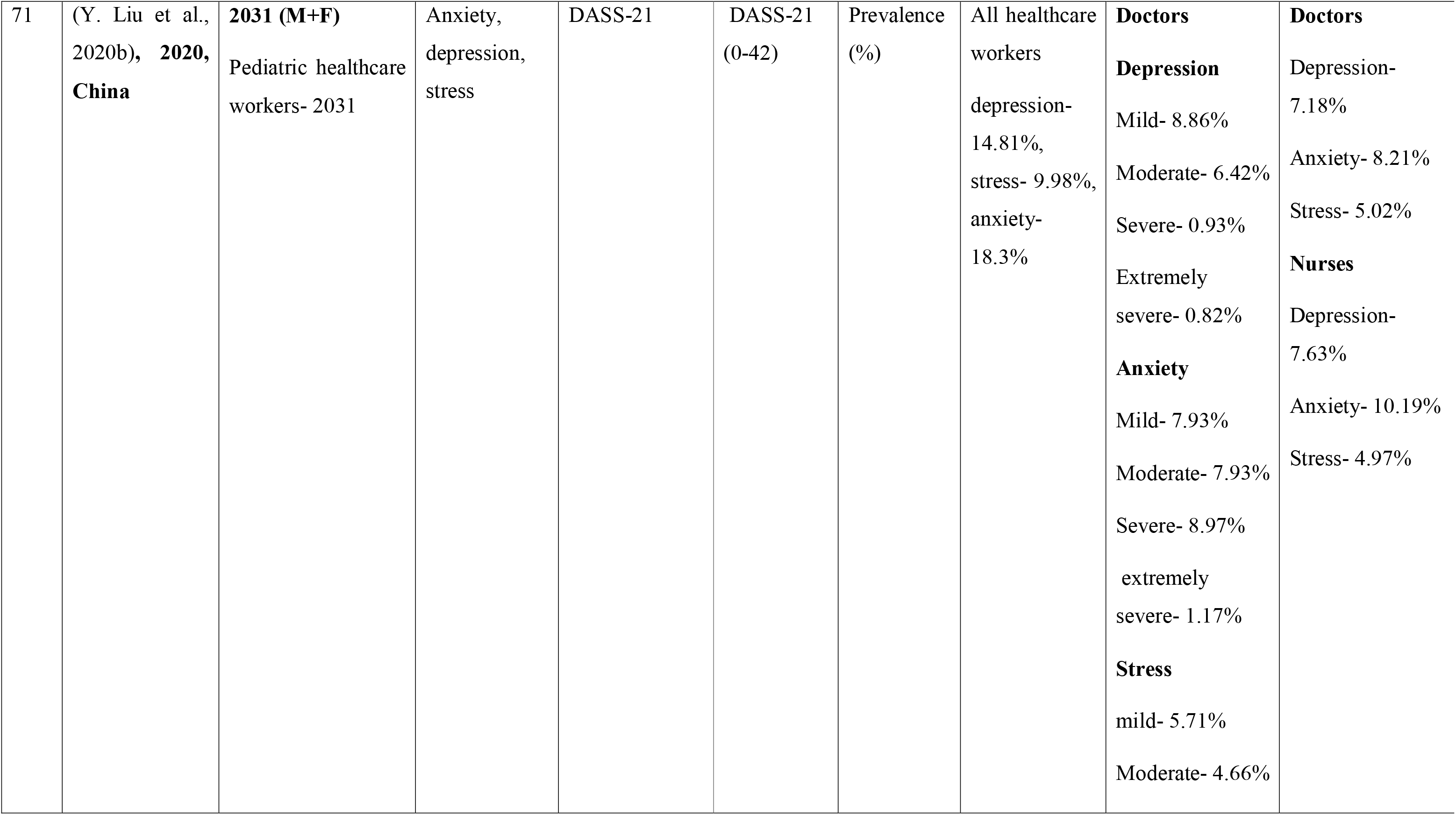
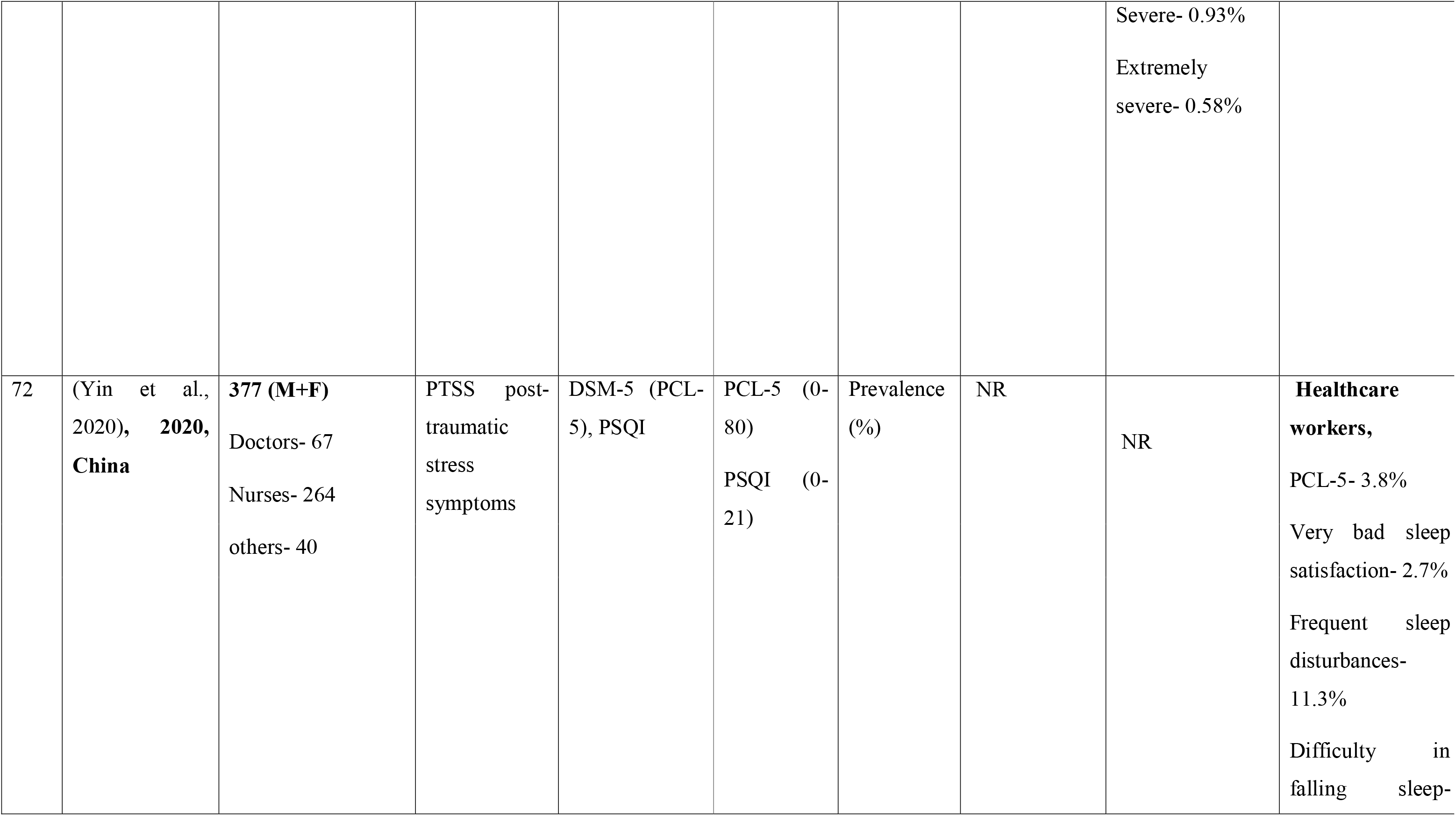
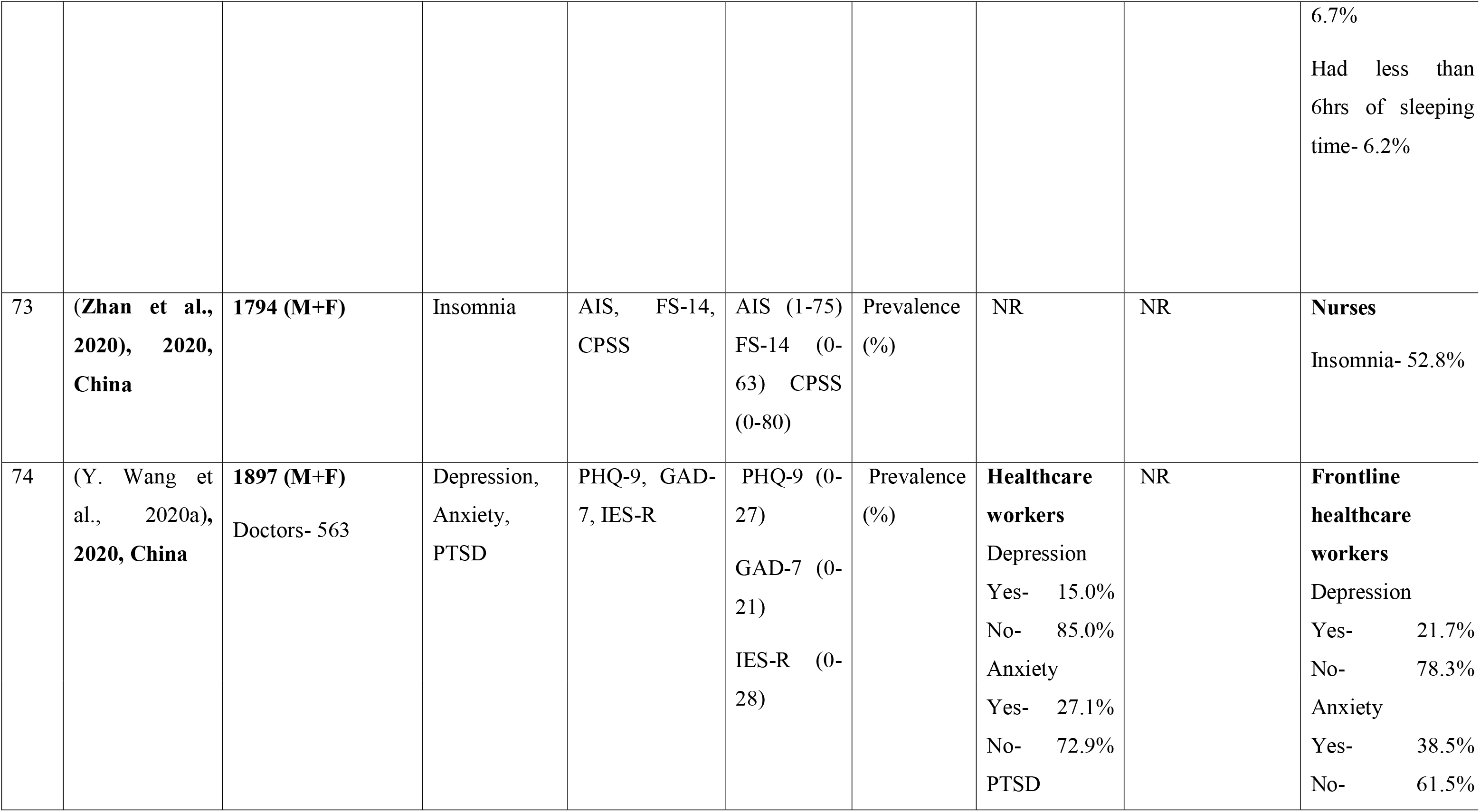
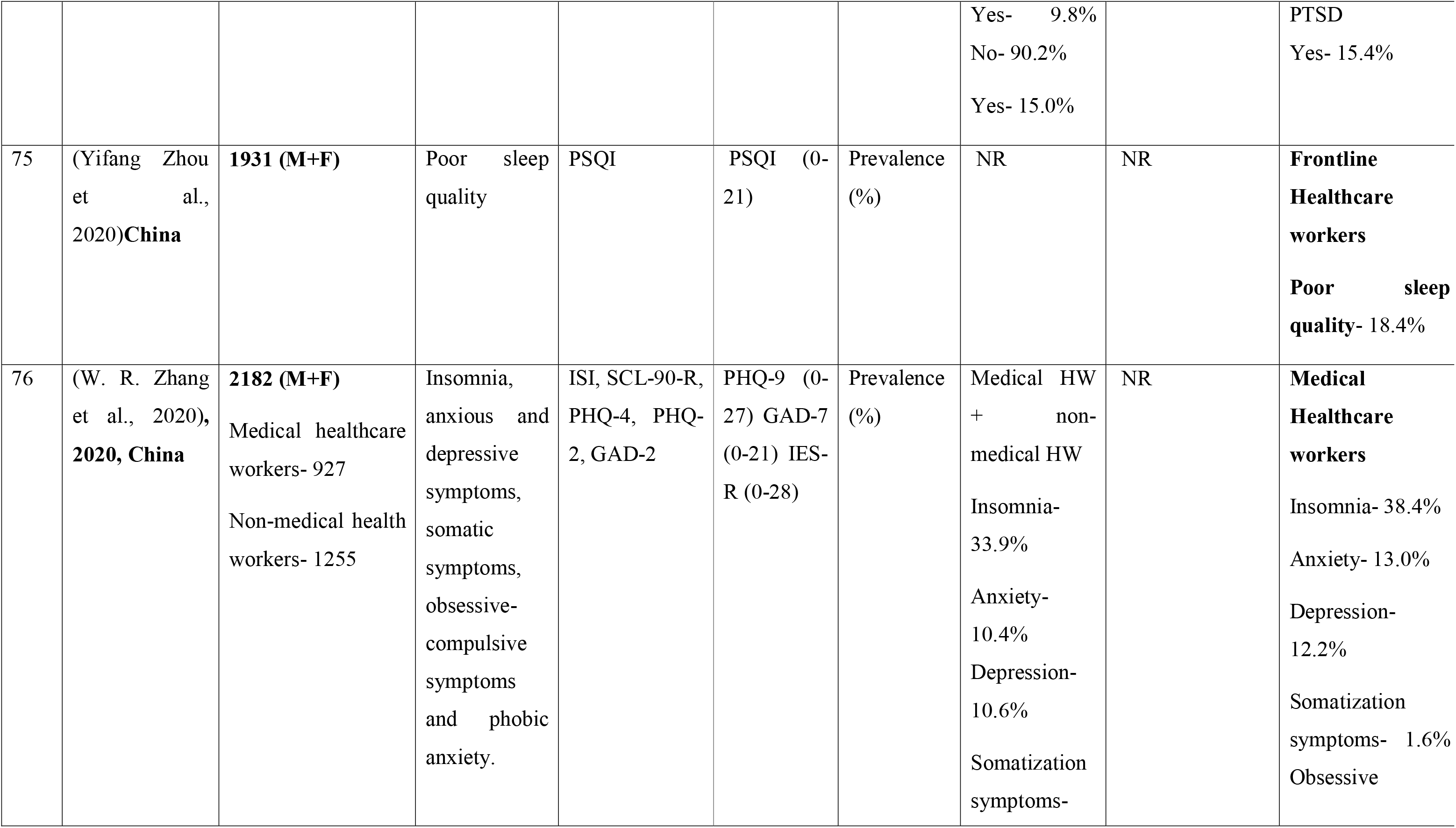
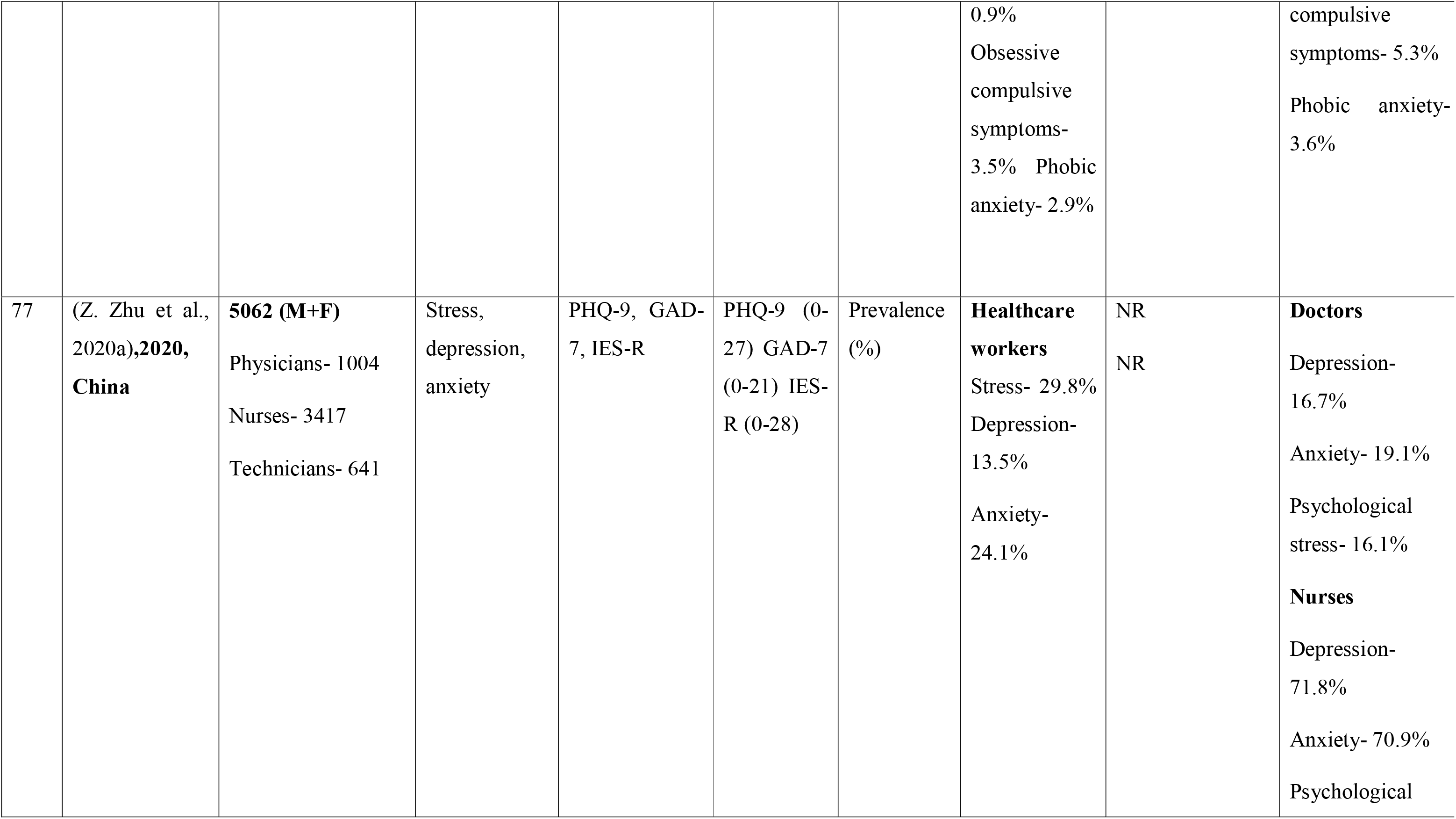
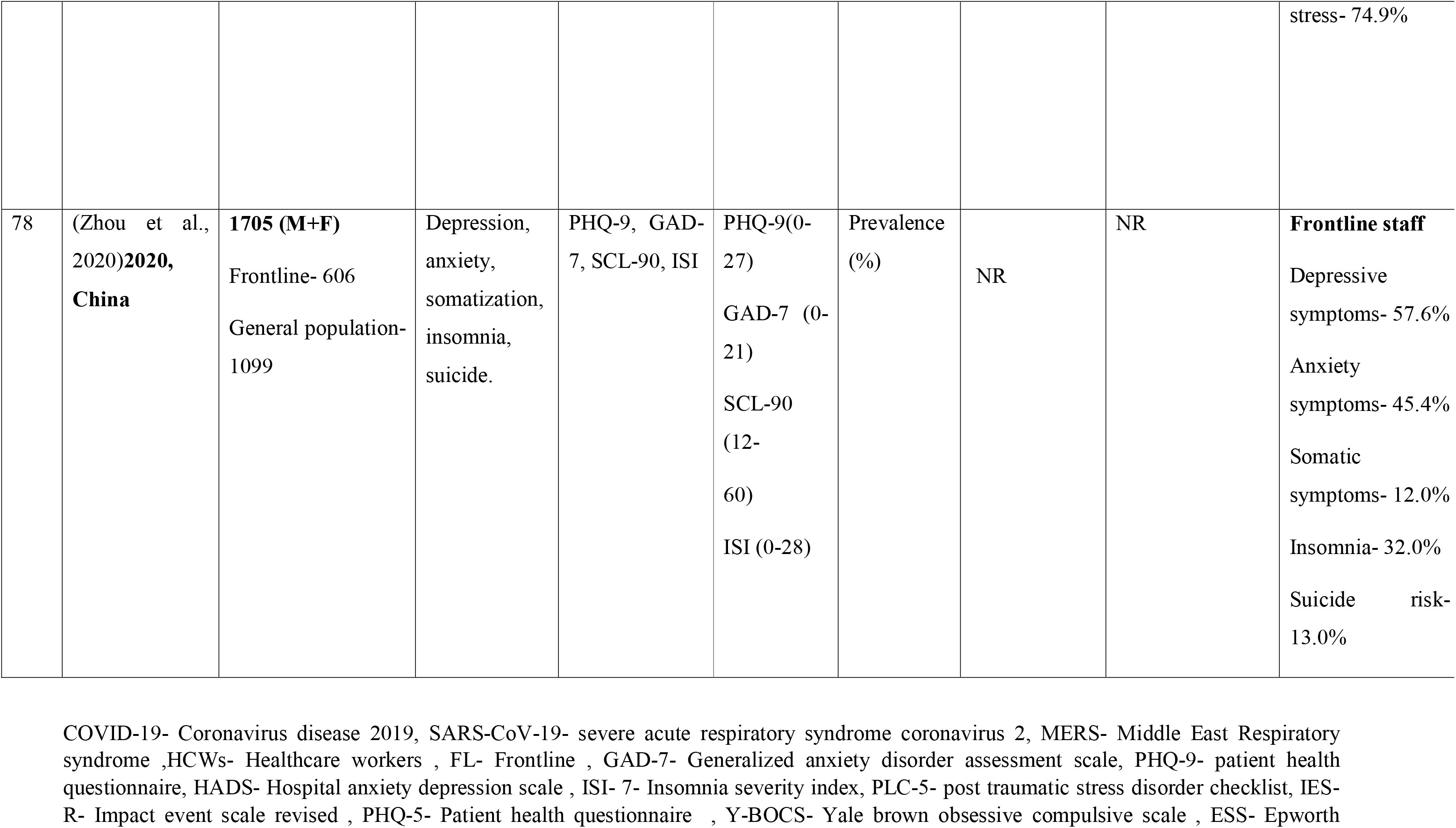
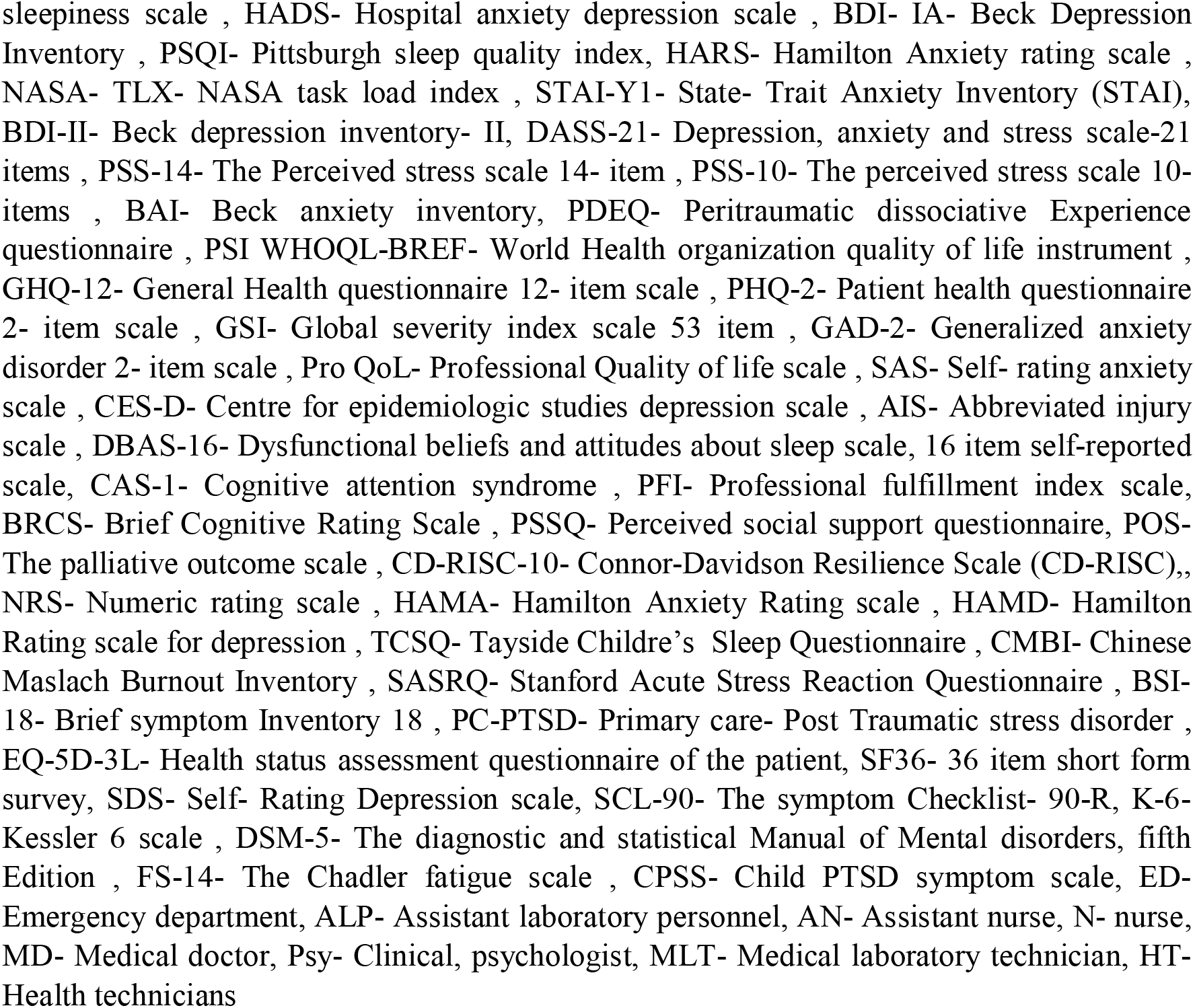
Summary of characteristics of included studies

The mental health data assessed by (An et al., 2020) reported prevalence of depression in junior nurses and tertiary hospital to be 66.1% and 89.8%, respectively Another study reported that nurses suffered from depression and anxiety more as compared to doctors 71.8%, 16.7% and 70%, 19.1%, respectively. (Zhu et al, 2020). A similar study was conducted by Xiao et al., 2020 reported prevalence of depression/anxiety in nurses (62.4%/58.5%) more as compared to doctors (53.7%/49.5%). As per (Wankovicz et al., 2020), 99.51% FHW had PHQ-9 ≥4 and 99.03% staff had GAD-7≥ 4. Another study used same scale and reported prevalence rate of depression and anxiety to be 90.92% and 73.46% (Pouralizadeh et al., 2020). Cross-sectional study conducted by Sahin et al., 2020 estimated that 77.6% of HCW had depression 60.2% had anxiety. Another study used DASS-21 scale to measure anxiety and depression and reported that 72.3% HCWs had depression whereas 85.7% had moderately-extreme severe anxiety (Sandesh et al., 2020). A study reported that 82.31% of HCW suffered from depression and 61.40 % had anxiety (Liang et al., 2020). Lee et al, 2020 reported HADS-D >11 was 39.3%% and < 11 was 18.1% and HADS-A > 11 was 19.2% and <11 was 38.1% in physicians, HADS-D<11 was 11% and > 11 was 30.7% for HADS-A >11 was 11% and < 11 was 31.4% (Lee et al., 2020) (Table 2).

There were 7 studies which solely analyzed the occurrence of anxiety in FHW due to COVID-19. Alenazi et al., 2020 reported the overall prevalence of anxiety in nursing group three times (27.90%) than medicine group (9.91%). A similar study showed the prevalence of anxiety in nurses (35.98%) to be more as compared to doctors (25.26%) by having 35.98% of anxiety (X. Yang et al, 2020). Another study showed that overall 39.81% healthcare employees had anxiety, from which 53.3% suffered severe, 40.4% moderate and 38.2% mild anxiety (Ayhan baser et al., 2020). Another study conducted in Philippines reported that 37.8% nurses had anxiety (Labrague and De los Santos, 2020). (Korkmaz et al., 2020) revealed that 70.71% healthcare workers experienced anxiety out of which 13% had severe anxiety, 20% had moderate and 38% mild levels of anxiety. According to Liang et al., 2020 and liu et al, 2020 HCWs experienced 12.5% of anxiety. (Table 2).

### 3.5 Insomnia

Eighteen cross-sectional studies estimated the prevalence of insomnia and poor sleep quality among the FHW (Table 2). Various studies reported prevalence of insomnia among 32-57% FHWs (Herrero San martin et al., 2020; Zhou et al; Zhang et al, 2020; Janato iddrissi et al, 2020). Another study showed that 6.2% HCW had less than 6 hours of sleep, 11.3% frequently had sleep disturbances, 2.7% had very bad sleep satisfaction, and 6.7% had difficulty in falling asleep (Yin et al., 2020). One study disclosed that 50.4% HCW had insomnia, out of which 2.1% had severe insomnia, 38.5% had sub-threshold and 9.55% had moderately severe insomnia (Sahin et al., 2020). About 8.27% HCW had ISI score ≥ 22, showing sleeplessness (Rossi Rodolfo et al., 2020). A similar study showed prevalence of 43.04% of insomnia in FHW, out of which 11.2% had moderate insomnia and 31.81% had mild symptoms of insomnia (Liang et al., 2020). Insomnia severity index of 11% nurses and 11.4% doctors came out to be more than 14 i.e. ISI>14. (Bhargava et al., 2020) and (Cheng et al., 2020) evaluated the prevalence of 30% poor sleep quality and insomnia in HCW. Another study showed that nurses (60.97%) had more insomnia than doctors (39.02%) (S. Wang et al, 2020). Another study reported that 1.6% HCW suffering from insomnia had severe depression, 5.8% had severe anxiety and 2.63% suffered from severe stress. Contrary to other studies this showed that doctors (50.52%) had more insomnia than nurses (24.21%) (Wasim et al., 2020) (Table 2).

### 3.6 Stress

Twenty studies assessed the stress level of FHW. Various studies reported the prevalence of stress among HCW in range of 60.8%-90.1% (Zhu et al., 2020; Luseno moreno et al, 2020; (Sandesh et al., 2020). Another study demonstrated that 82.57% of HCW had stress, out of which 3.7% had high stress levels and 78.9% had moderate stress (Wilson et al., 2020). Luseno moreno et al, 2020 revealed that 82.98% HCW were dealing with PTSD whereas Wang et al., 2020 reported only 15.4% FHW had PTSD. From one study it was disclosed that high stress levels were indicated in nurses (81.5%) than doctors (11.5%) (Li et al, 2020). Other studies also showed prevalence of stress and PTSD among HCW in the range of 21.7%-28.9% (Almater et al., 2020; Alshekaili et al., 2020a; Fauzi et al., 2020; Johnson et al., 2020; Kannampallil et al., 2020; Wang et al., 2020) (Table 2).

### 3.7 Distress

Seven studies evaluated the prevalence of distress among HC W. Chen et al, 2020 estimated that besides anxiety disorders suffered by frontline workers which came out to be 32.5%, 28.2% participants had psychological distress. Another study analyzed that physicians (62.0%) suffered more distress than residents (58.2%) (Civantos et al., 2020). Lai et al., 2020 examined distress using IES-R scoring tool and reported that 31.66% FHW were in distress along with 24.26% depression and 21.40% anxiety. Prasad et al., 2020 highlighted that 84.1% nurses tolerated distress during pandemic whereas Nie et al., 2020 reported prevalence rate of 25.1% of distress among nurses. In a similar article, 76.4% distress prevalence was observed in HCW (Şahin et al., 2020a). Yanez et al., 2020 used K-6 scoring tool and found that HCW suffered more with distress (26.1%) than anxiety (2.7%) (Table-2).

### 3.8 Other psychological symptoms

(Antonijevic et al., 2020b) evaluated resilience in HCW to be 10.33% whereas somatization, peritraumatic dissociation estimations were assessed by (Azoulay et al., n.d.) (Bhargava et al., 2020). Burnout (Civantos et al., 2020), suicidal ideation, (Hong et al., 2020)(Azoulay et al., n.d.), somatization and obsessive-compulsive disorder in FHW was assessed by various studies (H. Cai et al., 2020; Juan et al., 2020; B. Zhou et al., 2020)(Xiaoming et al., 2020). Zhang et al., 2020 in his study showed the phobic anxiety encountered by medical healthcare workers (Table 2).

## 4. DISCUSSION

Healthcare workers has acknowledged the outspread of COVID-19 conscientiously (Wasim et al., 2020). Medical staff during the course of pandemic went through exorbitant levels of anxiety, depression, insomnia, somatization and obsessive compulsive disorders and were also prone to grow high grade mental disorders (W. R. Zhang et al., 2020). Thus, in the context of the COVID-19 pandemic, this timely review is both relevant and urgent.

HCWs are presented with unique set of challenges and stressors which include uncertainties such as duration, lack of adequate PPE, gowns, respiratory mask, and face shields, magnitude of crisis, unpreparedness, health threats, transmission risk to loved ones and colleagues (Albott et al., 2020; Anmella et al., 2020; Hall, 2020; Shaukat et al., 2020; Walton et al., 2020). Family safety is the major concerns thus limiting their physical contact with family members to minimize the risk of viral transmission (Albott et al., 2020; Hall, 2020; Jun et al., 2020; Walton et al., 2020). Difficulty in falling asleep, lethargy, irritability generalized anxiety, dejection and feeling submerged because of daily demands served as factors for inconvenience (W. R. Zhang et al., 2020). Also, continuous influx of challenges in treating patients leads to mental exertion, boredom and decreased interest (Walton et al., 2020).

The reasons for their mental health disturbance are due to ambiguity in job role and poor shifts management, leading to lessened efficiency and burnout (Selamu et al., 2017)(Anmella et al., 2020)(AlAteeq et al., 2020). There is need to monitor these psychological morbidities on regular basis.

One study showed that, anxiety and depression levels among medical staff were higher than the national norms (Cheng et al., 2020). Unmarried or single FHWs, due to lower professional enervation and household liability, indicated surege in mental health problem (Z. Zhu et al., 2020c). According to a survey, anxiety levels showed the drift from first increasing then decreasing and was associated with the age, profession and education. Constant distress, worry, irritability, disturbed concentration, work alteration, difficulties in interpersonal relationships, somatization and depression, imposed uncertainty, and burdened HCW ((Albott et al., 2020)(Elhadi et al., 2020a).

Levels of depression and anxiety were also seen among the ICU professionals. A clinical study showed that female nurses were found to be more anxious than doctors. Also, young age appeared to be the risk factors for anxiety and depression (Caillet et al., 2020a). In other studies, incidences of anxiety levels were less among the trained staff working in close proximity with COVID patients. The psychological end results of the pandemic should make us question about the individualized care to HCWs (Caillet et al., 2020b)(Buselli et al., n.d.)2020; (Gallopeni et al., 2020); (Elhadi et al., 2020a).

Also, FHWs had reduced confidence than non-FHW in handling virus. Experiences HCW had less mental health issues as compared to interns (M. Zhang et al., 2020) Good training and relevant knowledge of the disease can effectively lessens the expanse of anxiety, consternation and precariousness (Yifang Zhou et al., 2020). Less educated workers were more likely to get insomnia (Wasim et al., 2020). A survey of 534 pediatric medical staff estimated that outpatient emergency departments and those having bachelor’s degree had high anxiety (Cheng et al., 2020).

Results from the studies conducted in China and other pandemics in the past indicated that FHW had higher distress levels in pandemic (J. Lai et al., 2020); (Prasad et al., 2020). Clinical PTSD symptoms were developed in those who had sub-clinical symptoms due to work setting and continuous challenges and traumatic occurrences, thus there is a need to monitor the sub-clinical symptoms among the persons treating COVID-19 patients (Johnson et al., 2020)(Caillet et al., 2020a). High level of stress lead to burnout which accentuate the exploration of interdependence between burnout, stress and PTSD (Johnson et al., 2020). Supplications have been made in several papers that mental aids should be called for HCWs and particular measures should be taken on psychiatric illness so as to reduce PTSD (Johnson et al., 2020). Women were seen more vulnerable in developing PTSD and required adequate social support by partners (Di Tella et al., 2020). Clinical study showed that, the prevalence of psychological distress among FHW was more than in general population in China (Nie et al., 2020).

In another study it was also reported that people were afraid to meet the nurses and doctors as they feared that they might carry virus with them and can transmit disease to them. FHWs also had to handle the stigmatization from the public which lead to psychological stress eventually (Yifang Zhou et al., 2020). Another considerable apprehension was mistreatment and abuse (either physical or verbal). About 52.3% physicians disclosed minimum one episode of certain kind of abuse. About 31% of the physicians experienced stigmatization by patients (Elhadi et al., 2020a).

Sleep disturbances among HCWs are individualistically associated with depression. About, 38% HCWs suffered from sleep disturbances (Wang et al., 2020). In accordance with a cross-sectional survey, conducted in China among HCW who were directly involved with COVID-19 infected patients, 50.4% suffered from depression, 44.6% from anxiety, 34% from insomnia and 71% from distress (Wańkowicz et al., 2020). The collaborative pervasiveness of sleep disturbances according to a recent systematic review and meta-analysis established that prevalence of sleep disorders among Chinese FHWs was found to be 39.2%. US doctors working in emergency departments had 31% of poor sleep quality and among Pakistani physicians it was 36.8% (Qiu et al., 2020)(Yifang Zhou et al., 2020).

Belief in conspiracy theory in regard to COVID-19 origin were considered to be related to the weak mental health (X. Chen et al., 2020). Service factors which include lack of quality training being given to HCW in providing mental healthcare in isolation units in hospitals, as in this situational setting of pandemic when transmission rates are high and people are dying, there are common mental health problems which are being faced by both patients and care providers. Lack of organizational support is also the major reason for lack of confidence and burnout and psychological morbidities. Inexperienced doctors with inadequate training, unqualified hand washing and improper knowledge of infection control are the factors responsible for the increment of psychological disturbances among medical staff contributing to the development of risks in their life (Xiang et al., 2020);(Kisely et al., 2020); Shaukat et al., 2020; (Albott et al., 2020); (Jun et al., 2020); (Y. Wang et al., 2020b)(Keubo et al., 2020); (Krishnamoorthy et al., 2020).

## 5. CONCLUSION

Medical staff has suffered from high degrees of mental disturbances due to miscellaneous inducements. Governing the robustness among FHW during this time of universal calamity caused due to outbreak of SARS-CoV-2 is very crucial. Many avoidable particulars can be contemplated as the components which are being used to control the current situation of the widespread are burdensome to recast. The prevailing phobia, can be cut down by the proper propagation of knowledge, information and resources of the current situation. Stigma, intolerance and prejudices can be subjugated by educating the healthcare workers, their families and general population. Health policy makers must weigh up the seriousness of the preventive approaches to attenuate the psychological manifestations. HCWs need to be emancipated and their roles and efforts are needed to be acknowledged. Psychological support and potent course of action should be available promptly at multifarious modalities as they play a condemnatory part at odds with COVID-19.

## Data Availability

All data produced in the present work are contained in the manuscript

## Acknowledgements

None.

## Contributors

**Rhythm Joshi**: Conceptualization, Methodology, Data Curation, Writing-Original Draft, Visualization. **Nidhi.B.Agarwal**: Validation, Writing-Review and Editing. **Dinesh Bhurani:** Writing-Review and Editing. **Mohd Ashif Khan**: Conceptualization, Methodology, Data Curation, Writing-Original Draft, Writing-Review and Editing, Visualization, Validation

## List of abbreviation

COVID-19: Coronavirus disease 2019
HCWs: Healthcare workers
FL: Frontline
GAD-7: Generalized anxiety disorder assessment scale
PHQ-9: patient health questionnaire
HADS: Hospital anxiety depression scale
ISI-7: Insomnia severity index
PLC-5: post traumatic stress disorder checklist
IES-R: Impact event scale revised
PHQ-5: Patient health questionnaire
Y-BOCS: Yale brown obsessive compulsive scale
ESS: Epworth sleepiness scale
HADS: Hospital anxiety depression scale
BDI-IA: Beck Depression Inventory
PSQI: Pittsburgh sleep quality index
HARS: Hamilton Anxiety rating scale
NASA: TLX-NASA task load index
STAI-Y1: State-Trait Anxiety Inventory (STAI)
BDI-II: Beck depression inventory-II
DASS-21: Depression, anxiety and stress scale-21 items
PSS-14: The Perceived stress scale 14-item
PSS-10: The perceived stress scale 10-items
BAI: Beck anxiety inventory
PDEQ: Peritraumatic dissociative Experience questionnaire
PSI WHOQL-BREF: World Health organization quality of life instrument
GHQ-12: General Health questionnaire 12-item scale
PHQ-2: Patient health questionnaire 2-item scale
GSI: Global severity index scale 53 item
GAD-2: Generalized anxiety disorder 2-item scale
Pro QoL: Professional Quality of life scale
SAS: Self-rating anxiety scale
EQ-5D-3L: Health status assessment questionnaire of the patient.
ED: Emergency department
ALP: Assistant Laboratory personnel
AN: Assistant nurse
N: Nurse
MD: Medical doctor
Psy: Clinical Psychologist
MLT: Medical laboratory technician
HT: Health Technician

